# Safety, tolerability and immunogenicity of SPVX02, a room temperature-stabilised Tetanus-Diphtheria vaccine compared to Tetadif and diTeBooster: a multicentre, phase 1, blinded, randomised clinical trial

**DOI:** 10.64898/2026.03.09.26347956

**Authors:** Thomas A.N. Reed, Patrick Clarke, Juana de la Torre Arrieta, Moona Razzaque, Arcadio Garcia de Castro, Rossen Alexiev, Catalina Panainte, Naimat Khan, Laura Grey, Mary Matheson, Hannah Cuthbertson, Ozgur Tuncer, Steve Chatfield, Bruce Roser, Alan Boyd, Jonathan S. Nguyen-Van Tam, Karen O’Hanlon, Adam P. Dale, Saul N. Faust

## Abstract

**Background:** Cold chain requirements limit vaccine accessibility and deployment. SPVX02 (Stablepharma Ltd), is a lyophilised, fridge-free version of Tetadif (BB-NCIPD EAD) tetanus-diphtheria vaccine, stable for at least 18 months at temperatures up to 30°C.

**Methods:** A multicentre, first-in-human phase 1, blinded, randomised clinical trial to evaluate the safety and tolerability of SPVX02 compared to existing approved tetanus-diphtheria (Td) vaccines was conducted. Healthy adults aged 18-55 (BMI <=32 kg/m^2^) who had received Td vaccination >=10 years previously were randomly assigned (1:1:1) to receive SPVX02, Tetadif, or diTeBooster (AJ Vaccines A/S). Participants and all investigatory staff were blinded to treatment allocation. Primary outcomes were incidence of adverse events during the trial period including incidence of adverse events reported in participant diaries for 7 days post-dose. Secondary outcomes were day 28 seroprotection rates. Analyses were descriptive. The trial is registered with ISRCTN (98920861).

**Findings:** Between April 1st and September 22nd, 2025, 120 healthy volunteers were screened and sixty participants enrolled at two of three sites. The demographic characteristics of participants were equivalent between groups. No serious adverse reactions, suspected unexpected adverse reactions, or serious adverse events occurred. Fifty-four participants experienced mild or moderate adverse events (AEs); none were severe (grade 3 or higher) AEs. Reactogenicity and tolerability profiles were similar across all groups. All participants had anti-tetanus toxoid (TT) levels >=1·0 IU/ml at Day 28. All participants in both the SPVX02 and Tetadif groups and 19 (95%) in the diTeBooster group had anti-diphtheria (DT) toxoid levels >=0·1 IU/ml at Day 28.

**Interpretation:** SPVX02 is safe, well tolerated, with TT and DT immunogenicity similar to approved Td vaccines. This trial provides first-in-human evidence that StablevaX (Stablepharma, UK) technology can safely reformulate an aluminium-adjuvanted vaccine stable up to 30°C for 18 months.

**Funding:** Funded by Innovate UK Smart Grant project #10083165 and Stablepharma Ltd.

**Research In Context:** *Evidence before this study:* We searched PubMed for randomised controlled trials with thermostable vaccines between database inception and June 2024 using the terms (“temperature stable”) OR (“thermostable”) AND (“vaccin*”) OR (“thermostable vaccine”) with no language restrictions. We identified 33 publications which described various in vitro and in vivo studies that have been performed by researchers as part of their efforts to develop thermostable vaccines. We also identified 2 publications which described randomised, controlled clinical trials that were conducted with thermostable vaccines; (1) a phase 2 clinical trial with ROTASIL, an oral rotavirus vaccine (Isanaka et al, NEJM, 2017) which is now licenced and distributed as a refrigerated product that must be stored but must be stored at 2-8°C; and (2) a phase 1 clinical trial with a current unapproved TB vaccine candidate, ID93+GLE-SE (Sagawa et al, Nature Comms, 2023) where there is no current public information regarding vaccine tolerance to freezing.

*Added value of this study:* This first in human study demonstrates that the reformulated thermostable aluminium hydroxide adjuvanted tetanus-diphtheria vaccine, SPVX02, is safe, well tolerated, and can boost immune responses to tetanus and diphtheria to similar levels as approved comparator vaccines. This vaccine can be stored and distributed at room temperature and is not affected by freezing. A larger Phase 2/3 trial is now planned to confirm these findings prior to consideration for market authorisation.

*Implications of all the available evidence:* The evidence presented here demonstrates that StablevaX technology can be successfully utilised to reformulate a vaccine to be thermostable at room temperature for an extended period of time, without compromising reactogenicity or immunogenicity. While we present data pertaining to a single vaccine, the reformulation and lyophilisation technology underpinning SPVX02 can be applied to many liquid vaccines and biological products. The WHO Immunization Agenda 2030 sets out the global strategy to improve vaccine access in resource-limited settings, prevent vaccine wastage and to reduce the logistical, financial and environmental impact of cold chain requirements. If proven successful across a broader range of vaccine products this technology has potential to significantly benefit global health.

## Introduction

Vaccine preventable diseases continue to affect a large proportion of the global population.^1–3^ Maintaining the cold chain presents a significant challenge to vaccine accessibility and deployment which many vaccination programmes are unable to sufficiently address, particularly when public health infrastructure is fragile.^4^ Most whole pathogen and subunit type vaccines require storage at between 2°C to 8°C, and nucleic acid vaccines commonly require short or longer-term storage at temperatures as low as −60°C to −80°C.^5,6^ Worldwide, a significant number of vaccine doses are spoiled due to over-heating or freezing during storage and transportation.^7^ The environmental impact of the cold chain is considerable as it accounts for a large proportion of energy consumption across the vaccine lifecycle and generates greenhouse gas emissions through refrigeration units and production of dry ice used for transportation.^8,9^ Modelling has shown that use of vaccines that do not rely on the cold chain could provide significant healthcare cost and productivity savings.^10^

Lyophilisation, a method of freeze-drying, is increasingly being used as a tool to improve the thermal stability of pharmaceutical products, including vaccines.^3^ However, to date, all marketed lyophilised vaccines still need to be stored at carefully controlled cold temperatures in a fridge or freezer with their diluent often stored separately.^3^

Tetanus and diphtheria are bacterial infections with significant vaccine-preventable morbidity and mortality.^3^ Globally, 79 million women and their babies remain unprotected against tetanus, and a recent study predicted that eliminating maternal and neonatal tetanus in countries where immunisation coverage is low would result in approximately 70000 prevented deaths over 10 years.^11^ Diphtheria has an overall mortality rate of 5-10% with the largest mortality burden in younger children.^12^ Although vaccination has considerably reduced the morbidity and mortality, diphtheria still poses a serious problem for child health in countries with poor immunisation coverage.^13,14^

Tetadif^®^ (BB-NCIPD EAD, Sofia, Bulgaria) is a WHO-prequalified aluminium-hydroxide adjuvanted combined tetanus and diphtheria vaccine with a well-established safety and immunogenicity profile that requires cold chain storage at 2-8°C and is destroyed by freezing. SPVX02 (StablevaX, Stablepharma Ltd., Bristol, United Kingdom) is a reformulated and lyophilised, fridge-free version of Tetadif which can be stored at temperatures up to 30°C and is not degraded by freezing.^15^ When stored at 30°C for 18 months, SPVX02 has demonstrated consistent levels of immunogenicity and potency in animal model challenge studies that are comparable to refrigerated Tetadif vaccine. From a practical perspective, reconstitution of SPVX02 only requires water for sterile injection. We conducted a phase 1, first-in-human trial to evaluate the safety, tolerability, and immunogenicity of SPVX02.

## Methods

### Design and participants

This phase 1, single-blind, randomised, active-controlled trial aimed to evaluate the safety and tolerability of SPVX02 (batch P126404A) compared with Tetadif (batch D0724-02) and diTeBooster (batch DT411A, AJ Vaccines A/S, Copenhagen, Denmark) in healthy adults. Immunogenicity was assessed as a secondary endpoint. The control vaccines are both well-established tetanus and diphtheria booster vaccines. Tetadif has a marketing authorisation in Bulgaria and is WHO pre-qualified, and diTeBooster has marketing authorisations in the European Union.^16^

Participants were enrolled at the NIHR Southampton Clinical Research Facility, University Hospital Southampton NHS Foundation Trust, UK, and at the Medicines Evaluation Unit, Manchester, UK. Six participants were screened at Queen Alexandra Hospital Portsmouth, UK, but not enrolled. All participants provided written informed consent before trial enrolment. The trial was reviewed and approved by the NHS Research Ethics Committee (London-London Bridge, 25/LO/0059) and the UK Medicines and Healthcare Products Regulatory Agency and is now closed to participants. ISRCTN registration: 98920861.

Healthy adults (aged 18-55 years) with BMI ≤32 kg/m^2^ and history of previous tetanus and diphtheria primary and/or booster immunisation were recruited. Previous immunisation status was confirmed by checking primary care immunisation records, or in the absence of complete vaccination records, by detection of anti-tetanus and anti-diphtheria serum antibody levels ≥0·01 IU/ml. Participants were excluded if they had received a tetanus or diphtheria vaccination within 10 years prior to enrolment. Other exclusion criteria included history of allergy or adverse event following receipt of any components of the trial vaccines, clinically significant systemic disease, and immunosuppression. Women of childbearing potential were required to have a negative pregnancy test at screening and on vaccination days, and to commit to effective contraception or true abstinence throughout study participation. A full list of inclusion and exclusion criteria are listed in the trial protocol (See Clinical Study Protocol).

### Randomisation and masking

Participants were randomly assigned via a predetermined schedule (S-cubed Biometrics, UK) in a 1:1:1 ratio to receive SPVX02, Tetadif, or diTeBooster. A sentinel group of 3 participants was enrolled and randomised, 2 participants received SPVX02, and 1 participant received Tetadif. Following completion of dosing and observations in the sentinel group of 3 participants, and with agreement from the Local Safety Committee, the remaining 57 participants were randomised ensuring an overall 1:1:1 ratio was maintained (See Clinical Study Protocol).

The clinical trial team comprised unblinded and blinded members, with unblinded members responsible only for the dispensing and administration of trial vaccines. All other trial components, including assessment of safety and reactogenicity, were performed by blinded team members. The generation of clinical and laboratory data were performed by fully-blinded team members prior to database lock.

### Procedures

SPVX02 was supplied as a lyophilised powder formulation in a single dose borosilicate glass vial. It was reconstituted with 0·55 ml of water for injection to provide a 0·6 ml suspension for injection. This gave sufficient volume to provide a dose of 0·5 mL, containing ≥20 IU (86·5 IU; 10 Lf) of purified tetanus toxoid, ≥2 IU (11·76 IU; 3·0 Lf) of purified diphtheria toxoid, ≤1·25 mg of aluminium hydroxide, sodium chloride and trehalose. Tetadif was supplied in single dose ampoules of 0·5 ml containing ≥20 IU (82·22 IU; 10 Lf) of purified tetanus toxoid, ≥2 IU (10·74 IU; 3·0 Lf) of purified diphtheria toxoid, ≤1·25 mg of aluminium hydroxide, and ≤0·05mg of thiomersal. DiTeBooster was supplied in single 0·5 ml dose prefilled syringes containing ≥20 IU (≥40 IU; 6·25 Lf) of purified tetanus toxoid, ≥2 IU (21 IU; 6·25 Lf) of purified diphtheria toxoid, 0·5 mg of aluminium hydroxide (See Supplementary File, pages 16-43). Throughout the trial SPVX02 was stored at room temperatures not exceeding 30°C, while Tetadif and diTeBooster vaccines were stored according to their marketing authorisation at 2-8°C.

To assess participant safety for the first in human SPVX02 formulation, the first three participants dosed in the study formed a sentinel group whereby 2 participants received the SPVX02 and 1 received Tetadif. Blinded safety data for these three participants were reviewed by the Local Safety Committee prior to the enrolment of any further participants. Following Local Safety committee approval, the remaining participants were then recruited.

Eligible participants were administered a single dose of allocated trial vaccine by intramuscular deltoid injection on day 1 at the trial site. Participants were observed for 30 minutes after vaccination for the occurrence of any immediate adverse events (AEs).

Participants were called by telephone on day 2 and visited the trial site again on day 7 and day 28. Unsolicited AEs were monitored up to day 28, while solicited AEs were monitored over seven days following vaccination using an electronic diary (OpenClinica, USA; software hosted on UK-based servers). Solicited events included localised symptoms (pain, swelling, tenderness, warmth, and erythema at the injection site), and generalised symptoms (fever, chills, myalgia, fatigue, headache, arthralgia, and rash).

Physical examination, including the measurement of vital signs (oral or tympanic temperature, blood pressure, heart rate, and respiratory rate), was done at screening and on day 1 prior to dosing and was carried out at the investigator’s discretion during the follow-up period. Blood and urine samples were collected for safety laboratory tests at screening and on day 1 and day 28.

Immunogenicity assessments were conducted on blood samples collected at day 1, and day 28. Anti-tetanus toxoid (TT) and anti-diphtheria toxoid (DT) IgG concentrations were quantified using validated enzyme linked immunosorbent assays (ELISAs), calibrated to WHO/NIBSC International Standards (Vaccine Development and Evaluation Centre, UK Health Security Agency Porton Down, UK. See Supplementary File, page 44).

Anti-diphtheria neutralising antibodies were assessed in serum samples collected at 28 days following a single dose of SPVX02, Tetadif or diTebooster. The Vero cell toxin neutralization test (TNT) was performed to measure the level of functional neutralising antibodies against diphtheria toxin. The assay was calibrated to WHO/NIBSC international standards and performed by the Respiratory and Vaccine Preventable Bacteria Reference Unit (UK Health Security Agency, London, UK. See Supplementary File, pages 45-51).

### Outcomes

Primary endpoints were the incidence of unsolicited AEs during the 28 day trial period, and the incidence of pre-defined solicited local and systemic reactogenicity AEs collected via participant diaries, observed for 7 days post-dose.

The secondary endpoints were seroprotection rates at 28 days post-dose, defined as serum anti-TT and anti-DT IgG antibody titres of ≥0·1 IU/ml each;^13,17^ longer term seroprotection rates at 28 days defined as serum anti-TT and anti-DT IgG antibody titres of ≥1·0 IU/ml each; and evaluation of geometric mean titre ratios (GMR) of anti-TT and anti-DT antibodies.

An exploratory endpoint included the determination of post-vaccination geometric mean titres (GMTs) of anti-diphtheria toxin neutralising antibodies in serum collected 28 days after a single dose of SPVX02, Tetadif or diTebooster (results shown in Supplementary File, Table S1, page 2).

### Statistical analysis

The sample size of sixty participants was deemed sufficient to evaluate safety and to allow for an impression of immunogenicity, the study was not powered for the testing of any statistical hypotheses. All analyses were conducted as specified in the Statistical Analysis Plan (Supplementary File, pages 52-130) and were reported using SAS software, version 9·4 (or higher) (SAS Institute Inc., Cary, NC, USA).

All primary and secondary endpoints were summarised descriptively, by vaccine group and time point, as appropriate. Categorical variables were summarised by frequency and percentage. 95% Clopper-Pearson confidence intervals were included for the seroprotection rates. Continuous variables were summarised by descriptive statistics. The geometric mean (GM) and geometric standard deviation (GSD) were derived by taking the mean and standard deviation (SD) of the log-transformed variables at each time point, and back-transforming. Similarly, the geometric mean ratio (GMR) of baseline and associated 95% CI for each vaccine group was obtained by deriving the mean, and 95% CI, change from baseline on the log-scale and back-transforming.

### Role of the funding source

The commercial funder (Stablepharma, UK) was involved in trial conceptualisation and design, data interpretation, writing of the trial report, writing of the manuscript, and the decision to submit for publication. The public funder had no role in any of these activities.

## Results

Between 1^st^ April and 22^nd^ September 2025, 120 participants were assessed for eligibility. Sixty participants were excluded (CONSORT diagram, Figure 1). Sixty participants were randomly assigned to each of the three intervention groups. None of the enrolled participants were withdrawn from the trial, and none were lost to follow up. The demographics of participants was similar between groups (Table 1).

**Figure 1.**
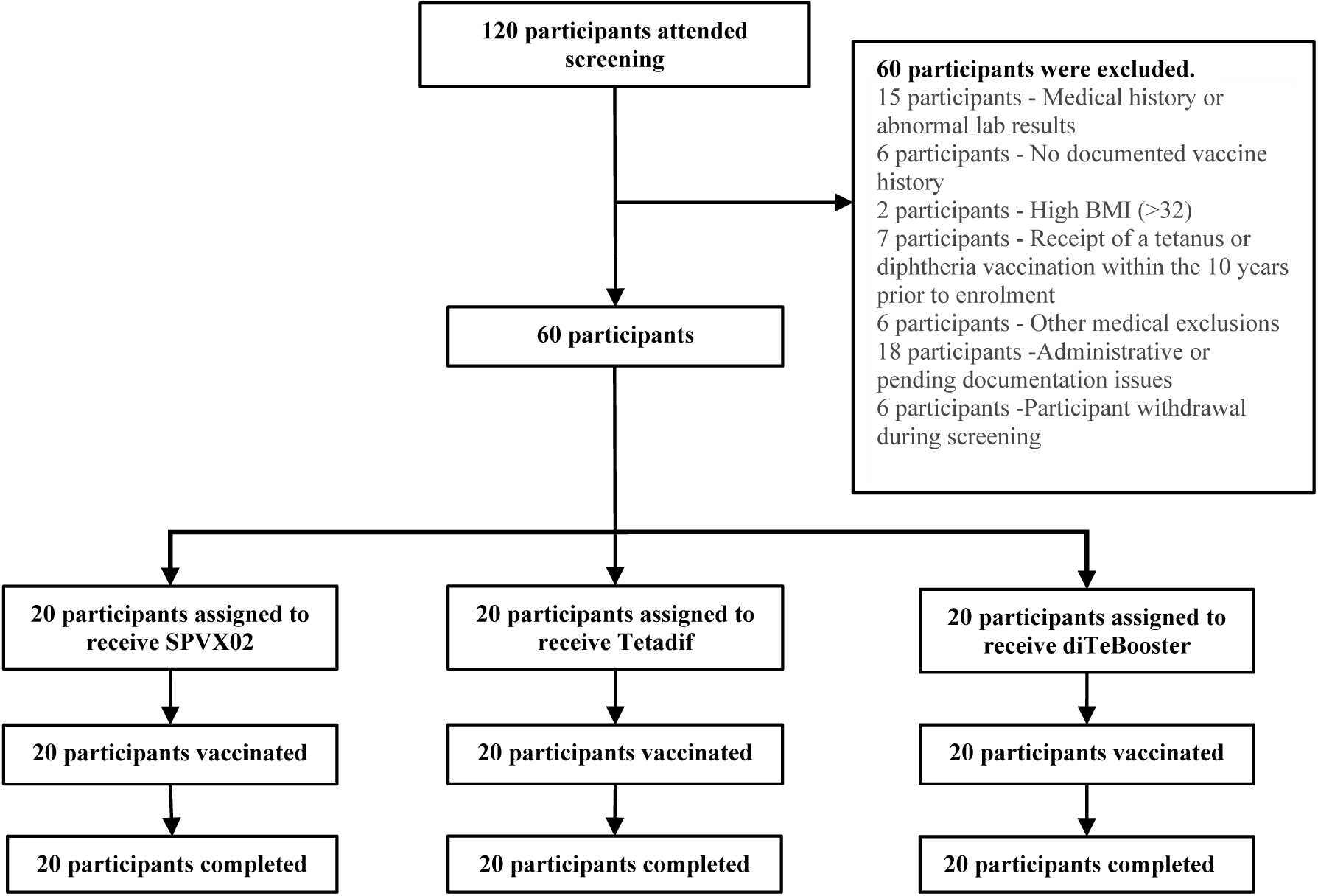
Consort Diagram.

**Table 1.** Demographics.

|  |  |  | **SPVX02**<br>**(N = 20)** | **Tetadif**<br>**(N = 20)** | **diTeBooster**<br>**(N = 20)** |
| --- | --- | --- | --- | --- | --- |
| Sex | Male | n (%) | 10 (50·0) | 11 (55·0) | 9 (45·0) |
|  | Female | n (%) | 10 (50·0) | 9 (45·0) | 11 (55·0) |
| Child Bearing Potential | Yes | n (%) | 9 (90·0) | 8 (88·9) | 10 (90·9) |
| Age at Informed Consent (years) |  | Mean (SD) | 37·6 (6·91) | 35·6 (9·25) | 36·7 (9·67) |
|  |  | Median | 36·5 | 32·5 | 36·5 |
|  |  | IQR | 32·5, 42·0 | 28·5, 41·5 | 29·0, 42·5 |
|  |  | Min, Max | 25, 50 | 23, 54 | 21, 54 |
| Self-Reported Ethnicity | White | n (%) | 19 (95·0) | 19 (95·0) | 19 (95·0) |
|  | Other | n (%) | 1 (5·0) | 1 (5·0) | 1 (5·0) |
| BMI at Screening (kg/m<sup>2</sup>) |  | Mean (SD) | 25·31 (2·98) | 24·34 (2·02) | 24·83 (3·74) |
|  |  | Median | 24·70 | 24·50 | 25·10 |
|  |  | IQR | 23·40, 26·65 | 22·50, 25·35 | 21·85, 28·00 |
|  |  | Min, Max | 21·3, 32·0 | 21·8, 29·0 | 18·3, 31·0 |

SD= standard deviation
IQR= interquartile range
BMI= body mass index

All three study vaccines were well tolerated with similar frequencies of treatment-emergent AEs recorded between vaccine groups (Table 2). Most participants experienced solicited AEs. Forty participants (66·7%) experienced pain at the injection site, 19 (31·7%) experienced myalgia, 16 (18·3%) experienced headache, and 10 (16·7%) experienced fatigue following vaccination. Figure 2 shows related adverse events by vaccine and grade.

**Figure 2.**
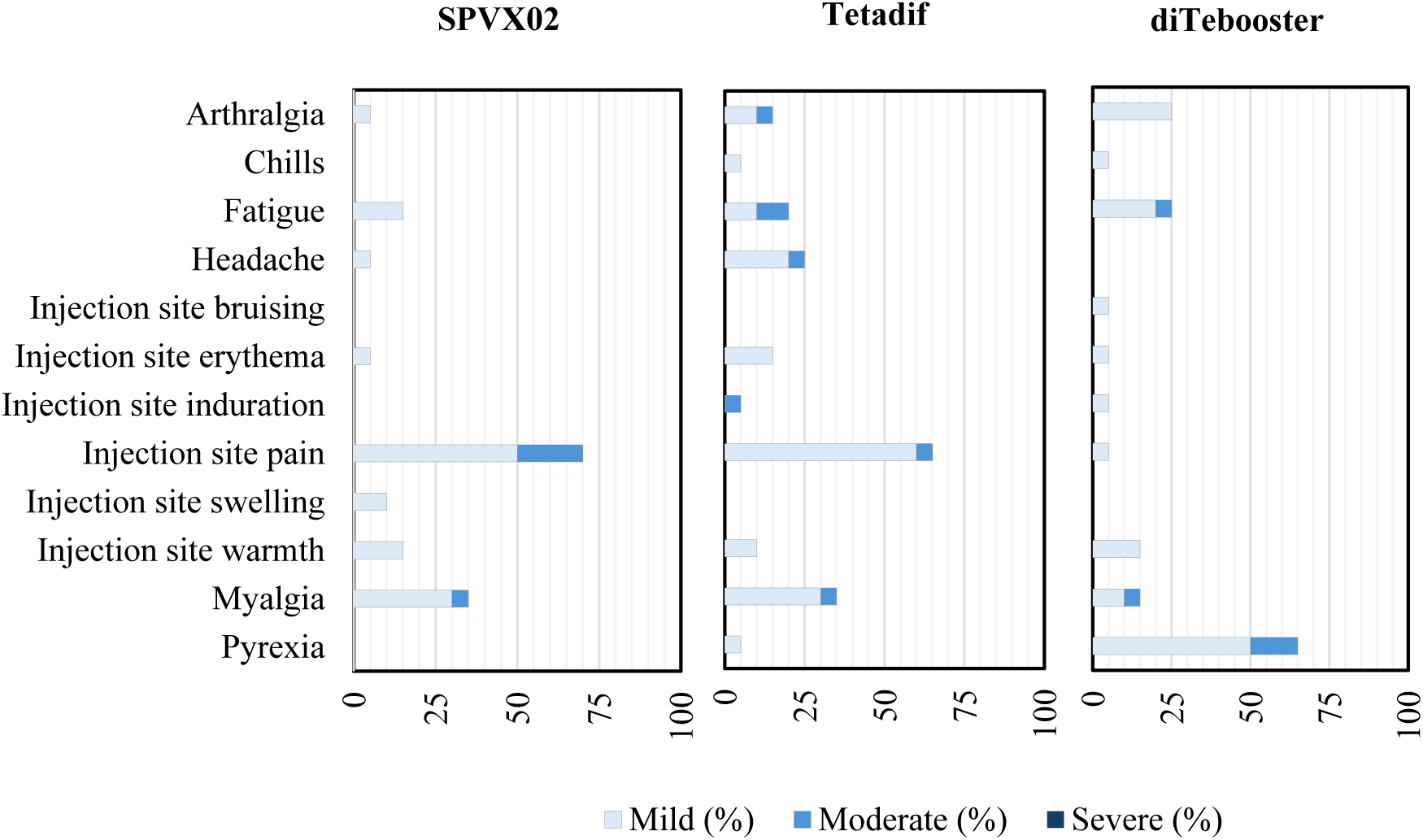
Related adverse events reported up to 7 days post dose. Adverse events were collected for 7 days using participant diaries and are presented as treatment-emergent adverse events (TEAEs). All TEAEs were mild or moderate; no grade 3 or higher events were observed. No skin rash was reported. Tenderness was captured as injection-site pain in the treatment-emergent adverse event classifications.

**Table 2.** Treatment-Emergent Adverse Events.

|  | SPVX02<br>(N=20) |  | Tetadif<br>(N=20) |  | diTeBooster<br>(N=20) |  | Overall<br>(N=60) |  |
| --- | --- | --- | --- | --- | --- | --- | --- | --- |
| System Organ Class<br>Preferred Term [1] | n | % | n | % | n | % | n | % |
| Number of Participants with any Adverse Event (AE) | 19 | 95 | 17 | 85 | 18 | 90 | 54 | 90 |
| Number of participants with vaccine-related AE | 17 | 89.5 | 16 | 94.1 | 18 | 100 | 51 | 94.4 |
| Number of participants with unrelated AE | 2 | 10.5 | 1 | 5.8 | 0 | 0 | 3 | 5.5 |
| **Related adverse events reported up to 7 days post dose** |  |  |  |  |  |  |  |  |
| Arthralgia |  |  |  |  |  |  |  |  |
| Mild | 1 | 5 | 2 | 10 | 1 | 5 | 4 | 6.7 |
| Moderate | 0 | 0 | 1 | 5 | 0 | 0 | 1 | 1.7 |
| Severe | 0 | 0 | 0 | 0 | 0 | 0 | 0 | 0 |
| Chills |  |  |  |  |  |  |  |  |
| Mild | 0 | 0 | 1 | 5 | 1 | 5 | 2 | 3.3 |
| Moderate | 0 | 0 | 0 | 0 | 0 | 0 | 0 | 0 |
| Severe | 0 | 0 | 0 | 0 | 0 | 0 | 0 | 0 |
| Fatigue |  |  |  |  |  |  |  |  |
| Mild | 3 | 15 | 2 | 10 | 2 | 10 | 7 | 11.7 |
| Moderate | 0 | 0 | 2 | 10 | 1 | 5 | 3 | 5 |
| Severe | 0 | 0 | 0 | 0 | 0 | 0 | 0 | 0 |
| Headache |  |  |  |  |  |  |  |  |
| Mild | 1 | 5 | 4 | 20 | 5 | 25 | 10 | 16.7 |
| Moderate | 0 | 0 | 1 | 5 | 0 | 0 | 1 | 1.7 |
| Severe | 0 | 0 | 0 | 0 | 0 | 0 | 0 | 0 |
| Injection site bruising |  |  |  |  |  |  |  |  |
| Mild | 0 | 0 | 0 | 0 | 1 | 5 | 1 | 1.7 |
| Moderate | 0 | 0 | 0 | 0 | 0 | 0 | 0 | 0 |
| Severe | 0 | 0 | 0 | 0 | 0 | 0 | 0 | 0 |
| Injection site erythema |  |  |  |  |  |  |  |  |
| Mild | 1 | 5 | 3 | 15 | 0 | 0 | 4 | 6.7 |
| Moderate | 0 | 0 | 0 | 0 | 0 | 0 | 0 | 0 |
| Severe | 0 | 0 | 0 | 0 | 0 | 0 | 0 | 0 |
| Injection site induration |  |  |  |  |  |  |  |  |
| Mild | 0 | 0 | 0 | 0 | 0 | 0 | 0 | 0 |
| Moderate | 0 | 0 | 1 | 5 | 0 | 0 | 1 | 1.7 |
| Severe | 0 | 0 | 0 | 0 | 0 | 0 | 0 | 0 |
| Injection site pain |  |  |  |  |  |  |  |  |
| Mild | 10 | 50 | 12 | 60 | 10 | 50 | 32 | 53.3 |
| Moderate | 4 | 20 | 1 | 5 | 3 | 15 | 8 | 13.3 |
| Severe | 0 | 0 | 0 | 0 | 0 | 0 | 0 | 0 |
| Injection site swelling |  |  |  |  |  |  |  |  |
| Mild | 2 | 10 | 0 | 0 | 1 | 5 | 3 | 5 |
| System Organ Class<br>Preferred Term [1] | n | % | n | % | n | % | n | % |
| Moderate | 0 | 0 | 0 | 0 | 0 | 0 | 0 | 0 |
| Severe | 0 | 0 | 0 | 0 | 0 | 0 | 0 | 0 |
| Injection site warmth |  |  |  |  |  |  |  |  |
| Mild | 3 | 15 | 2 | 10 | 3 | 15 | 8 | 13·3 |
| Moderate | 0 | 0 | 0 | 0 | 0 | 0 | 0 | 0 |
| Severe | 0 | 0 | 0 | 0 | 0 | 0 | 0 | 0 |
| Myalgia |  |  |  |  |  |  |  |  |
| Mild | 6 | 30 | 6 | 30 | 4 | 20 | 16 | 26·7 |
| Moderate | 1 | 5 | 1 | 5 | 1 | 5 | 3 | 5 |
| Severe | 0 | 0 | 0 | 0 | 0 | 0 | 0 | 0 |
| Pyrexia |  |  |  |  |  |  |  |  |
| Mild | 0 | 0 | 1 | 5 | 1 | 5 | 2 | 3·3 |
| Moderate | 0 | 0 | 0 | 0 | 0 | 0 | 0 | 0 |
| Severe | 0 | 0 | 0 | 0 | 0 | 0 | 0 | 0 |
| **Adverse events reported up to 28 days post dose** |  |  |  |  |  |  |  |  |
| Back pain |  |  |  |  |  |  |  |  |
| Mild | 0 | 0 | 0 | 0 | 0 | 0 | 0 | 0 |
| Moderate | 0 | 0 | 1 | 5 | 0 | 0 | 1 | 1·7 |
| Severe | 0 | 0 | 0 | 0 | 0 | 0 | 0 | 0 |
| Diarrhoea |  |  |  |  |  |  |  |  |
| Mild | 0 | 0 | 1 | 5 | 1 | 5 | 2 | 3·3 |
| Moderate | 0 | 0 | 0 | 0 | 0 | 0 | 0 | 0 |
| Severe | 0 | 0 | 0 | 0 | 0 | 0 | 0 | 0 |
| Dry eye |  |  |  |  |  |  |  |  |
| Mild | 0 | 0 | 0 | 0 | 0 | 0 | 0 | 0 |
| Moderate | 0 | 0 | 0 | 0 | 1 | 5 | 1 | 1·7 |
| Severe | 0 | 0 | 0 | 0 | 0 | 0 | 0 | 0 |
| Gastroenteritis |  |  |  |  |  |  |  |  |
| Mild | 0 | 0 | 0 | 0 | 0 | 0 | 0 | 0 |
| Moderate | 1 | 5 | 0 | 0 | 0 | 0 | 1 | 1·7 |
| Severe | 0 | 0 | 0 | 0 | 0 | 0 | 0 | 0 |
| Herpes zoster |  |  |  |  |  |  |  |  |
| Mild | 0 | 0 | 0 | 0 | 0 | 0 | 0 | 0 |
| Moderate | 0 | 0 | 1 | 5 | 0 | 0 | 1 | 1·7 |
| Severe | 0 | 0 | 0 | 0 | 0 | 0 | 0 | 0 |
| Hypothermia |  |  |  |  |  |  |  |  |
| Mild | 0 | 0 | 1 | 5 | 0 | 0 | 1 | 1·7 |
| Moderate | 0 | 0 | 0 | 0 | 0 | 0 | 0 | 0 |
| Severe | 0 | 0 | 0 | 0 | 0 | 0 | 0 | 0 |
| Lymph node pain |  |  |  |  |  |  |  |  |
| Mild | 0 | 0 | 0 | 0 | 1 | 5 | 1 | 1·7 |
| System Organ Class<br>Preferred Term [1] | n | % | n | % | n | % | n | % |
| Moderate | 0 | 0 | 0 | 0 | 0 | 0 | 0 | 0 |
| Severe | 0 | 0 | 0 | 0 | 0 | 0 | 0 | 0 |
| Musculoskeletal pain |  |  |  |  |  |  |  |  |
| Mild | 0 | 0 | 0 | 0 | 0 | 0 | 0 | 0 |
| Moderate | 0 | 0 | 1 | 5 | 0 | 0 | 1 | 1.7 |
| Severe | 0 | 0 | 0 | 0 | 0 | 0 | 0 | 0 |
| Nasopharyngitis |  |  |  |  |  |  |  |  |
| Mild | 1 | 5 | 0 | 0 | 0 | 0 | 1 | 1.7 |
| Moderate | 0 | 0 | 0 | 0 | 0 | 0 | 0 | 0 |
| Severe | 0 | 0 | 0 | 0 | 0 | 0 | 0 | 0 |
| Oropharyngeal pain |  |  |  |  |  |  |  |  |
| Mild | 0 | 0 | 0 | 0 | 0 | 0 | 0 | 0 |
| Moderate | 0 | 0 | 1 | 5 | 0 | 0 | 1 | 1.7 |
| Severe | 0 | 0 | 0 | 0 | 0 | 0 | 0 | 0 |
| Otitis externa |  |  |  |  |  |  |  |  |
| Mild | 0 | 0 | 1 | 5 | 0 | 0 | 1 | 1.7 |
| Moderate | 0 | 0 | 0 | 0 | 0 | 0 | 0 | 0 |
| Severe | 0 | 0 | 0 | 0 | 0 | 0 | 0 | 0 |
| Pollakiuria |  |  |  |  |  |  |  |  |
| Mild | 0 | 0 | 1 | 5 | 0 | 0 | 1 | 1.7 |
| Moderate | 0 | 0 | 0 | 0 | 0 | 0 | 0 | 0 |
| Severe | 0 | 0 | 0 | 0 | 0 | 0 | 0 | 0 |
| Presyncope |  |  |  |  |  |  |  |  |
| Mild | 0 | 0 | 1 | 5 | 0 | 0 | 1 | 1.7 |
| Moderate | 0 | 0 | 0 | 0 | 0 | 0 | 0 | 0 |
| Severe | 0 | 0 | 0 | 0 | 0 | 0 | 0 | 0 |
| Pruritus |  |  |  |  |  |  |  |  |
| Mild | 0 | 0 | 1 | 5 | 1 | 5 | 2 | 3.3 |
| Moderate | 0 | 0 | 0 | 0 | 0 | 0 | 0 | 0 |
| Severe | 0 | 0 | 0 | 0 | 0 | 0 | 0 | 0 |
| Rhinitis |  |  |  |  |  |  |  |  |
| Mild | 0 | 0 | 0 | 0 | 0 | 0 | 0 | 0 |
| Moderate | 1 | 5 | 0 | 0 | 0 | 0 | 1 | 1.7 |
| Severe | 0 | 0 | 0 | 0 | 0 | 0 | 0 | 0 |
| Urinary tract infection |  |  |  |  |  |  |  |  |
| Mild | 0 | 0 | 0 | 0 | 0 | 0 | 0 | 0 |
| Moderate | 0 | 0 | 0 | 0 | 1 | 5 | 1 | 1.7 |
| Severe | 0 | 0 | 0 | 0 | 0 | 0 | 0 | 0 |
| System Organ Class<br>Preferred Term [1] | n | % | n | % | n | % | n | % |
| **Laboratory adverse events reported up to 28 days post dose** |  |  |  |  |  |  |  |  |
| Alanine aminotransferase increased |  |  |  |  |  |  |  |  |
| Mild | 1 | 5 | 0 | 0 | 0 | 0 | 1 | 1.7 |
| Moderate | 0 | 0 | 0 | 0 | 0 | 0 | 0 | 0 |
| Severe | 0 | 0 | 0 | 0 | 0 | 0 | 0 | 0 |
| Blood fibrinogen increased |  |  |  |  |  |  |  |  |
| Mild | 0 | 0 | 0 | 0 | 0 | 0 | 0 | 0 |
| Moderate | 0 | 0 | 1 | 5 | 0 | 0 | 1 | 1.7 |
| Severe | 0 | 0 | 0 | 0 | 0 | 0 | 0 | 0 |
| Haemoglobin decreased |  |  |  |  |  |  |  |  |
| Mild | 0 | 0 | 1 | 5 | 0 | 0 | 1 | 1.7 |
| Moderate | 0 | 0 | 0 | 0 | 0 | 0 | 0 | 0 |
| Severe | 0 | 0 | 0 | 0 | 0 | 0 | 0 | 0 |
| Hypophosphatemia |  |  |  |  |  |  |  |  |
| Mild | 1 | 5 | 0 | 0 | 0 | 0 | 1 | 1.7 |
| Moderate | 0 | 0 | 0 | 0 | 0 | 0 | 0 | 0 |
| Severe | 0 | 0 | 0 | 0 | 0 | 0 | 0 | 0 |
| Lymphopenia |  |  |  |  |  |  |  |  |
| Mild | 1 | 5 | 0 | 0 | 0 | 0 | 1 | 1.7 |
| Moderate | 0 | 0 | 0 | 0 | 0 | 0 | 0 | 0 |
| Severe | 0 | 0 | 0 | 0 | 0 | 0 | 0 | 0 |
| Neutropenia |  |  |  |  |  |  |  |  |
| Mild | 1 | 5 | 0 | 0 | 0 | 0 | 1 | 1.7 |
| Moderate | 0 | 0 | 0 | 0 | 0 | 0 | 0 | 0 |
| Severe | 0 | 0 | 0 | 0 | 0 | 0 | 0 | 0 |
| Thrombocytopenia |  |  |  |  |  |  |  |  |
| Mild | 0 | 0 | 1 | 5 | 0 | 0 | 1 | 1.7 |
| Moderate | 0 | 0 | 0 | 0 | 0 | 0 | 0 | 0 |
| Severe | 0 | 0 | 0 | 0 | 0 | 0 | 0 | 0 |

No serious adverse reactions (SARs), suspected unexpected serious adverse reactions (SUSARs), or serious adverse events (SAEs) occurred. In total, 54 participants across all vaccine groups experienced AEs, ranging in severity from mild to moderate. No participant experienced a severe (grade 3 or higher) AE. There were no observed differences in AE profile between vaccine groups (Table 2). Fifty-one participants experienced AEs which were determined to be related to the study intervention with 17, 16, and 18 in the SPVX02, Tetadif, and diTeBooster groups respectively. Six participants had laboratory AEs of mild severity (Four received SPVX02, two Tetadif and zero diTeBooster). There were no laboratory AEs moderate (grade 2) or higher. No SAEs occurred during the study, and all AEs self-resolved, without intervention.

All participants had longer-term seroprotective anti-TT antibody levels ≥1·0 IU/ml at Day 28. This included all 13, 13, and 9 participants who had baseline anti-tetanus antibody titres <1·0 IU/m in the SPVX02, Tetadif, and diTeBooster groups respectively. The ratios of anti-TT baseline GMT to Day 28 GMT were GMR 16·6, 8·5, and 15·5 for SPVX02, Tetadif, and diTeBooster respectively (Table 3).

**Table 3.**
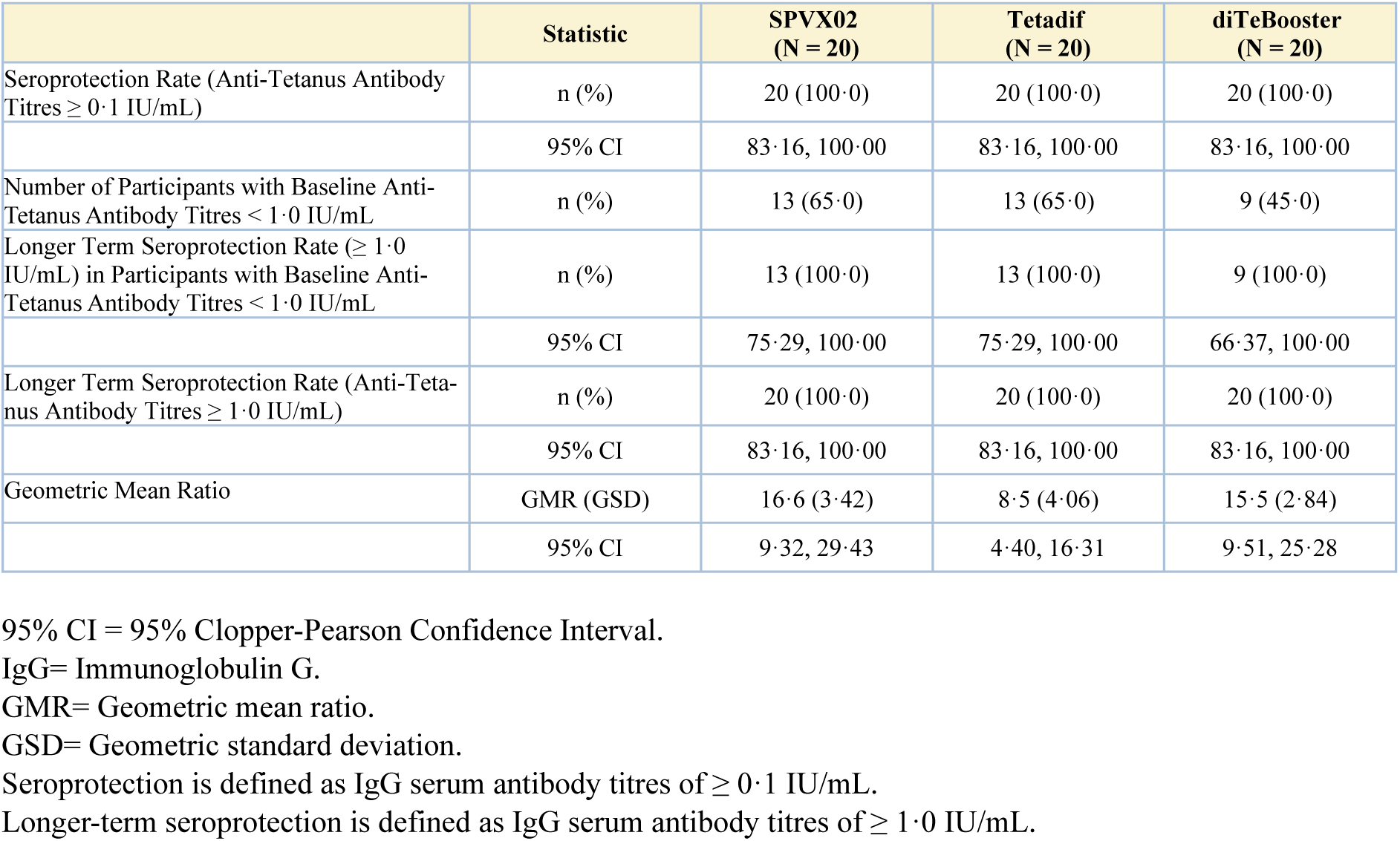
Seroprotection Rates – Tetanus.

All participants in both the SPVX02 and Tetadif groups and 19 (95%) in the diTeBooster group had seroprotective anti-DT antibody levels ≥0·1 IU/ml at Day 28. Sixteen (80%) of the SPVX02 participants, 17 of Tetadif (85%) and 19 of diTeBooster (95%) participants achieved longer-term seroprotective anti-DT antibody levels ≥1·0 IU/ml at Day 28. Of those who had baseline anti-diphtheria antibody titres <1·0 IU/mL, long-term seroprotection at Day 28 was achieved by 13 (76·5%) in the SPVX02, 15 (83·3%) in Tetadif and 18 (94·7%) in diTeBooster group. The ratios of Day 28 GMT to anti-DT baseline GMT were GMR 10·1, 9·3, and 22·8 for SPVX02, Tetadif, and diTeBooster respectively (Table 4).

**Table 4.** Seroprotection Rates – Diphtheria.

|  | Statistic | SPVX02<br>(N = 20) | Tetadif<br>(N = 20) | diTeBooster<br>(N = 20) |
| --- | --- | --- | --- | --- |
| Seroprotection Rate (Anti-Diphtheria Antibody Titres $\geq 0.1$ IU/mL) | n (%) | 20 (100.0) | 20 (100.0) | 19 (95.0) |
|  | 95% CI | 83.16, 100.00 | 83.16, 100.00 | 75.13, 99.87 |
| Number of Participants with Baseline Anti-Diphtheria Antibody Titres $< 1.0$ IU/mL | n (%) | 17 (85.0) | 18 (90.0) | 19 (95.0) |
| Longer Term Seroprotection Rate ($\geq 1.0$ IU/mL) in Participants with Baseline Anti-Diphtheria Antibody Titres $< 1.0$ IU/mL | n (%) | 13 (76.5) | 15 (83.3) | 18 (94.7) |
|  | 95% CI | 50.10, 93.19 | 58.58, 96.42 | 73.97, 99.87 |
| Longer Term Seroprotection Rate (Anti-Diphtheria Antibody Titres $\geq 1.0$ IU/mL) | n (%) | 16 (80.0) | 17 (85.0) | 19 (95.0) |
|  | 95% CI | 56.34, 94.27 | 62.11, 96.79 | 75.13, 99.87 |
| Geometric Mean Ratio | GMR (GSD) | 10.1 (3.36) | 9.3 (3.24) | 22.8 (2.63) |
|  | 95% CI | 5.70, 17.73 | 5.35, 16.06 | 14.54, 35.89 |

95% CI = 95% Clopper-Pearson Confidence Interval.
IgG=immunoglobulin G.
GMR= Geometric mean ratio.
GSD= Geometric standard deviation.
Seroprotection is defined as IgG serum antibody titres of $\geq 0.1$ IU/mL.
Longer-term seroprotection is defined as IgG serum antibody titres of $\geq 1.0$ IU/mL.

The majority of participants across all treatment groups developed neutralising antibody titres for diphtheria consistent with long-term seroprotection. In the SPVX02 group, 18 (90%) had neutralising antibodies levels ≥1·0 IU/ml, with the remaining two (10%) having protective titres ≥0.1 IU/ml. In the Tetadif group, 18 (90%) had neutralising antibodies levels ≥1·0 IU/ml, with the remaining two (10%) having protective titres ≥0·1 IU/ml. In the diTeBooster group, 19 (95%) had neutralising antibodies ≥1·0 IU/ml, with one (5%) showing the lowest level of neutralising antibody titre, indicating some degree of protection (0·064 IU/mL) (Supplementary File, Table S1, page 2).

## Discussion

This first-in-human clinical trial has established the safety and tolerability of SPVX02 and shows for the first time that a room-temperature-stabilized formulation is broadly similar to the immune performance of the globally-marketed Tetadif vaccine that protects against tetanus and diphtheria. Deployment of vaccines that remain stable outside the cold chain could transform global immunisation strategies, particularly in low- and middle-income countries, by improving coverage, reducing waste, and supporting rapid response during outbreaks.

All AEs occurred at similar frequencies in the SPVX02 group compared to both the Tetadif original formulation and the second approved comparator, diTeBooster. Mild to moderate injection site pain was the most commonly reported AE, which occurred evenly across all vaccine groups.

For anti-TT immune responses, SPVX02 showed similar antibody titre boosting rates in participants with anti-TT levels <0·1 IU/ml at baseline compared to the participants in both Tetadif and diTeBooster groups. Anti-TT antibody GMR of day 28 compared to baseline was similar for SPVX02 and diTeBooster. The lower GMR (8·5, 95% CI 4·4-16·3) of Tetadif compared its temperature-stabilised formulation SPVX02 (16·6, 95% CI 9·3-29·4) may be stochastic due to the small sample size.

For anti-DT immune responses, SPVX02 showed similar antibody titre boosting rates and baseline to day 28 GMR as the original Tetadif formulation. Participants who received diTeBooster had higher anti-DT antibody levels at day 28 and higher day 28 GMR than those who received SPVX02 or Tetadif. This is a likely consequence of the quantity and calculated potency of diphtheria toxoid in diTeBooster being approximately double that of Tetadif and SPVX02 (assessed using the European pharmacopoeia diphtheria potency assay (2.7.8): diTeBooster potency 21 IU/0·5mL compared to Tetadif and SPVX02 potency, 10·74 and 11·76 IU/0·5mL) (see Supplementary File, pages 16-21).

In parallel to this study, a stability study programme was conducted (NetpharmaLab, Spain) to demonstrate the robustness of the temperature stabilised SPVX02 formulation when exposed to 18 months storage temperatures of 30°C and 75% relative humidity; and 6 months storage temperatures of 40°C and 75% relative humidity. SPVX02 was also subjected to three freeze-thaw cycles. Across all tested timepoints and temperatures (with data currently available up to 18 months at 30°C and up to 6 months at 40°C) SPVX02 remained within specifications for potency and twenty other attributes with no evidence of degradation or loss of immunological function. These data align with the clinical safety and immunogenicity outcomes observed in the phase 1 clinical trial (see Supplementary File, page 3-15).

The randomised, active-controlled study design was its key strength. Limitations of the study should be discussed. First, the short duration of this study does not permit evaluation of delayed or rare safety events; therefore safety conclusions are limited to short-term reactogenicity and AEs observed within the 28-day study period. However, this approach is consistent with that used in comparable vaccine trials.^18,19^ Second, immunogenicity was only measured through today 28. Nevertheless, the immune parameters assessed are well-established correlates of protection.^20^ Sustained immunity following TT- and DT-containing vaccination has been documented in long-term serological studies, showing that neutralising antitoxin antibodies can persist for many years after primary immunisation, with gradual waning over time.^21^ Third, initially slow recruitment occurred due to limited availability of detailed vaccination history and restrictive body mass index (BMI) eligibility criteria. After enrolment of 19 participants, a protocol amendment broadened the acceptable BMI range in line with current UK population data^22^ and introduced antibody screening as a proxy for prior tetanus and diphtheria vaccination.

In conclusion, we have demonstrated that SPVX02 is safe, well tolerated, and can boost immune responses to TT and DT to similar levels as approved vaccines. While a larger phase 2/3 trial is planned to achieve marketing authorisation, this trial has shown for the first time in humans that the StablevaX (Stablepharma, UK) technology can successfully reformulate a aluminium hydroxide adjuvanted vaccine to be stable at room temperature for an extended period of time that otherwise had to be maintained at 2-8 °C. The reformulation and lyophilisation technology underpinning SPVX02 can be applied to many liquid vaccines and biologic products. By eliminating reliance on the cold chain and preventing degradation due to freezing, SPVX02 and other vaccines subject to StablevaX technology could substantially reduce vaccine wastage and improve access in resource-limited settings.

## Data Availability

All data produced in the present study are available upon reasonable request to the authors

## Author contributions

SNF was the trial chief investigator. TR, APD, JdlTA, KOH, AB, JVT, SNF contributed to the protocol and design of the study. LG designed the statistical analysis plan. TR, PC, APD, MK and SNF recruited participants and gathered trial data. MM and HC generated the immunogenicity data. LG conducted the safety and immunogenicity analyses. TR, PC, APD and SNF drafted the report. All other others contributed to the implementation and data collection. All authors reviewed and approved the final report.

## Declarations of Interest

KOH, JdlTA, AGdC, OT and BR are employees of Stablepharma Ltd. R Alexiev is an employee of BB-NCPID-EAD. SC is the Chairperson of the Stablepharma Ltd Board and is a non-executive director. SNF has advised Stablepharma for 9 years and holds a small quantity of Stablepharma share options. JSN-V-T, AB, and SC, are all remunerated members of the Stablepharma advisory board and each hold a small quantity of Stablepharma share options. LG has served as a consultant to Stablepharma Ltd. APD has received grant funding from ILiAD Biotechnologies and Moderna Ltd, and has performed consultancy work for GSK. He receives no personal financial payment for this work. RA is an employee of BB-NCIPD EAD who manufacture and supply Tetadif bulk vaccine to Stablepharma under licence agreement. SNF acts on behalf of University Hospital Southampton NHS Foundation Trust as an Investigator and/or providing consultative advice on clinical trials and studies of COVID-19 and other vaccines funded or sponsored by vaccine manufacturers including Janssen, Moderna, Pfizer, BioNTech, AstraZeneca, GlaxoSmithKline, Novavax, Seqirus, Sanofi, Medimmune, Merck and Valneva vaccines and antimicrobials. He receives no personal financial payment for this work.

## Acknowledgements

Lisa Berry (Lead phase 1 research matron), Laura Presland and the staff and resources of the NIHR Southampton Clinical Research Facility; Johanna Mouland and the staff and resources of Queen Alexandra Hospital Portsmouth; Deb Henshaw (Head of CRA) and Abbie Lannon (Senior CRA), and the staff and resources of the Medicines Evaluation Unit, Manchester. Immunoassay Group, Clinical Evaluation Team UKHSA conducted the ELISA assays. David Litt, Head, Vaccine Preventable Bacteria Section, Respiratory and Vaccine Preventable Bacteria Reference Unit and Laboratory Surveillance Lead, UK Health Security Agency, London, UK, conducted the neutralising assays. Professor Robert Read, Professor Chris Edwards and Associate Professor Helena Lee served as members of the independent safety monitoring committee. SNF is an NIHR Senior Investigator.

## Notes

### Clinical Trial

ISRCTN registration: 98920861

### Author Declarations

The trial was reviewed and approved by the NHS Research Ethics Committee (London-London Bridge, 25/LO/0059).

