## Supplementary file for "Safety, tolerability and immunogenicity of SPVX02, a room temperature-stabilised Tetanus-Diphtheria vaccine compared to Tetadif and diTeBooster: a multicentre, phase 1, blinded, randomised clinical trial"

##### List of Tables

1. Table S1. Neutralising Antibody Titres for Diphtheria .............................. 2
2. Table S2. In-Vivo Potency (Lethal Challenge Test) of Tetanus and Diphtheria Toxoids in SPVX02 (Lot Number P126402E) .............................. 5

##### List of Figures

1. Figure S1. In-Vivo Potency of Tetanus and Diphtheria .............................. 2

#### 1. Neutralising Antibodies: Post-vaccination Geometric Mean Titres (GMTs) of Anti-Diphtheria

Table S1 presents post-vaccination neutralising antibody responses against diphtheria at Day 28 following administration of a single dose of SPVX02, Tetadif or diTeBooster.

The table includes seroprotection rates and geometric mean titres (GMTs) with 95% confidence intervals.

For the analysis of geometric mean titres, the neutralising antibody titres were log10-transformed prior to analysis. Values above the upper limit of quantification (4.096 IU/mL) were set to the upper limit for calculation purposes. The proportion of participants with neutralising antibody titres ≥4.096 IU/mL was 50% for SPVX02, 45% for Tetadif and 85% for diTeBooster. Geometric mean titres (GMTs) were calculated by back-transformation of the mean log10 titres.

**Table S1. Neutralising antibody titres for diphtheria**

| | **Statistic** | **SPVX02 (N = 20)** | **Tetadif (N = 20)** | **diTeBooster (N = 20)** |
|---|---|---|---|---|
| Seroprotection Rate | n | 20 | 20 | 19 |
| | (%) | (100.0) | (100.0) | (95.0) |
| | 95% CI | 83.16, 100.00 | 83.16, 100.00 | 75.13, 99.87 |
| Longer-Term Seroprotection Rate | n | 18 | 18 | 19 |
| | (%) | (90.0) | (90.0) | (95.0) |
| | 95% CI | 68.30, 98.76 | 68.30, 98.76 | 75.13, 99.87 |
| Geometric Mean Titres | n | 20 | 20 | 20 |
| | GMT | 2.27 | 1.98 | 3.1 |
|  | 95% CI | 1.61, 3.21 | 1.38, 2.84 | 2.06, 4.68 |

Seroprotection is defined as IgG serum antibody titre of >= 0.1 IU/mL.
Longer-term seroprotection is defined as IgG serum antibody titre of >= 1.0 IU/mL.
95% CI = 95% Clopper-Pearson Confidence Interval.
GMT= Geometric mean titres.

#### 2. In-Vivo Potency Challenge Testing Using SPVX02 Drug Product in Stability Studies

A long-term stability study was initiated on 22nd December 2023 to support the development of SPVX02. The stability study assesses the maintenance of SPVX02 vaccine attributes over an extended storage period of up to 4 years. This stability study is ongoing and includes two batches of SPVX02: the engineering batch (Lot Number P126402E; stability study initiated on the 22nd December 2023) and the clinical trial batch (Lot Number P126404A; stability study initiated on the 30th September 2024). Both batches of SPVX02 were manufactured under GMP conditions and are subsequently analysed for 21 predefined quality attributes at each stability study timepoint. Testing timepoints include: 1 month at 40°C and 75% relative humidity (RH) with 3 freeze thaw cycles, 3, 6, 9, 12, 18, 24, 36 and 48 months at 30°C and 75% relative humidity, and 3 and 6 months at 40°C and 75% relative humidity.

This section presents the in-vivo potency challenge study data generated to date, with data generated up to the 18 month timepoint for the SPVX02 engineering batch and up to the 12 month timepoint for the SPVX02 clinical batch.

In-vivo potency testing is performed according to the compendial method required by the European Pharmacopoeia for the assessment vaccine potency and is routinely used for batch release and stability evaluation of tetanus diphtheria vaccines.

**Methods:**

**Diphtheria:** European Pharmacopoeia (Ph. Eur. 2.7.6).
Healthy guinea pigs (250–350 g) were immunised subcutaneously with serial dilutions of the test vaccine or a reference preparation calibrated in International Units (IU). After 28 days, animals were challenged subcutaneously with diphtheria toxin at a dose corresponding to approximately 100 $LD_{50}$ per animal. Survival was recorded 4 days post-challenge. Vaccine potency was calculated relative to the reference preparation based on the proportion of surviving animals in each group, using parallel-line statistical analysis. Assays were considered valid only if confidence limits, dose–response linearity and challenge dose requirements specified by the Pharmacopoeia were met.

**Tetanus:** European Pharmacopoeia (Ph. Eur. 2.7.8).
Animals were immunised subcutaneously with graded dilutions of the test vaccine or a reference vaccine calibrated in International Units. After a 28-day immunisation period, animals were challenged subcutaneously with tetanus toxin at a dose equivalent to approximately 50 × $PD_{50}$ (or $LD_{50}$ where permitted). Animals were monitored daily, and survival or absence of tetanus paralysis was recorded 4–5 days after challenge. Vaccine potency was calculated relative to the reference preparation using standard parallel-line statistical methods. Assays were considered valid only if confidence limits, dose–response linearity and challenge dose requirements specified by the Pharmacopoeia were met.

**Results:**

Complete protection against supra lethal doses of active tetanus and diphtheria toxins in immunised guinea-pigs was shown with SPVX02 engineering batch (Lot Number P126402E) following initial batch release, after storage for 1 month at 40°C and 75% relative humidity with 3 freeze thaw cycles, after storage at 30°C and 75% relative humidity for 3, 6, 9, 12 and 18 months (see Figure S1 and Table S2), and after storage at 40°C and 75% relative humidity for 3 and 6 months (see also Table S2). These studies support the hypothesis that the reformulation of Tetadif bulk vaccine to produce SPVX02, and the prolonged storage at high temperatures, do not affect the potency of the SPVX02 vaccine.

Comparable potency results were obtained for the clinical trial material batch (Lot Number P126404A) at batch release, after storage for 1 month at 40°C and 75% relative humidity with 3 freeze thaw cycles, after storage at 30°C and 75% relative humidity for 3, 6, 9, 12 months and storage for 3 and 6 months at 40°C and 75% relative humidity (data not shown).

**Figure S1: In-Vivo Potency of Tetanus and Diphtheria Toxoids in SPVX02 Engineering Batch (Lot Number P126402E).**

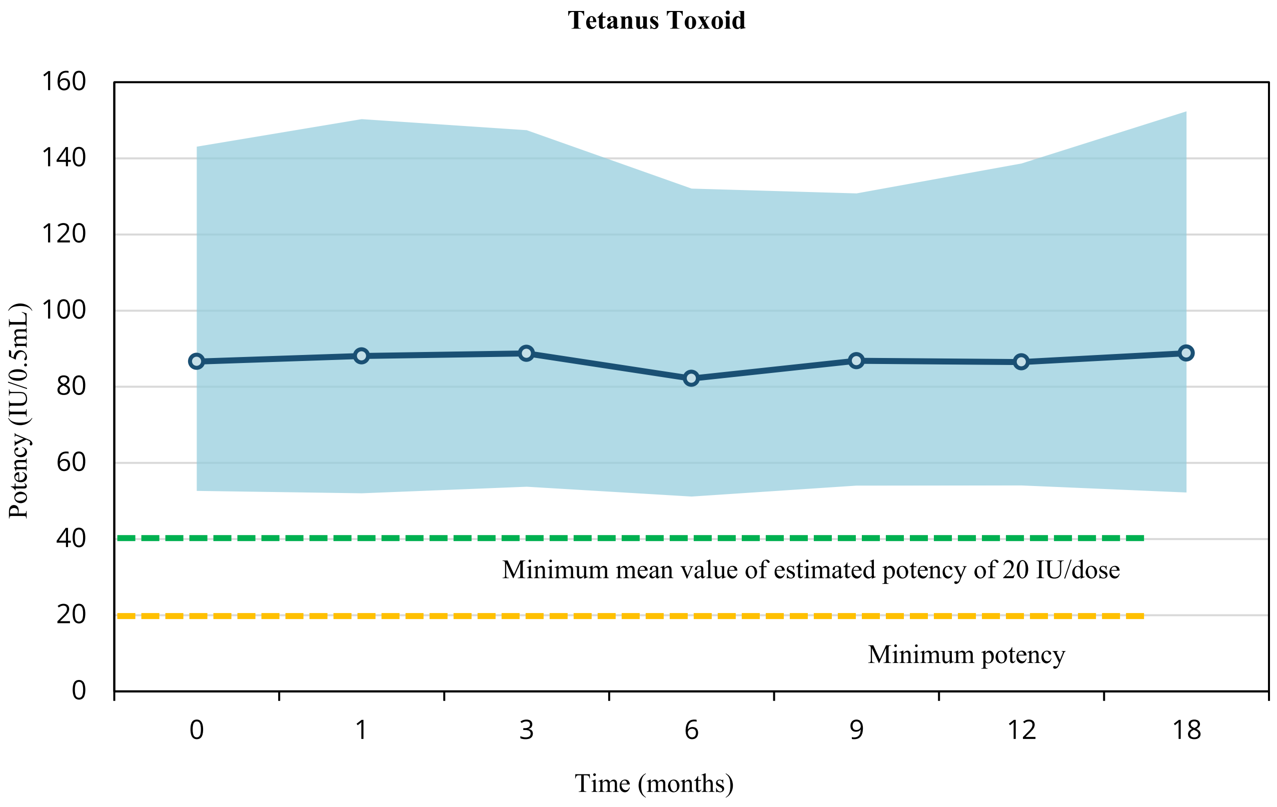

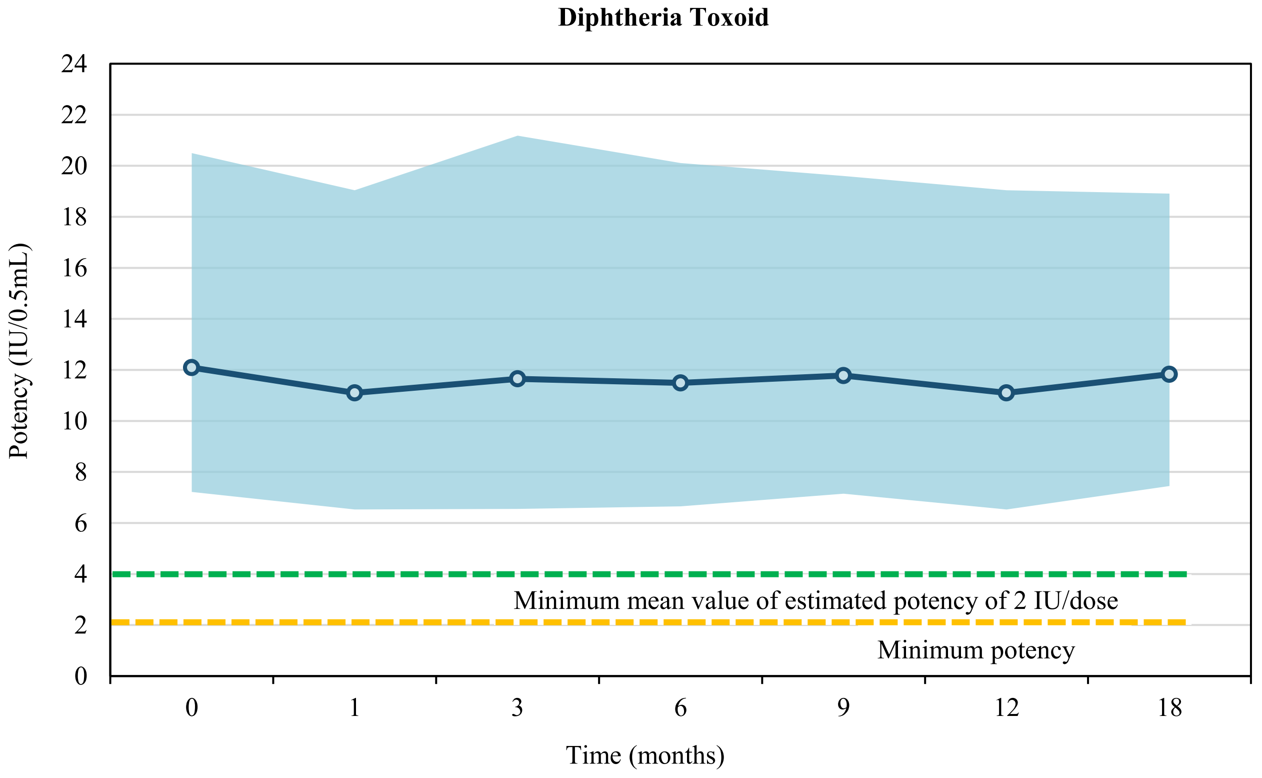

**Table S2: In-Vivo Potency (Lethal Challenge Test) of Tetanus and Diphtheria toxoids in SPVX02 Engineering Batch (Lot Number P126402E)**

| TETANUS TOXOID CHALLENGE TEST | | | | | | | | | |
|---|---|---|---|---|---|---|---|---|---|
| | **T=0 (batch release)** | **T=1 month[a] (freeze-thaw)** | **T=3 months at 30±2°C[b]** | **T=3 months at 40±2°C[c]** | **T=6 months at 30±2°C[b]** | **T=6 months at 40±2°C[c]** | **T=9 months at 30±2°C[b]** | **T=12 months at 30±2°C[b]** | **T=18 months at 30±2°C[b]** |
| **N° Units Administered** | **N° Died/N° Animals Challenged** | | | | | | | | |
| 16 IU | 0/8 | 0/8 | 0/8 | 0/8 | 0/8 | 0/8 | 0/8 | 0/8 | 0/8 |
| 8 IU | 3/8 | 3/8 | 2/8 | 1/8 | 2/8 | 2/8 | 2/8 | 2/8 | 3/8 |
| 4 IU | 4/8 | 4/8 | 5/8 | 6/8 | 6/8 | 6/8 | 5/8 | 4/8 | 5/8 |
| 2 IU | 8/8 | 7/8 | 7/8 | 7/8 | 7/8 | 7/8 | 8/8 | 8/8 | 7/8 |
| | **Potency (95% CL) (IU/0.5 mL)** | | | | | | | | |
| | **86.63** (52.67-143.05) | **88.07** (52.02-150.27) | **88.75** (53.76-147.37) | **89.80** (55.71-145.30) | **82.17** (51.17-132.03) | **82.17** (51.17-132.03) | **86.82** (54.05-130.77) | **86.50** (54.08-138.65) | **88.81** (52.22-152.34) |
| **DIPHTHERIA TOXOID CHALLENGE TEST** | | | | | | | | | |
| | **T=0 (batch release)** | **T=1 month[a] (freeze-thaw)** | **T=3 months at 30±2°C[b]** | **T=3 months at 40±2°C[c]** | **T=6 months at 30±2°C[b]** | **T=6 months at 40±2°C[c]** | **T=9 months at 30±2°C[b]** | **T=12 months at 30±2°C[b]** | **T=18 months at 30±2°C[b]** |
| **N° Units Administered** | **N° Died/N° Animals Challenged** | | | | | | | | |
| 40 IU | 0/8 | 0/8 | 1/8 | 0/8 | 1/8 | 0/8 | 0/8 | 0/8 | 0/8 |
| 20 IU | 3/8 | 3/8 | 2/8 | 2/8 | 2/8 | 3/8 | 2/8 | 3/8 | 1/8 |
| 10 IU | 5/8 | 5/8 | 3/8 | 5/8 | 3/8 | 5/8 | 3/8 | 5/8 | 3/8 |
| 5 IU | 7/8 | 7/8 | 7/8 | 7/8 | 8/8 | 7/8 | 8/8 | 7/8 | 8/8 |
| | **Potency (95% CL) (IU/0.5 mL)** | | | | | | | | |
| | **12.09** (7.22-20.50) | **11.10** (6.53-19.04) | **11.65** (6.55-21.18) | **11.09** (6.72-18.42) | **11.49** (6.65-20.11) | **11.10** (6.53-19.04) | **11.78** (7.15-19.60) | **11.10** (6.53-19.04) | **11.83** (7.45-18.91) |

[a]Data from freeze-thaw stability testing of SPVX02 stored at 21 days at 40±2°C / 75±5% RH then (-20±2°C for 24 hours: 40±2°C / 75±5% RH for 24 hours) x3.
[b]Data from long-term stability testing of SPVX02 stored at 30±2°C 75±5%RH.
[c]Data from accelerated stability testing of SPVX02 stored at 40±2°C 75±5% RH.
Abbreviations: CL=confidence limit; IU=international units; N°=Number; RH=relative humidity; T=time.

**01/2008:20708**
**corrected 11.0**

### 2.7.8. ASSAY OF TETANUS VACCINE (ADSORBED)

The potency of tetanus vaccine is determined by administration of the vaccine to animals (guinea-pigs or mice) followed either by challenge with tetanus toxin (method A or B) or by determination of the titre of antibodies against tetanus toxoid in the serum of the guinea-pigs (method C). In both cases, the potency of the vaccine is calculated by comparison with a reference vaccine, calibrated in International Units. For methods A and B, in countries where the paralysis method is not obligatory, the $LD_{50}$ method may be used. For the $LD_{50}$ method, the number of animals and the procedure are identical to those described for the paralysis method, but the end-point is the death of the animal rather than paralysis.

The International Unit is the activity contained in a stated amount of the International Standard for tetanus toxoid (adsorbed). The equivalence in International Units of the International Standard is stated by the World Health Organization.

*Tetanus vaccine (adsorbed) BRP* is calibrated in International Units with reference to the International Standard.

The method chosen for the assay of tetanus vaccine (adsorbed) depends on the intended purpose. Method A or B is used:

1. during development of a vaccine, to assay batches produced to validate the production;
2. wherever revalidation is needed following a significant change in the manufacturing process.

Method A or B may also be used for the routine assay of batches of vaccine, but in the interests of animal welfare, method C is used wherever possible.

Method C may be used, except as specified under 1 and 2 above, after verification of the suitability of the method for the product. For this purpose, a suitable number of batches (usually 3) are assayed by method C and method A or B. Where different vaccines (monovalent or combinations) are prepared from tetanus toxoid of the same origin and with comparable levels (expressed in Lf/mL) of the same tetanus toxoid, suitability demonstrated for the combination with the highest number of components can be assumed to be valid for combinations with fewer components and for monovalent vaccines. Any combinations containing a whole-cell pertussis component or containing haemophilus type b conjugate vaccine with tetanus toxoid in the same vial must always be assessed separately.

For combinations containing diphtheria and tetanus components, the serological assay (method C) can be performed with the same group of animals used for the serological assay of the diphtheria vaccine (adsorbed) (*2.7.6*) when the common immunisation conditions for the tetanus and the diphtheria components (for example, doses, duration) have been demonstrated to be valid for the combined vaccine.

The design of the assays described below uses multiple dilutions for the test and reference preparations. Based on the potency data obtained in multiple-dilution assays, it may be possible to reduce the number of animals needed to obtain a statistically significant result by applying a simplified model such as a single dilution for both test and reference preparations. Such a model enables the analyst to determine whether the potency of the test preparation is significantly higher than the minimum required, but does not give information on the dose-response curves and their linearity, parallelism and significant slope. The simplified model allows for a considerable reduction in the number of animals required and must be considered by each analyst in accordance with the provisions of the European Convention for the Protection of Vertebrate Animals Used for Experimental and Other Scientific Purposes.

Where a single-dilution assay is used, production and test consistency over time are monitored via suitable indicators and by carrying out a full multiple-dilution assay periodically, for example every 2 years. For serological assays, suitable indicators to monitor test consistency are:

- the mean and standard deviation of relative antitoxin titres or scores of the serum samples obtained after administration of a fixed dose of the vaccine reference preparation;
- the antitoxin titres or scores of run controls (positive and negative serum samples);
- the ratio of antitoxin titres or scores for the positive serum control to the serum samples corresponding to the reference vaccine.

#### METHOD A. CHALLENGE TEST IN GUINEA-PIGS

*SELECTION AND DISTRIBUTION OF THE TEST ANIMALS*

Use in the test healthy guinea-pigs from the same stock, each weighing 250-350 g. Use guinea-pigs of the same sex or with males and females equally distributed between the groups. Distribute the guinea-pigs in not fewer than 6 equal groups; use groups containing a number of animals sufficient to obtain results that fulfil the requirements for a valid assay prescribed below. If the activity of the challenge toxin has to be determined, include 3 further groups of 5 guinea-pigs as unvaccinated controls.

*SELECTION OF THE CHALLENGE TOXIN*

Select a preparation of tetanus toxin containing not less than 50 times the 50 per cent paralytic dose per millilitre. If the challenge toxin preparation has been shown to be stable, it is not necessary to verify the paralytic dose for every assay.

*PREPARATION OF THE CHALLENGE TOXIN SOLUTION*

Immediately before use, dilute the challenge toxin with a suitable diluent (for example, peptone buffered saline solution pH 7.4) to obtain a stable challenge toxin solution containing approximately 50 times the 50 per cent paralytic dose per millilitre. If necessary, use portions of the challenge toxin solution diluted 1 to 16, 1 to 50 and 1 to 160 with the same diluent to determine the activity of the toxin.

*DILUTION OF THE TEST AND REFERENCE PREPARATIONS*

Using a 9 g/L solution of *sodium chloride R*, prepare dilutions of the vaccine to be examined and of the reference preparation, such that for each, the dilutions form a series differing by not more than 2.5-fold steps and in which the intermediate dilutions, when injected subcutaneously at a dose of 1.0 mL per guinea-pig, protect approximately 50 per cent of the animals from the paralytic effects of the subcutaneous injection of the quantity of tetanus toxin prescribed for this test.

*IMMUNISATION AND CHALLENGE*

Allocate the dilutions, 1 to each of the groups of guinea-pigs, and inject subcutaneously 1.0 mL of each dilution into each guinea-pig in the group to which that dilution is allocated. After 28 days, inject subcutaneously into each animal 1.0 mL of the challenge toxin solution (containing 50 times the 50 per cent paralytic dose).

*DETERMINATION OF THE ACTIVITY OF THE CHALLENGE TOXIN*

If necessary, allocate the 3 dilutions made from the challenge toxin solution, 1 to each of the 3 groups of 5 guinea-pigs, and inject subcutaneously 1.0 mL of each solution into each guinea-pig in the group to which that solution is allocated. The activity and stability of the challenge toxin are determined by carrying out a suitable number of determinations of the 50 per cent paralytic dose. It is then not necessary to repeat the determination for each assay.

*READING AND INTERPRETATION OF RESULTS*

Examine the guinea-pigs twice daily. Remove and euthanise all animals showing definite signs of tetanus paralysis. Count the number of guinea-pigs without paralysis 5 days after injection of the challenge toxin. Calculate the potency of the vaccine to be examined relative to the potency of the reference preparation on the basis of the proportion of challenged animals without paralysis in each group of vaccinated guinea-pigs, using the usual statistical methods (for example, *5.3*).

*REQUIREMENTS FOR A VALID ASSAY*

The test is not valid unless:

- for both the vaccine to be examined and the reference preparation, the 50 per cent protective dose lies between the largest and smallest doses of the preparations given to the guinea-pigs;
- where applicable, the number of paralysed animals in the 3 groups of 5 injected with the dilutions of the challenge toxin solution indicates that the challenge was approximately 50 times the 50 per cent paralytic dose;
- the confidence limits ($P = 0.95$) are not less than 50 per cent and not more than 200 per cent of the estimated potency;
- the statistical analysis shows a significant slope and no deviation from linearity and parallelism of the dose-response curves (chapter *5.3* describes possible alternatives if significant deviations are observed).

The test may be repeated but when more than 1 test is performed the results of all valid tests must be combined in the estimate of potency.

METHOD B. CHALLENGE TEST IN MICE

*SELECTION AND DISTRIBUTION OF THE TEST ANIMALS*

Use in the test healthy mice from the same stock, about 5 weeks old and from a strain shown to be suitable. Use mice of the same sex or with males and females equally distributed between the groups. Distribute the mice in not fewer than 6 equal groups; use groups containing a number of animals sufficient to obtain results that fulfil the requirements for a valid assay prescribed below. If the challenge toxin to be used has not been shown to be stable or has not been adequately standardised, include 3 further groups of not fewer than 5 mice to serve as unvaccinated controls.

*SELECTION OF THE CHALLENGE TOXIN*

Select a preparation of tetanus toxin containing not less than 100 times the 50 per cent paralytic dose per millilitre. If the challenge toxin preparation has been shown to be stable, it is not necessary to verify the paralytic dose for every assay.

*PREPARATION OF THE CHALLENGE TOXIN SOLUTION*

Immediately before use, dilute the challenge toxin with a suitable diluent (for example, peptone buffered saline solution pH 7.4) to obtain a stable challenge toxin solution containing approximately 50 times the 50 per cent paralytic dose in 0.5 mL. If necessary, use portions of the challenge toxin solution diluted 1 to 16, 1 to 50 and 1 to 160 with the same diluent to determine the activity of the toxin.

*DILUTION OF THE TEST AND REFERENCE PREPARATIONS*

Using a 9 g/L solution of *sodium chloride R*, prepare dilutions of the vaccine to be examined and of the reference preparation, such that for each, the dilutions form a series differing by not more than 2.5-fold steps and in which the intermediate dilutions, when injected subcutaneously at a dose of 0.5 mL per mouse, protect approximately 50 per cent of the animals from the paralytic effects of the subcutaneous injection of the quantity of tetanus toxin prescribed for this test.

*IMMUNISATION AND CHALLENGE*

Allocate the dilutions, 1 to each of the groups of mice, and inject subcutaneously 0.5 mL of each dilution into each mouse in the group to which that dilution is allocated. After 28 days, inject subcutaneously into each animal 0.5 mL of the challenge toxin solution (containing 50 times the 50 per cent paralytic dose).

*DETERMINATION OF THE ACTIVITY OF THE CHALLENGE TOXIN*

If necessary, allocate the 3 dilutions made from the challenge toxin solution, 1 to each of the 3 groups of not fewer than 5 mice, and inject subcutaneously 0.5 mL of each solution into each mouse in the group to which that solution is allocated.

*READING AND INTERPRETATION OF RESULTS*

Examine the mice twice daily. Remove and euthanise all animals showing definite signs of tetanus paralysis. Count the number of mice without paralysis 4 days after injection of the challenge toxin. Calculate the potency of the vaccine to be examined relative to the potency of the reference preparation on the basis of the proportion of challenged animals without paralysis in each group of vaccinated mice, using the usual statistical methods (for example, *5.3*).

*REQUIREMENTS FOR A VALID ASSAY*

The test is not valid unless:

- for both the vaccine to be examined and the reference preparation, the 50 per cent protective dose lies between the largest and smallest doses of the preparations given to the mice;
- where applicable, the number of paralysed animals in the 3 groups of not fewer than 5 injected with the dilutions of the challenge toxin solution, indicates that the challenge dose was approximately 50 times the 50 per cent paralytic dose;
- the confidence limits ($P = 0.95$) are not less than 50 per cent and not more than 200 per cent of the estimated potency;
- the statistical analysis shows a significant slope and no deviation from linearity and parallelism of the dose-response curves (chapter *5.3* describes possible alternatives if significant deviations are observed).

The test may be repeated but when more than 1 test is performed the results of all valid tests must be combined in the estimate of potency.

METHOD C. DETERMINATION OF ANTIBODIES IN GUINEA-PIGS

*SELECTION AND DISTRIBUTION OF THE TEST ANIMALS*

Use in the test healthy guinea-pigs from the same stock, each weighing 250-350 g. Use guinea-pigs of the same sex or with males and females equally distributed between the groups. Distribute the guinea-pigs in not fewer than 6 equal groups; use groups containing a number of animals sufficient to obtain results that fulfil the requirements for a valid assay prescribed below. Use a further group of non-vaccinated guinea-pigs of the same origin to provide a negative serum control. If test consistency has been demonstrated, a reference negative serum control may be used.

*REFERENCE PREPARATION*

Use a suitable reference preparation such as *tetanus vaccine (adsorbed) BRP* or a batch of vaccine shown to be effective in clinical studies, or a batch representative thereof, and which has been calibrated in International Units with reference to *tetanus vaccine (adsorbed) BRP* or the International Standard for tetanus toxoid (adsorbed).

*DILUTION OF THE TEST AND REFERENCE PREPARATIONS*

Using a 9 g/L solution of *sodium chloride R* as diluent, prepare serial dilutions of the vaccine to be examined and the reference preparation; series differing by 2.5- to 5-fold steps have been found to be suitable. Use not fewer than 3 dilutions within the range of, for example, 0.5-16 IU/mL for each series. Use the dilutions for immunisation preferably within 1 h of preparation. Allocate 1 dilution to each group of guinea-pigs.

*IMMUNISATION*

Inject subcutaneously to each guinea-pig 1.0 mL of the dilution allocated to its group.

*BLOOD SAMPLING*

35-42 days after immunisation, take a blood sample from each vaccinated and control guinea-pig using a suitable method.

*PREPARATION OF SERUM SAMPLES*

Avoid frequent freezing and thawing of serum samples. To avoid microbial contamination, it is preferable to carry out manipulations in a laminar-flow cabinet.

*DETERMINATION OF ANTIBODY TITRE*

Determine the relative antibody titre or score of each serum sample by a suitable immunochemical method (*2.7.1*). The methods shown below (enzyme-linked immunosorbent assay (ELISA) and toxin-binding inhibition (ToBI)) have been found to be suitable.

*CALCULATION OF POTENCY*

Calculate the potency of the vaccine to be examined in International Units relative to the reference preparation, using the usual statistical methods (for example, *5.3*).

*REQUIREMENTS FOR A VALID ASSAY*

The test is not valid unless:

- the confidence limits ($P$ = 0.95) are not less than 50 per cent and not more than 200 per cent of the estimated potency;
- the statistical analysis shows a significant slope and no deviation from linearity and parallelism of the dose-response curves (chapter *5.3* describes possible alternatives if significant deviations are observed).

The test may be repeated but when more than 1 test is performed the results of all valid tests must be combined in the estimate of potency.

*The following section is published for information.*

#### Assay of tetanus vaccine (adsorbed): guidelines

METHOD A. CHALLENGE TEST IN GUINEA-PIGS

*READING AND INTERPRETATION OF RESULTS*

In order to minimise suffering in the test animals, it is recommended to note the degree of paralysis on a scale such as that shown below. The scale gives typical signs when subcutaneous injection of the challenge toxin is made mid-ventrally, directly behind the sternum with the needle pointing towards the neck of the guinea-pig. Grade T3 is taken as the end-point, but with experience grade T2 can be used instead. Tetanus toxin produces in at least 1 of the forelimbs paralysis that can be recognised at an early stage. The tetanus grades in guinea-pigs are characterised by the following signs:

- T1: slight stiffness of 1 forelimb, but difficult to observe;
- T2: paresis of 1 forelimb which still can function;
- T3: paralysis of 1 forelimb. The animal moves reluctantly, the body is often slightly banana-shaped owing to scoliosis;
- T4: the forelimb is completely stiff and the toes are immovable. The muscular contraction of the forelimb is very pronounced and usually scoliosis is observed;
- T5: tetanus seizures, continuous tonic spasm of muscles;
- D: death.

METHOD B. CHALLENGE TEST IN MICE

*READING AND INTERPRETATION OF RESULTS*

In order to minimise suffering in the test animals, it is recommended to note the degree of paralysis on a scale such as that shown below. The scale gives typical signs when injection of the challenge toxin is made in the dorsal region, close to one of the hind legs. Grade T3 is taken as the end-point, but with experience grade T2 can be used instead. Tetanus toxin produces in the toxin-injected hind leg paresis followed by paralysis that can be recognised at an early stage. The tetanus grades in mice are characterised by the following signs:

- T1: slight stiffness of toxin-injected hind leg, only observed when the mouse is lifted by the tail;
- T2: paresis of the toxin-injected hind leg, which still can function for walking;
- T3: paralysis of the toxin-injected hind leg, which does not function for walking;
- T4: the toxin-injected hind leg is completely stiff with immovable toes;
- T5: tetanus seizures, continuous tonic spasm of muscles;
- D: death.

METHOD C. DETERMINATION OF ANTIBODIES IN GUINEA-PIGS

*PREPARATION OF SERUM SAMPLES*

For the preparation of serum samples, the following technique has been found to be suitable. Invert the tubes containing blood samples 6 times and allow to stand at 37 °C for 2 h, then at 4 °C for 2 h. Centrifuge at room temperature at 800 *g* for 20 min. Transfer the serum to sterile tubes and store at a temperature below – 20 °C. At least a 40 per cent yield of serum is obtained by this procedure.

*DETERMINATION OF ANTIBODY TITRE*

The ELISA and ToBI tests shown below are given as examples of immunochemical methods that have been found to be suitable for the determination of antibody titre.

**Determination of antibody titre in guinea-pig serum by enzyme-linked immunosorbent assay (ELISA)**. Dilutions of test and reference sera are made on ELISA plates coated with tetanus toxoid. A positive guinea-pig serum control and a negative guinea-pig serum control are included on each plate to monitor the assay performance. Peroxidase-conjugated rabbit or goat antibody directed against guinea-pig-IgG is added, followed by a peroxidase substrate. Optical density is measured and the relative antibody titre is calculated using the usual statistical methods (for example, *5.3*).

*Reagents and equipment*

- *ELISA plates*: 96 wells, columns 1-12, rows A-H.
- *Clostridium tetani guinea-pig antiserum (for vaccines-human use) BRP* (positive control serum).
- *Peroxidase conjugate.* Peroxidase-conjugated rabbit or goat antibody directed against guinea-pig IgG.
- *Tetanus toxoid.*
- *Carbonate coating buffer pH 9.6.* Dissolve 1.59 g of *anhydrous sodium carbonate R* and 2.93 g of *sodium hydrogen carbonate R* in 1000 mL of *water R*. Distribute into 150 mL bottles and sterilise by autoclaving at 121 °C for 15 min.
- *Phosphate-buffered saline pH 7.4 (PBS).* Dissolve with stirring 80.0 g of *sodium chloride R*, 2.0 g of *potassium dihydrogen phosphate R*, 14.3 g of *disodium hydrogen phosphate dihydrate R* and 2.0 g of *potassium chloride R* in 1000 mL of *water R*. Store at room temperature to prevent crystallisation. Dilute to 10 times its volume with *water R* before use.
- *Citric acid solution.* Dissolve 10.51 g of *citric acid monohydrate R* in 1000 mL of *water R* and adjust the solution to pH 4.0 with a 400 g/L solution of *sodium hydroxide R*.
- *Washing buffer.* PBS containing 0.5 g/L of *polysorbate 20 R*.
- *Diluent block buffer.* PBS containing 0.5 g/L of *polysorbate 20 R* and 25 g/L of dried skimmed milk.

– *Peroxidase substrate.* Shortly before use, dissolve 10 mg of *diammonium 2,2′-azinobis(3-ethylbenzothiazoline-6-sulfonate) R* (ABTS) in 20 mL of citric acid solution. Immediately before use add 5 μL of *strong hydrogen peroxide solution R*.

*Method*

The description below is given as an example of a suitable plate layout but others may be used. Wells 1A-H are for negative control serum and wells 2A-H and 12A-H are for positive control serum for assay monitoring. Wells 3-11A-H are for test samples.

Coat each well of the ELISA plates with 100 μL of tetanus toxoid solution (0.5 Lf/mL in carbonate coating buffer pH 9.6). Allow to stand overnight at 4 °C in a humid atmosphere. To avoid temperature gradient effects, do not stack more than 4 plates high. On the following day, wash the plates thoroughly with washing buffer. Block the plates by addition of 100 μL of diluent block buffer to each well. Incubate in a humid atmosphere at 37 °C for 1 h. Wash the plates thoroughly with washing buffer. Place 100 μL of diluent block buffer in each well of the plates, except those of row A. Prepare suitable dilutions of negative control serum, positive control serum (from about 0.01 IU/mL) and test sera. Allocate the negative control serum to column 1, positive control serum to columns 2 and 12 and test sera to columns 3-11 and add 100 μL of each serum to the first 2 wells of the column to which it is allocated. Using a multichannel micropipette, make twofold serial dilutions from row B down the plate to row H, by transferring 100 μL from one well to the next. Discard 100 μL from the last row so that all wells contain 100 μL. Incubate at 37 °C for 2 h. Wash thoroughly with washing buffer. Prepare a suitable dilution (a 2000-fold dilution has been found to be suitable) of peroxidase conjugate in diluent block buffer and add 100 μL to each well. Incubate at 37 °C in a humid atmosphere for 1 h. Wash the plates thoroughly with washing buffer. Add 100 μL of peroxidase substrate to each well. Allow to stand at room temperature, protected from light, for 30 min. Read the plates at 405 nm in the same order as addition of substrate was made.

**Determination of antibody titre in guinea-pig serum by toxin- or toxoid-binding inhibition (ToBI)**. Tetanus toxin or toxoid is added to serial dilutions of test and reference sera; the serum/antigen mixtures are incubated overnight. To determine unbound toxin or toxoid, the mixtures are transferred to an ELISA plate coated with tetanus antitoxin. Peroxidase-conjugated equine anti-tetanus IgG is added followed by a peroxidase substrate. Optical density is measured and the antibody titre is calculated using the usual statistical methods (for example, *5.3*). A positive control serum and a negative control serum are included on each plate to monitor assay performance.

*Reagents and equipment*

– *Round-bottomed, rigid polystyrene microplates.*

– *Flat-bottomed ELISA plates.*

– *Tetanus toxin or tetanus toxoid.*

– *Clostridium tetani guinea-pig antiserum (for vaccines-human use) BRP* (positive control serum).

– *Equine anti-tetanus IgG.*

– *Peroxidase-conjugated equine anti-tetanus IgG.*

– *Carbonate buffer pH 9.6.* Dissolve 1.5 g of *anhydrous sodium carbonate R*, 2.39 g of *sodium hydrogen carbonate R* and 0.2 g of *sodium azide R* in 1000 mL of *water R*, adjust to pH 9.6 and autoclave at 121 °C for 20 min.

– *Sodium acetate buffer pH 5.5.* Dissolve 90.2 g of *anhydrous sodium acetate R* in 900 mL of *water R*, adjust to pH 5.5 using a saturated solution of *citric acid monohydrate R* and dilute to 1000 mL with *water R*.

– *Phosphate-buffered saline pH 7.2 (PBS).* Dissolve 135.0 g of *sodium chloride R*, 20.55 g of *disodium hydrogen phosphate dihydrate R* and 4.80 g of *sodium dihydrogen phosphate monohydrate R* in *water R* and dilute to 15 L with the same solvent. Autoclave at 100 °C for 60 min.

– *Diluent buffer.* PBS containing 5 g/L of *bovine albumin R* and 0.5 g/L of *polysorbate 80 R*.

– *Block buffer.* PBS containing 5 g/L of *bovine albumin R*.

– *Tetramethylbenzidine solution.* 6 g/L solution of *tetramethylbenzidine R* in *ethanol (96 per cent) R*. The substance dissolves within 30-40 min at room temperature.

– *Peroxidase substrate.* Mix 90 mL of *water R*, 10 mL of sodium acetate buffer pH 5.5, 1.67 mL of tetramethylbenzidine solution and 20 μL of *strong hydrogen peroxide solution R*.

– *Washing solution.* Tap water containing 0.5 g/L of *polysorbate 80 R*.

*Method*

Block the microplates by placing in each well 150 μL of block buffer. Cover the plates with a lid or sealer. Incubate in a humid atmosphere at 37 °C for 1 h. Wash the plates thoroughly with washing solution. Place 100 μL of PBS in each well. Place 100 μL of reference guinea-pig tetanus antitoxin in the first well of a row. Place 100 μL of undiluted test sera in the first well of the required number of rows. Using a multichannel micropipette, make twofold serial dilutions across the plate (up to column 10), by transferring 100 μL from one well to the next. Discard 100 μL from the last column so that all wells contain 100 μL. Prepare a 0.1 Lf/mL solution of tetanus toxin or toxoid using PBS as diluent. Add 40 μL of this solution to each well except those of column 12. The wells of column 11 are a positive control. Add 40 μL of PBS to the wells of column 12 (negative control). Shake the plates gently and cover them with lids. Coat the ELISA plates: immediately before use make a suitable dilution of equine anti-tetanus IgG in carbonate buffer pH 9.6 and add 100 μL to each well. Incubate the 2 series of plates overnight in a humid atmosphere at 37 °C. To avoid temperature gradient effects, do not stack more than 4 plates high. Cover the plates with lids. On the following day, wash the ELISA plates thoroughly with washing solution. Block the plates by placing in each well 125 μL of block buffer. Incubate at 37 °C in a humid atmosphere for 1 h. Wash the plates thoroughly with washing solution. Transfer 100 μL of the pre-incubation mixture from the polystyrene plates to the corresponding wells of the ELISA plates, starting with column 12 and then continuing from 1 to 11. Cover the plates with a lid. Incubate at 37 °C in a humid atmosphere for 2 h. Wash the ELISA plates thoroughly with washing solution. Make a suitable dilution (a 4000-fold dilution has been found to be suitable) of the peroxidase-conjugated equine anti-tetanus IgG in diluent buffer. Add 100 μL of the dilution to each well and cover the plates with a lid. Incubate at 37 °C in a humid atmosphere for 1.5 h. Wash the ELISA plates thoroughly with washing solution. Add 100 μL of peroxidase substrate to each well. A blue colour develops. Incubate the plates at room temperature. Stop the reaction at a given time (within 10 min) by the addition of 100 μL of a 2 M solution of sulfuric acid prepared from *sulfuric acid R* to each well in the same order as the addition of substrate. The colour changes from blue to yellow. Measure the absorbance at 450 nm immediately after addition of the sulfuric acid or maintain the plates in the dark until reading.

Appropriate hydrolysis times are usually between 3 min and 15 min, but deviations are permissible if better linearity of the dose-response relationship is thus obtained.

Calculate the potency of the test preparation by the usual statistical methods (for example, *5.3*).

**04/2017:20705**

### 2.7.5. ASSAY OF HEPARIN

The anticoagulant activity of heparin is determined *in vitro* by its ability to accelerate the inhibition of thrombin, factor IIa (anti-IIa assay), by antithrombin. The International Unit is the activity contained in a stated amount of the International Standard for unfractionated heparin. *Heparin sodium BRP*, calibrated in International Units by comparison with the International Standard using the 2 assays given below, is used as the reference preparation.

The assay of anti-factor Xa activity is carried out to determine the ratio of anti-factor Xa activity to anti-factor IIa activity.

For anti-IIa and anti-Xa assays, carry out the assay by determining the absorbance (end-point method) or the change of absorbance per minute (kinetic method).

ANTI-FACTOR IIa ACTIVITY

**Reference and test solutions**

Prepare 4 independent series of 4 dilutions each of the substance to be examined and of *heparin sodium BRP* in *tris(hydroxymethyl)aminomethane-EDTA buffer solution pH 8.4 R1*; a concentration range within 0.005 IU and 0.03 IU per millilitre is suitable. The dilutions chosen must give a linear response when results are plotted as absorbance against log concentration.

**Procedure**

Label 16 tubes for the dilutions of the substance to be examined and 16 tubes for the dilutions of the reference preparation: $T_1$, $T_2$, $T_3$, $T_4$ for each of the 4 series of dilutions of the substance to be examined and $S_1$, $S_2$, $S_3$, $S_4$ for each of the 4 series of dilutions of the reference preparation. To each of the 32 tubes add 100 µL of *antithrombin III solution R5* and 50 µL of the appropriate dilution of the substance to be examined or the reference preparation. After each addition, mix but do not allow bubbles to form. Treating the tubes in 2 subsequent series in the order $S_1$, $S_2$, $S_3$, $S_4$, $T_1$, $T_2$, $T_3$, $T_4$, $T_1$, $T_2$, $T_3$, $T_4$, $S_1$, $S_2$, $S_3$, $S_4$, allow to equilibrate at 37 °C (water-bath or heating block) for at least 1 min and add to each tube 25 µL of *human thrombin solution R2*. Incubate for exactly 1 min and add 50 µL of a chromogenic substrate specific to factor IIa at a concentration suitable for the assay (for example, D-phenylalanyl-L-pipecolyl-L-arginine-4-nitroanilide dihydrochloride dissolved in *water R* to give a 1.25 mM solution).

For the kinetic method, transfer the mixtures to semi-micro cuvettes and measure the change in absorbance per minute (*2.2.25*) at 405 nm using a suitable reading device.

For the end-point method, stop the reaction after exactly 4 min by adding 50 µL of a 20 per cent *V/V* solution of *glacial acetic acid R*. Assess whether exactly 4 min of incubation with the chromogenic substrate yields the optimal absorbance reading and, if necessary, adjust the incubation time to give the best dose-response curve. Then, transfer the mixtures to semi-micro cuvettes and measure the absorbance (*2.2.25*) at 405 nm using a suitable reading device.

Determine the blank amidolytic activity at the beginning and at the end of the procedure in a similar manner, using *tris(hydroxymethyl)aminomethane-EDTA buffer solution pH 8.4 R1* instead of the reference and test solutions; the 2 blank values do not differ significantly.

Calculate the regression of the absorbance on log concentrations of the solutions of the substance to be examined and of *heparin sodium BRP*, and calculate the potency of the substance to be examined in International Units per millilitre using the usual statistical methods for parallel-line assays (*5.3*).

ANTI-FACTOR Xa ACTIVITY

**Reference and test solutions**

Prepare 4 independent series of 4 dilutions each of the substance to be examined and of *heparin sodium BRP* in *tris(hydroxymethyl)aminomethane-EDTA buffer solution pH 8.4 R1*; a concentration range within 0.03 IU and 0.375 IU per millilitre is suitable. The dilutions chosen must give a linear response when results are plotted as absorbance against log concentration.

**Procedure**

Label 16 tubes for the dilutions of the substance to be examined and 16 tubes for the dilutions of the reference preparation: $T_1$, $T_2$, $T_3$, $T_4$ for each of the 4 series of dilutions of the substance to be examined and $S_1$, $S_2$, $S_3$, $S_4$ for each of the 4 series of dilutions of the reference preparation. To each of the 32 tubes add 50 µL of *antithrombin III solution R6* and 50 µL of the appropriate dilution of the substance to be examined or the reference preparation. After each addition, mix but do not allow bubbles to form. Treating the tubes in 2 subsequent series in the order $S_1$, $S_2$, $S_3$, $S_4$, $T_1$, $T_2$, $T_3$, $T_4$, $T_1$, $T_2$, $T_3$, $T_4$, $S_1$, $S_2$, $S_3$, $S_4$, allow to equilibrate at 37 °C (water-bath or heating block) for 1 min and add to each tube 100 µL of *bovine factor Xa solution R2*. Incubate for exactly 2 min and add 100 µL of a chromogenic substrate specific to factor Xa at a concentration suitable for the assay (for example, *N*-α-benzyloxycarbonyl-D-arginyl-L-glycyl-L-arginine-4-nitroanilide dihydrochloride dissolved in *water R* to give a 1 mM solution).

For the kinetic method, transfer the mixtures to semi-micro cuvettes and measure the change in absorbance per minute (*2.2.25*) at 405 nm using a suitable reading device.

For the end-point method, stop the reaction after exactly 4 min by adding 50 µL of a 20 per cent *V/V* solution of *glacial acetic acid R*. Assess whether exactly 4 min of incubation with the chromogenic substrate yields the optimal absorbance reading and, if necessary, adjust the incubation time to give the best dose-response curve. Then, transfer the mixtures to semi-micro cuvettes and measure the absorbance (*2.2.25*) at 405 nm using a suitable reading device.

Determine the blank amidolytic activity at the beginning and at the end of the procedure in a similar manner, using *tris(hydroxymethyl)aminomethane-EDTA buffer solution pH 8.4 R1* instead of the reference and test solutions; the 2 blank values do not differ significantly.

Calculate the regression of the absorbance on log concentrations of the solutions of the substance to be examined and of *heparin sodium BRP*, and calculate the potency of the substance to be examined in International Units per millilitre using the usual statistical methods for parallel-line assays (*5.3*).

**01/2008:20706**
**corrected 6.0**

### 2.7.6. ASSAY OF DIPHTHERIA VACCINE (ADSORBED)

The potency of diphtheria vaccine is determined by administration of the vaccine to guinea-pigs followed either by challenge with diphtheria toxin (method A or B) or by determination of the titre of antibodies against diphtheria toxin or toxoid in the serum of guinea-pigs (method C). In both cases, the potency of the vaccine is calculated by comparison with a reference preparation, calibrated in International Units.

The International Unit is the activity contained in a stated amount of the International Standard, which consists of a quantity of diphtheria toxoid adsorbed on aluminium hydroxide. The equivalence in International Units of the International Standard is stated by the World Health Organization (WHO).

*Diphtheria vaccine (adsorbed) BRP* is suitable for use as a reference preparation.

The method chosen for the assay of diphtheria vaccine (adsorbed) depends on the intended purpose. Method A or B is used:

1. during development of a vaccine, to assay batches produced to validate the production;
2. wherever revalidation is needed following a significant change in the manufacturing process.

Method A or B may also be used for the routine assay of batches of vaccine, but in the interests of animal welfare, method C is used wherever possible.

Method C may be used, except as specified under 1 and 2 above, after verification of the suitability of the method for the product. For this purpose, a suitable number of batches (usually 3) are assayed by method C and method A or B. Where different vaccines (monovalent or combinations) are prepared from diphtheria toxoid of the same origin, and with comparable levels (expressed in Lf/mL) of the same diphtheria toxoid, suitability demonstrated for the combination with the highest number of components can be assumed to be valid for combinations with fewer components and for monovalent vaccines. Any combinations containing a whole-cell pertussis component or containing haemophilus type b conjugate vaccine with diphtheria toxoid or CRM 197 diphtheria protein as carrier in the same vial must always be assessed separately.

For combinations containing diphtheria and tetanus components, the serological assay (method C) can be performed with the same group of animals used for the serological assay of the tetanus vaccine (adsorbed) (*2.7.8*) when the common immunisation conditions for the diphtheria and the tetanus components (for example, doses, duration) have been demonstrated to be valid for the combined vaccine.

The design of the assays described below uses multiple dilutions for the test and reference preparations. Once the analyst has sufficient experience with this method for a given vaccine, it is possible to apply a simplified model such as a single dilution for both test and reference preparations. Such a model enables the analyst to determine whether the potency of the test preparation is significantly higher than the minimum required, but does not give information on linearity, parallelism and the dose-response curve. The simplified model allows for a considerable reduction in the number of animals required and must be considered by each analyst in accordance with the provisions of the European Convention for the Protection of Vertebrate Animals Used for Experimental and Other Scientific Purposes.

Where a single-dilution assay is used, production and test consistency over time are monitored via suitable indicators and by carrying out a full multiple-dilution assay periodically, for example every 2 years. For serological assays, suitable indicators to monitor test consistency are:

- the mean and standard deviation of relative antitoxin titres or scores of the serum samples obtained after administration of a fixed dose of the vaccine reference preparation;
- the antitoxin titres or scores of run controls (positive and negative serum samples);
- the ratio of antitoxin titres or scores for the positive serum control to the serum samples corresponding to the reference vaccine.

#### METHOD A: INTRADERMAL CHALLENGE TEST IN GUINEA-PIGS

*SELECTION AND DISTRIBUTION OF THE TEST ANIMALS*

Use in the test healthy, white guinea-pigs from the same stock and of a size suitable for the prescribed number of challenge sites, the difference in body mass between the heaviest and the lightest animal being not greater than 100 g. Use guinea-pigs of the same sex or with males and females equally distributed between the groups. Distribute the guinea-pigs in not fewer than 6 equal groups; use groups containing a number of animals sufficient to obtain results that fulfil the requirements for a valid assay prescribed below. If the challenge toxin to be used has not been shown to be stable or has not been adequately standardised, include 5 guinea-pigs as unvaccinated controls.

*SELECTION OF THE CHALLENGE TOXIN*

Select a preparation of diphtheria toxin containing 67 to 133 lr/100 in 1 Lf and 25 000 to 50 000 minimal reacting doses for guinea-pig skin in 1 Lf. If the challenge toxin preparation has been shown to be stable, it is not necessary to verify the activity for every assay.

*PREPARATION OF THE CHALLENGE TOXIN SOLUTION*

Immediately before use, dilute the challenge toxin with a suitable diluent to obtain a challenge toxin solution containing about 0.0512 Lf in 0.2 mL. Prepare from this a further series of 5 four-fold dilutions containing about 0.0128, 0.0032, 0.0008, 0.0002 and 0.00005 Lf in 0.2 mL.

*DILUTION OF THE TEST AND REFERENCE PREPARATIONS*

Using a 9 g/L solution of *sodium chloride R*, prepare dilutions of the vaccine to be examined and of the reference preparation, such that for each, the dilutions form a series differing by not more than 2.5-fold steps and in which the intermediate dilutions, when injected subcutaneously at a dose of 1.0 mL per guinea-pig, will result in an intradermal score of approximately 3 when the animals are challenged.

*IMMUNISATION AND CHALLENGE*

Allocate the dilutions, 1 to each of the groups of guinea-pigs, and inject subcutaneously 1.0 mL of each dilution into each guinea-pig in the group to which that dilution is allocated. After 28 days, shave both flanks of each guinea-pig and inject 0.2 mL of each of the 6 toxin dilutions intradermally into 6 separate sites on each of the vaccinated guinea-pigs in such a way as to minimise interference between adjacent sites.

*DETERMINATION OF THE ACTIVITY OF THE CHALLENGE TOXIN*

If necessary, inject the unvaccinated control animals with dilutions containing 80, 40, 20, 10 and $5 \times 10^{-6}$ Lf of the challenge toxin.

*READING AND INTERPRETATION OF RESULTS*

Examine all injection sites 48 h after injection of the challenge toxin and record the incidence of specific diphtheria erythema. Record also the number of sites free from such reactions as the intra-dermal challenge score. Tabulate the intradermal challenge scores for all the animals receiving the same dilution of vaccine and use those data with a suitable transformation, such as $(\text{score})^2$ or arcsin $((\text{score}/6)^2)$, to obtain an estimate of the relative potency for each of the test preparations by parallel-line quantitative analysis.

*REQUIREMENTS FOR A VALID ASSAY*

The test is not valid unless:

- for both the vaccine to be examined and the reference preparation, the mean score obtained at the lowest dose level is less than 3 and the mean score at the highest dose level is more than 3;

– where applicable, the toxin dilution that contains $40 \times 10^{-6}$ Lf gives a positive erythema in at least 80 per cent of the control guinea-pigs and the dilution containing $20 \times 10^{-6}$ Lf gives a positive erythema in less than 80 per cent of the guinea-pigs (if these criteria are not met a different toxin has to be selected);

– the confidence limits ($P$ = 0.95) are not less than 50 per cent and not more than 200 per cent of the estimated potency;

– the statistical analysis shows no deviation from linearity and parallelism.

The test may be repeated but when more than 1 test is performed the results of all valid tests must be combined in the estimate of potency.

#### METHOD B: LETHAL CHALLENGE TEST IN GUINEA-PIGS

*SELECTION AND DISTRIBUTION OF THE TEST ANIMALS*

Use in the test healthy guinea-pigs from the same stock, each weighing 250-350 g. Use guinea-pigs of the same sex or with males and females equally distributed between the groups. Distribute the guinea-pigs in not fewer than 6 equal groups; use groups containing a number of animals sufficient to obtain results that fulfil the requirements for a valid assay prescribed below. If the challenge toxin to be used has not been shown to be stable or has not been adequately standardised, include 4 further groups of 5 guinea-pigs as unvaccinated controls.

*SELECTION OF THE CHALLENGE TOXIN*

Select a preparation of diphtheria toxin containing not less than 100 $LD_{50}$ per millilitre. If the challenge toxin preparation has been shown to be stable, it is not necessary to verify the lethal dose for every assay.

*PREPARATION OF THE CHALLENGE TOXIN SOLUTION*

Immediately before use, dilute the challenge toxin with a suitable diluent to obtain a challenge toxin solution containing approximately 100 $LD_{50}$ per millilitre. If necessary, use portions of the challenge toxin solution diluted 1 to 32, 1 to 100 and 1 to 320 with the same diluent.

*DILUTION OF THE TEST AND REFERENCE PREPARATIONS*

Using a 9 g/L solution of *sodium chloride R*, prepare dilutions of the vaccine to be examined and of the reference preparation, such that for each, the dilutions form a series differing by not more than 2.5-fold steps and in which the intermediate dilutions, when injected subcutaneously at a dose of 1.0 mL per guinea-pig, protect approximately 50 per cent of the animals from the lethal effects of the subcutaneous injection of the quantity of diphtheria toxin prescribed for this test.

*IMMUNISATION AND CHALLENGE*

Allocate the dilutions, 1 to each of the groups of guinea-pigs, and inject subcutaneously 1.0 mL of each dilution into each guinea-pig in the group to which that dilution is allocated. After 28 days, inject subcutaneously into each animal 1.0 mL of the challenge toxin solution (100 $LD_{50}$).

*DETERMINATION OF THE ACTIVITY OF THE CHALLENGE TOXIN*

If necessary, allocate the challenge toxin solution and the 3 dilutions made from it, 1 to each of the 4 groups of 5 guinea-pigs, and inject subcutaneously 1.0 mL of each solution into each guinea-pig in the group to which that solution is allocated.

*READING AND INTERPRETATION OF RESULTS*

Count the number of surviving guinea-pigs 4 days after injection of the challenge toxin. Calculate the potency of the vaccine to be examined relative to the potency of the reference preparation on the basis of the proportion of animals surviving in each of the groups of vaccinated guinea-pigs, using the usual statistical methods (for example, *5.3*).

*REQUIREMENTS FOR A VALID ASSAY*

The test is not valid unless:

– for both the vaccine to be examined and the reference preparation, the 50 per cent protective dose lies between the largest and smallest doses of the preparations given to the guinea-pigs;

– where applicable, the number of animals that die in the 4 groups of 5 injected with the challenge toxin solution and its 3 dilutions indicates that the challenge dose was approximately 100 $LD_{50}$;

– the confidence limits ($P$ = 0.95) are not less than 50 per cent and not more than 200 per cent of the estimated potency;

– the statistical analysis shows no deviation from linearity and parallelism.

The test may be repeated but when more than 1 test is performed the results of all valid tests must be combined in the estimate of potency.

#### METHOD C. DETERMINATION OF ANTIBODIES IN GUINEA-PIGS

*SELECTION AND DISTRIBUTION OF THE TEST ANIMALS*

Use in the test healthy guinea-pigs from the same stock, each weighing 250-350 g. Use guinea-pigs of the same sex or with males and females equally distributed between the groups. Distribute the guinea-pigs in not fewer than 6 equal groups; use groups containing a number of animals sufficient to obtain results that fulfil the requirements for a valid assay prescribed below. Use a further group of non-vaccinated guinea-pigs of the same origin to provide a negative serum control. If test consistency has been demonstrated, a reference negative serum control may be used.

*REFERENCE PREPARATION*

Use a suitable reference preparation such as *diphtheria vaccine (adsorbed) BRP* or a batch of vaccine shown to be effective in clinical studies, or a batch representative thereof, and which has been calibrated in International Units with reference to *diphtheria vaccine (adsorbed) BRP* or the International Standard for diphtheria toxoid (adsorbed).

*DILUTION OF THE TEST AND REFERENCE PREPARATIONS*

Using a 9 g/L solution of *sodium chloride R* as diluent, prepare serial dilutions of the vaccine to be examined and the reference preparation; series differing by 2.5- to 5-fold steps have been found to be suitable. Use not fewer than 3 dilutions within the range of, for example, 0.5-16 IU/mL for the reference vaccine and within the range of, for example, 1:2 to 1:125 for the vaccine to be examined. Use the dilutions for immunisation preferably within 1 h of preparation. Allocate 1 dilution to each group of guinea-pigs.

*IMMUNISATION*

Inject subcutaneously to each guinea-pig 1.0 mL of the dilution allocated to its group.

*BLOOD SAMPLING*

35-42 days after immunisation, take a blood sample from each vaccinated and control guinea-pig using a suitable method.

*PREPARATION OF SERUM SAMPLES*

Avoid frequent freezing and thawing of serum samples. To avoid microbial contamination, it is preferable to carry out manipulations in a laminar-flow cabinet.

*DETERMINATION OF ANTIBODY TITRE*

Determine the relative antibody titre or score of each serum sample by a suitable immunochemical method (*2.7.1*). The methods shown below (enzyme-linked immunosorbent assay (ELISA) and Vero cell assay) have been found to be suitable.

*CALCULATION OF POTENCY*

Calculate the potency of the vaccine to be examined in International Units relative to the reference preparation, using the usual statistical methods (for example, *5.3*).

*REQUIREMENTS FOR A VALID ASSAY*

The test is not valid unless:

- the confidence limits ($P$ = 0.95) are not less than 50 per cent and not more than 200 per cent of the estimated potency;
- the statistical analysis shows a significant slope and no deviation from linearity and parallelism of the dose-response curves (chapter *5.3* describes possible alternatives if significant deviations are observed).

The test may be repeated but when more than 1 test is performed the results of all valid tests must be combined in the estimate of potency.

*The following section is published for information.*

#### Assay of diphtheria vaccine (adsorbed): guidelines

METHOD C. DETERMINATION OF ANTIBODIES IN GUINEA-PIGS

*PREPARATION OF SERUM SAMPLES*

For the preparation of serum samples, the following technique has been found to be suitable. Invert the tubes containing blood samples 6 times and allow to stand at 37 °C for 2 h, then at 4 °C for 2 h. Centrifuge at room temperature at 800 *g* for 20 min. Transfer the serum to sterile tubes and store at a temperature below – 20 °C. At least a 40 per cent yield of serum is obtained by this procedure.

*DETERMINATION OF ANTIBODY TITRE*

The ELISA and Vero cell assays shown below are given as examples of immunochemical methods that have been found to be suitable for the determination of antibody titre.

**Determination of antibody titre in guinea-pig serum by enzyme-linked immunosorbent assay (ELISA)**. Dilutions of test and reference sera are made on ELISA plates coated with diphtheria toxoid. A positive guinea-pig serum control and a negative guinea-pig serum control are included on each plate to monitor the assay performance. Peroxidase-conjugated rabbit or goat antibody directed against guinea-pig-IgG is added, followed by a peroxidase substrate. Optical density is measured and the relative antibody titre is calculated using the usual statistical methods (for example, *5.3*).

*Reagents and equipment*

- *ELISA plates*: 96 wells, columns 1-12, rows A-H.
- *Diphtheria guinea-pig antiserum (for vaccines-human use)* (positive control serum), obtained by immunisation of guinea-pigs using *diphtheria vaccine (adsorbed) BRP.*
- *Peroxidase conjugate.* Peroxidase-conjugated rabbit or goat antibody directed against guinea-pig IgG.
- *Diphtheria toxoid.*
- *Carbonate coating buffer pH 9.6.* Dissolve 1.59 g of *anhydrous sodium carbonate R* and 2.93 g of *sodium hydrogen carbonate R* in 1000 mL of *water R.* Distribute into 150 mL bottles and sterilise by autoclaving at 121 °C for 15 min.
- *Phosphate-buffered saline pH 7.4 (PBS).* Dissolve with stirring 80.0 g of *sodium chloride R*, 2.0 g of *potassium dihydrogen phosphate R*, 14.3 g of *disodium hydrogen phosphate dihydrate R* and 2.0 g of *potassium chloride R* in 1000 mL of *water R.* Store at room temperature to prevent crystallisation. Dilute to 10 times its volume with *water R* before use.
- *Citric acid solution.* Dissolve 10.51 g of *citric acid monohydrate R* in 1000 mL of *water R* and adjust the solution to pH 4.0 with a 400 g/L solution of *sodium hydroxide R.*
- *Washing buffer.* PBS containing 0.5 g/L of *polysorbate 20 R.*
- *Diluent blocking buffer.* PBS containing 0.5 g/L of *polysorbate 20 R* and 25 g/L of dried skimmed milk.
- *Peroxidase substrate.* Shortly before use, dissolve 10 mg of *diammonium 2,2′-azinobis(3-ethylbenzothiazoline-6-sulfonate) R* (ABTS) in 20 mL of citric acid solution. Immediately before use add 5 µL of *strong hydrogen peroxide solution R.*

*Method*

The description below is given as an example of a suitable plate layout but others may be used. Wells 1A-H are for negative control serum and wells 2A-H and 12A-H are for positive control serum for assay monitoring. Wells 3-11A-H are for test samples.

Coat each well of the ELISA plates with 100 µL of diphtheria toxoid solution (0.5 Lf/mL in carbonate coating buffer pH 9.6). Allow to stand overnight at 4 °C in a humid atmosphere. To avoid temperature gradient effects, do not stack more than 4 plates high. On the following day, wash the plates thoroughly with washing buffer. Block the plates by addition of 100 µL of diluent block buffer to each well. Incubate in a humid atmosphere at 37 °C for 1 h. Wash the plates thoroughly with washing buffer. Place 100 µL of diluent block buffer in each well of the plates, except those of row A. Prepare suitable dilutions of negative control serum, positive control serum (from about 0.01 IU/mL) and test sera. Allocate the negative control serum to column 1, positive control serum to columns 2 and 12 and test sera to columns 3-11 and add 100 µL of each serum to the first 2 wells of the column to which it is allocated. Using a multichannel micropipette, make twofold serial dilutions from row B, down the plate to row H, by transferring 100 µL from one well to the next well. Discard 100 µL from the last row so that all wells contain 100 µL. Incubate at 37 °C for 2 h. Wash thoroughly with washing buffer. Prepare a suitable dilution (a 2000-fold dilution has been found to be suitable) of peroxidase conjugate in diluent block buffer and add 100 µL to each well. Incubate at 37 °C in a humid atmosphere for 1 h. Wash the plates thoroughly with washing buffer. Add 100 µL of peroxidase substrate to each well. Allow to stand at room temperature, protected from light, for 30 min. Read the plates at 405 nm in the same order as addition of substrate was made.

**Determination of antibody titre in guinea-pig serum by Vero cell assay**. The method used relies either on metabolic inhibition (method 1) or on cytotoxicity (method 2) as the end point, and on either microscopic (cell morphology) or visual (colour) inspection of the cells.

The limit of detection is specific for each antitoxin and is usually between 0.015 IU/mL (method 1) and 0.05 IU/mL (method 2).

The endpoint is taken as the highest serum dilution protecting cells from the diphtheria toxin effect. The antitoxin activity is calculated with respect to guinea-pig or WHO reference standard, and expressed in International Units per millilitre.

*Reagents and equipment*

- *Flat-bottomed tissue culture plates*: 96 wells, columns 1-12, rows A-H.
- *75 cm$^2$ tissue culture flasks.*
- *Diphtheria toxin.*
- *Diphtheria guinea-pig antiserum (for vaccines-human use)* (positive control serum), obtained by immunisation of guinea-pigs with *diphtheria vaccine (adsorbed) BRP.*
- *Vero cells* (African Green Monkey kidney cells). Cell passages from P2 to P15 are suitable for use.

*Method 1.* The diphtheria toxin causes a cytopathogenic effect on Vero cells leading to cellular lysis. Antibodies directed against diphtheria toxin may inhibit this cytopathogenic effect. Consequently, the potency of a diphtheria vaccine may be indirectly determined with the help of this cell culture system if different serum dilutions from immunised animals are cultured with a constant toxin concentration. In the Vero cell assay, yellow colour indicates viable cells, red colour dead cells. When only part of the cells are dead, the colour may be orange.

*Reagents and equipment*

- *Modified MEM.* Minimum Essential Medium (MEM) with Earle's Salts, without L-glutamine and sodium bicarbonate.
- *Modified medium 199.* Medium 199, with Hanks' Solution and L-glutamine, without sodium bicarbonate.
- *Foetal bovine serum.*
- *Sodium bicarbonate 7.5 per cent solution.*
- *Trypsin solution:* trypsin 2.5 per cent solution.
- *EDTA solution:* EDTA 0.02 per cent (Versene 1:5000) solution.
- *Modified D-PBS.* Dulbecco's phosphate buffered saline (D-PBS), without calcium, or magnesium.
- *L-glutamine 200mM solution.*
- *Penicillin/streptomycin solution.*
- *Primary culture medium.* To 50 mL of modified MEM add 440 mL of *water R*, 5 mL of L-glutamine 200 mM solution, and 10 mL of sodium bicarbonate 7.5 per cent solution. To 25 mL of this medium add 1.25 mL of foetal bovine serum.
- *Maintenance culture medium.* Similar to the primary culture medium except that 0.5 mL instead of 1.25 mL of foetal bovine serum is added to 20 mL of the enriched MEM medium.
- *Medium A.* To 50.0 mL of modified medium 199 add 440.0 mL of *water R*, 5.0 mL of L-glutamine 200 mM solution and 10.0 mL of sodium bicarbonate 7.5 per cent solution.
- *Medium B.* To 150.0 mL of medium A add 3.0 mL of foetal bovine serum and 0.3 mL of penicillin/streptomycin solution.
- *Medium C.* To 22.0 mL of medium A add 0.44 mL of foetal bovine serum and 0.44 mL of penicillin/streptomycin solution.

Vero cells are cultured in tissue culture flasks (for example 75 $cm^2$/250 mL) in an incubator at 36 ± 1 °C, 5 per cent $CO_2$ and 90 per cent relative humidity. Vero cells are first grown in the primary culture medium. After 2-3 days of growth, the primary culture medium is replaced by the maintenance culture medium. When a confluent monolayer is obtained, the culture supernatant is discarded and the cell layer washed gently with modified D-PBS. Add a mixture of 1 volume of trypsin solution and 1 volume of EDTA solution to the flask. Swirl the flask gently and incubate in the $CO_2$ incubator for about 3 min until the cells start to break from the monolayer. Vigorously tap the side of the flask to make the cells fall. Resuspend the cells in 5-6 mL of fresh medium C to obtain a homogeneous suspension. Prepare a cell suspension in medium C containing approximately $1 \times 10^5$ cells/mL.

Place 25 μL of medium B in each well except those of column 1. Place 25 μL of the diphtheria guinea-pig antiserum (for vaccines-human use) (positive control serum, working dilution in medium B of 0.40 IU/mL) in wells A1, A2 and A11. Place 25 μL of guinea-pig serum samples in wells B-G of columns 1, 2 and 11. Place 25 μL of negative control serum in row H of columns 1, 2 and 11. Using a multichannel micropipette, make twofold serial dilutions across the plate (from column 2 up to column 10 for rows A-G and up to column 8 for row H). Discard 25 μL from the wells in column 10 in rows A-G, and from well H8.

Reconstitute the diphtheria toxin with saline solution to give a solution of 50 IU/mL. Prepare a 50-fold dilution of this diphtheria toxin dilution in medium B to obtain a working solution of 1.0 IU/mL. Add 25 μL of this working solution to wells A12 and B12 (toxin control). Make twofold serial dilutions by tranferring 25 μL from one well to the next, from well B12 down to H12. Change the tip between each dilution. Discard 25 μL from well H12. Add 25 μL of medium B to wells B12-H12. Then, place 25 μL of the working dilution of the diphtheria toxin (1.0 IU/mL) in each well of rows A-H, from column 1-10, except in wells H9 and H10 (cells only, without serum and without toxin).

Cover the plates with lids or sealer and shake gently. Incubate the plates for at least 2 h in a humid container in a $CO_2$ incubator at 37 °C. Add 200 μL of cell suspension containing 1 x $10^5$ cells/mL to all the wells. Cover the plates with sealer. Incubate at 37 °C for 5 days. Check for microbial contamination by microscopic examination.

Yellow wells are recorded as negative and red wells indicate dead cells and are recorded as positive. A colour between yellow and red indicates a mixture of viable and dead cells and is recorded as positive/negative. The results based on the change in colour can be confirmed by reading viable and dead cells under the microscope.

The potency of the guinea-pig antiserum samples is obtained by comparing the last well of the standard preparation showing complete neutralisation of the toxin, with the last well of the sample demonstrating the same effect. For calculations of potency, it must be remembered that the endpoint may be between a negative well and a positive/negative well.

*Method 2*: Thiazolyl blue MTT is reduced to a blue/black formazan product by the mitochondrial dehydrogenase of viable cells, and thus serves as a quantitative measure of living cells present, indicating when the toxin has been neutralised by the antitoxin. White or colourless wells indicate absence of viable cells due to insufficient antitoxin to neutralise the toxin.

*Reagents and equipment*

- *MEM* (Minimal Essential Media).
- *Newborn calf serum.*
- *Antibiotic solution* (containing 10 000 units of penicillin, 10 mg of streptomycin and 25 μg of amphotericin B per millilitre).
- *L-glutamine 200mM solution.*
- *Trypsin-EDTA.*
- *Thiazolyl blue MTT* [3-(4,5-dimethylthiazol-2-yl)-2,5-diphenyltetrazolium bromide].
- *1 M HEPES buffer pH 8.1.* Dissolve 18.75 g of HEPES in 82.5 mL of *water R* and 30.0 mL of *2 M sodium hydroxide R*.
- *Glucose solution (10 per cent).*
- *Complete culture medium.* Mix 200 mL of MEM with 10 mL of newborn calf serum, 3.0 mL of 1 M HEPES buffer pH 8.1, 2.0 mL of glucose solution (10 per cent), 2.0 mL of antibiotic solution and 2.0 mL of L-glutamine 200mM solution.
- *Phosphate-buffered saline pH 7.4 (PBS).* Dissolve 10.0 g of *sodium chloride R*, 0.75 g of *potassium chloride R*, 1.44 g of *disodium hydrogen phosphate dodecahydrate R*, and 0.125 g of *potassium dihydrogen phosphate R* in *water R*, and dilute to 1000.0 mL with the same solvent. Adjust the pH if necessary. Autoclave at 120 °C for 15 min.
- *Thiazolyl blue MTT solution.* Dissolve 0.1 g of thiazolyl blue MTT in 20 mL of PBS. Sterilise by filtration (0.2 μm) and store in dark bottle.
- *pH adjuster solution.* Mix 40 mL of *acetic acid R* with 1.25 mL of *1 M hydrochloric acid* and 8.75 mL of *water R*.
- *Extraction buffer pH 4.7.* Dissolve 10 g of *sodium laurilsulfate R* in *water R* and add 50 mL of *dimethylformamide R*, and dilute to 100 mL with *water R*. Adjust the pH with an appropriate volume of pH adjuster solution.

Vero cells are cultured in tissue culture flasks (for example 75 $cm^2$/250 mL) in an incubator at 36 ± 1 °C, 5 per cent $CO_2$ and 90 per cent relative humidity. Vero cells are grown in the complete culture medium. After 6-7 days of growth, a confluent monolayer is obtained, the culture supernatant is discarded and the cell layer is washed 3 times with trypsin-EDTA: gently pipette out the medium, add 0.5-1 mL of trypsin-EDTA, swirl the flask and tip the trypsin out. Do this twice, and the 3rd time, place the flask in the incubator for 5 min until the cells start to break from the monolayer. Vigorously tap the side of the flask to make the cells fall. Resuspend the cells in 6-25 mL of fresh complete culture medium to obtain a homogeneous suspension. Prepare a cell suspension in complete culture medium containing approximately $4 \times 10^5$ cells/mL.

Place 50 µL of complete culture medium in each well except those of column 1. Place 100 µL of diphtheria guinea pig antiserum (for vaccines-human use) (positive control serum, working dilution in complete culture medium of 0.12 IU/mL) in well A1 and 50 µL in well A11. Place 100 µL of guinea pig test serum samples, diluted if necessary, in wells B1-G1. Add 50 µL of the same sample to wells B11-G11 in the corresponding row. Place 100 µL of negative control serum in well H1 and 50 µL in well H11. Using a multi-channel micropipette, make twofold serial dilutions by transferring 50 µL from one well to the next working across the plate (from column 1-10 for rows A-G and from column 1-8 for row H).

Diphtheria toxin of known activity and Lf content is diluted to a suitable working stock containing at least 4 minimum cytopathic doses in complete culture medium. Add 50 µL of the diluted toxin to each well except H9 and H10 (cell control), A11-H11 (serum control) and A12-H12 (toxin control). Add 100 µL of diluted toxin to well A12 and make twofold serial dilutions by transferring 50 µL from one well to the next working down the plate (from well A12-H12). Discard 50 µL from well H12. Add 50 µL of complete medium to wells H9 and H10.

Cover the plates with a lid or sealer and leave for 1 h at room temperature to allow toxin neutralisation to occur. 50 µL of cell suspension containing approximately $4 \times 10^5$ cells/mL is added to each well. The plates are sealed and incubated at 37 °C for 6 days. Check for microbial contamination by microscopic examination. 10 µL of thyazolyl blue MTT solution is added to each well. The plates are incubated at 37 °C for a further 2-4 h. Then, the medium is removed and 100 µL of extraction buffer pH 4.7 is added to each well. The plates are incubated at 37 °C and left overnight to aid the extraction process. Once extraction and solubilisation is complete, plates are visually examined or read at 570 nm.

Blue/black wells are recorded as negative (all the cells are alive, toxin neutralisation by antitoxin) and white or colourless wells indicate dead cells (no toxin neutralisation) and are recorded as positive.

The potency of the test antitoxin is obtained by comparing the last well of the reference antitoxin preparation showing neutralisation of the toxin, with the last well of the antitoxin preparation demonstrating the same effect. The neutralising antibody titre of the sample being examined can be calculated by multiplication of the dilution factor with total number of International Units per millilitre of the reference preparation at the end point. The test is valid if all the cells in the toxin control are dead and reference antitoxin gives a neutralisation in at least the first 2 dilutions tested.

**07/2011:20707**

#### 2.7.7. ASSAY OF PERTUSSIS VACCINE (WHOLE CELL)

The potency of pertussis vaccine (whole cell) is determined by comparing the dose necessary to protect mice against the effects of a lethal dose of *Bordetella pertussis*, administered intracerebrally, with the quantity of a reference preparation, calibrated in International Units, needed to give the same protection.

The International Unit is the activity contained in a stated amount of the International Standard which consists of a quantity of dried pertussis vaccine. The equivalence in International Units of the International Standard is stated by the World Health Organization.

**Selection and distribution of the test animals**. Use in the test healthy mice less than 5 weeks old of a suitable strain from the same stock, the difference in mass between the heaviest and the lightest being not greater than 5 g. Distribute the mice in 6 groups of not fewer than 16 and 4 groups of 10. The mice must all be of the same sex or the males and females distributed equally between the groups.

**Selection of the challenge strain and preparation of the challenge suspension**. Select a suitable strain of *B. pertussis* capable of causing the death of mice within 14 days of intracerebral injection. If more than 20 per cent of the mice die within 48 h of the injection the strain is not suitable. Make one subculture from the strain and suspend the harvested *B. pertussis* in a solution containing 10 g/L of *casein R* hydrolysate and 6 g/L of *sodium chloride R* and having a pH of 7.0 to 7.2 or in another suitable solution. Determine the opacity of the suspension. Prepare a series of dilutions in the same solution and allocate each dilution to a group of 10 mice. Inject intracerebrally into each mouse a dose (0.02 mL or 0.03 mL) of the dilution allocated to its group. After 14 days, count the number of mice surviving in each group. From the results, calculate the expected opacity of a suspension containing 100 $LD_{50}$ in each challenge dose. For the test of the vaccine to be examined make a fresh subculture from the same strain of *B. pertussis* and prepare a suspension of the harvested organisms with an opacity corresponding to about 100 $LD_{50}$ in each challenge dose. Prepare 3 dilutions of the challenge suspension.

**Determination of potency**. Prepare 3 serial dilutions of the vaccine to be examined and 3 similar dilutions of the reference preparation such that in each the intermediate dilution may be expected to protect about 50 per cent of the mice from the lethal effects of the challenge dose of *B. pertussis*. Suggested doses are 1/8, 1/40 and 1/200 of the human dose of the vaccine to be examined and 0.5 IU, 0.1 IU and 0.02 IU of the reference preparation, each dose being contained in a volume not exceeding 0.5 mL. Allocate the 6 dilutions, one to each of the groups of not fewer than 16 mice, and inject intraperitoneally into each mouse one dose of the dilution allocated to its group. After 14 - 17 days inject intracerebrally into each animal in the groups of not fewer than 16, one dose of the challenge suspension. Allocate the challenge suspension and the 3 dilutions made from it, one to each of the groups of 10 mice, and inject intracerebrally one dose of each suspension into each mouse in the group to which that suspension is allocated. Exclude from consideration any mice that die within 48 h of challenge. Count the number of mice surviving in each of the groups after 14 days. Calculate the potency of the vaccine to be examined relative to the potency of the reference preparation on the basis of the numbers of animals surviving in each of the groups of not fewer than 16.

The test is not valid unless:

- for both the vaccine to be examined and the reference preparation, the 50 per cent protective dose lies between the largest and the smallest doses given to the mice;
- the number of animals that die in the 4 groups of 10 injected with the challenge suspension and its dilutions indicates that the challenge dose is approximately 100 $LD_{50}$; and
- the statistical analysis shows no deviation from linearity or parallelism.

The test may be repeated but when more than one test is performed the results of all valid tests must be combined.

**БУЛ БИО – НЦЗПБ ЕАД** **BB – NCIPD EAD**

1504 София, бул. Янко Сакъзов 26
Факс: ** 359 2 943 34 55
Телефон Изпълнителен директор:
** 359 2 944 61 91


1504 Sofia, 26 Yanko Sakazov blvd.
Fax: **359 2 943 34 55
Telephone Executive Director:
**359 2 944 61 91


### CERTIFICATE OF ANALYSIS

**SPVX02 CTM**

Tetanus-Diphtheria (adsorbed) (with reduced dose of diphtheria for adults)

**Lot № P126404A**

Date of manufacture: August 2024

| № | Test/Method | Acceptance Criteria | Result |
|---|---|---|---|
| 1. | Appearance | Suspension of white or grey particles dispersed in colourless or pale-yellow liquids and may form a sediment at the bottom of the container. Free of visible particles. | Pass |
| 2. | pH | 6.0 - 7.0 | 6.51 |
| 3. | Aluminium content | Maximum 1.25 mg/0.5 mL | 0.60 |
| 4. | Sodium Chloride content | 7.0 - 10.0 g/L | 9.07 |
| 5. | Identity of diphtheria | Identity complies | Pass |
| 6. | Identity of tetanus | Identity complies | Pass |
| 7. | Sterility | Sterile | Pass |
| 8. | Residual free formaldehyde | Maximum 0.2 g/L | Less than 0.2 |
| 9. | Degree of adsorption for tetanus | Not less than 80 % | 100 |
| 10. | Degree of adsorption for diphtheria | Not less than 80 % | 100 |
| 11. | Specific toxicity test for tetanus | None of the animals show abnormal reaction | Pass |
| 12. | Specific toxicity test for diphtheria | None of the animals show abnormal reaction | Pass |
| 13. | Potency test for tetanus component | Not less than 20 IU/0.5 mL | 86.50 (54.08-138.65) |
| 14. | Potency test for diphtheria component | Not less than 2 IU/0.5 mL | 11.76 (7.42-18.69) |

**Conclusion:**

**SPVX02 CTM, Lot № P126404A** has been successfully tested according to a list of standard qualified assays with defined acceptance limits (per Annex 1 of the Service Agreement between BB-NCIPD and Stablepharma, signed on 13th of June 2023).

Date of issue: 04.12.2024

Head of Quality Control Department:
R. Alexiev, PhD

Confidential level 1

patheon
by Thermo Fisher Scientific

### CERTIFICATE OF ANALYSIS

Certificate N°: **0280/2024** Batch number: **P126404A**

Product name: **SPVX02 CTM** Code Patheon: **364101**

Client: **Stablepharma Ltd.**

Package size and type: **Bulk**

**Results**

| Test description | Acceptance Criteria | Results |
|---|---|---|
| Moisture Content (Karl Fisher Method) | ≤ 3.0% | 0.9% |
| Appearance (LYO) | Stoppered and crimped glass vial containing a white lyoplug of approximately 5 mm height without signs of collapse | Stoppered and crimped glass vial containing a white lyoplug of approximately 5 mm height without signs of collapse |
| Appearance (Rec.Sol) | Suspension of white or grey particles dispersed in a colourless or pale yellow liquid and may form a sediment at the bottom of the container | Suspension of white particles dispersed in a colourless liquid and may form a sediment at the bottom of the container |
| pH (Rec.Sol) | 6.0 – 7.0 | 6.5 |
| Extractable volume | Not less than 0.5 ml (1 dose) | 0.5 ml |
| Osmolality | 500-750 mOsm/Kg | 687 mOsm/Kg |
| Reconstitution time | <1 minute with gentle mixing – no visible residue as an undissolved matter | 21 seconds – no visible residue as an undissolved matter |
| Sterility (direct inoculation) | Sterile | I° test: Sterile<br>II° test: Sterile |
| Bacterial Endotoxins | ≤ 20 EU/mL | < 5 EU/mL |

The material identified above has been analyzed and found to be COMPLIANT with release specification SPFS0692.

Printed on

Monza, 14/11/2024

Quality Control Manager

*Electronically signed by: Riccardo Taribello*
*Reason: Approver of the GxP document*
*Date: Nov 14, 2024 14:10 GMT+1*

1504 София, бул. Янко Сакъзов 26
Факс: **359 2 943 34 55
Телефон Управител:
**359 2 944 61 91


1504 Sofia, 26 Yanko Sakazov blvd.
Fax: **359 2 943 34 55
Telephone General Manager:
**359 2 944 61 91


### CERTIFICATE of ANALYSIS

#### TETADIF

Tetanus - Diphtheria toxoid (adsorbed) (with reduced dose of diphtheria for adults)
Susp. inj., amp – 0.5 ml - 1 dose, IM

**Lot №: D0727-02**

Date of manufacture: April 2023

| № | Test / Method | Acceptance Criteria | Result |
|---|---|---|---|
| 1. | Appearance | Suspension of white or gray particles dispersed in colorless or pale yellow liquids and may form a sediment at the bottom of the container | Pass |
| 2. | Identity | Identical | Pass |
| 3. | pH | 6.0 – 7.0 | 6.38 |
| 4. | Adjuvant | Aluminium hydroxide not more than 1.25 mg / 0,5ml | 0.59 mg / 0.5 ml |
| 5. | Sodium chloride | 7.0 – 10.0 g / L | 8.63 g / L |
| 6. | Free formaldehyde | Not more than 0.2 g / L | < 0.2 g / L |
| 7. | Sterility | Sterile | Sterile |
| 8. | Degree of adsorption* | Not less than 80 per cent | 100 per cent |
| 9. | Specific toxicity - in guinea-pigs* | None of the animals shows abnormal reaction | Pass |
| 10. | Potency*<br>- tetanus component<br>-diphtheria component | Lower confidence limit (P=0.95) of the estimated potency<br>Not less than 20.0 IU/dose<br>Not less than 2.0 IU/dose | 82.22 IU / 0,5 ml (50.06-135.23)<br>10.74 IU / 0,5 ml (6.08-19.16) |
| 11. | Extractable volume | Not less than 0.5 ml | Pass |
| 12. | Storage condition | Store at 2°C - 8°C; Do not freeze | |
| 13. | Expiry date | 36 months of the start date of the last satisfactory potency determination (i.e. the date on which the test animals were immunized with the vaccine) is taken as the start date for the shelf-life | March 2026 |

* - Test is performed at stage final bulk

***Conclusion:***

**Tetadif Lot № D0727-02 amp – 0.5 ml – 1 dose** meets all national requirements and satisfies Part A of the WHO Recommendations to assure the quality, safety and efficacy of DT based vaccines (WHO TRS No.980, 2014) and European Pharmacopoeia.
The Vaccine has been produced in a firm with approved Quality Management System Standards: ISO 9001:2015 and ISO 14001:2016.

**Date of issue:** 27.06.2023

**Head of Quality Control Department:**
V. Milanova, PhD

Confidentiality Level 1

### Certificate of Analysis (CoA)
### diTeBooster

### Batch DT411A

Batch no.: DT411A
Expiry date: 05-2026

Volume of bulk: 10565.4 mL
Volume of final bulk: 60.0 L

**Used components:**
Diphtheria toxoid AJV bulk, batch no.: D 94
Tetanus toxoid AJV bulk, batch no.: T 332

| **Test** | **Results** | **Requirements** |
|---|---|---|
| **BULK (B)** | | |
| B 1 Lf- concentration | Di: 13.1 Lf/mL<br>Te: 12.6 Lf/mL | Di: 12.5 ± 2.5 Lf/mL final bulk<br>Te: 12.5 ± 2.5 Lf/mL final bulk |
| **FINAL BULK (FB)** | | |
| FB 1 Sterility | No growth | No growth |
| **FINAL LOT (FL)** | | |
| Before labelling (FL-) | | |
| FL 1 Al-concentration | First, day 1: 0.9 mg Al/mL<br>Last, day 1: 1.0 mg Al/mL | 0.9-1.1 mg Al/mL |
| FL 2 Sterility | No growth | No growth |
| FL 3 Pyrogens | Complies<br>(0.10°C/3 rabbits) | Complies<br>(≤ 1.15°C/3 rabbits)<br>(≤ 2.80°C/6 rabbits) |
| FL 4 pH | 7.2 | 6.5 ≤ pH ≤ 7.5 |
| FL 5<br>Di:Potency lower confidens limit | Di: 21 IU/0.5mL | Di: $P=0.95_{\text{lower c.l.}}$ ≥2 IU/0.5mL |
| Te Potency | Te: ≥ 20 IU/0.5mL | Te: ≥ 20 IU/0.5mL |

After labelling (FL+)

| | | |
|---|---|---|
| FL 6 Volume | 0.58 mL/syringe | 0.56-0.61 mL/syringe |
| FL 7 Appearance | Suspension of white particles in colourless liquid | Suspension of white or grey particles in colourless or light yellow liquid |
| FL 8 Identification | Di: Positive in eluate<br>Te: Positive in eluate<br>aP: Negative in eluate | Di and Te: Positive in eluate<br><br>aP: Negative in eluate |
| FL 9 Degree of adsorption | Di: 100% (0 Lf/mL in supernatant)<br>Te: 100% (0 Lf/mL in supernatant) | Di: 100% (0 Lf/mL in supernatant)<br>Te: 100% (0 Lf/mL in supernatant) |

Date of release: 18.04.2024

18-04-2024

X

Signed by: Kim Skjold Rehmeier (KIRE)

*Delegated Qualified Person*
*MSc Pharmacy*
*QA Operations Department*
*Quality Assurance Area*

### SUMMARY OF PRODUCT CHARACTERISTICS

#### 1. NAME OF THE MEDICINAL PRODUCT

**TETADIF suspension for injection**
*Diphtheria and tetanus vaccine (adsorbed, reduced antigen(s) content)*

#### 2. QUALITATIVE AND QUANTITATIVE COMPOSITION

TETADIF is a suspension of purified and adsorbed tetanus and diphtheria toxoid onto aluminum adsorbent. They are produced from toxins of *Corynebacterium diphtheriae* and *Clostridium tetani* by detoxification and purification. Thiomersal is used as a preservative in the manufacturing of multi-dose vials. No preservative are used in the manufacturing of single-dose product (ampoules).

**One dose (0.5 ml) contains:**

| | |
|---|---|
| Purified tetanus toxoid* | not less than 40 International Units (IU) (10.0 Lf) |
| Purified diphtheria toxoid* | not less than 4 International Units (IU) (3.0 Lf) |

*Adsorbed onto aluminium hydroxide ($Al^{+++}$)  not more than 1.25 mg

The vaccine in multi-dose vials contains the preservative thiomersal – not less than 0.05 mg.
See section 4.3.

Excipient with known effect
The vaccine contains less than 1 mmol sodium (23 mg) per dose, i.e. it is essentially 'sodium-free'.

For the full list of excipients, see section 6.1.

#### 3. PHARMACEUTICAL FORM

Suspension for injection.

TETADIF is a suspension of white or grey particles dispersed in colourless or pale yellow liquids.

#### 4. CLINICAL PARTICULARS

##### 4.1 Therapeutic indications

TETADIF is indicated for:
1. Re-immunization against tetanus and diphtheria of children over 7 years of age and adults.
2. Primary immunization against tetanus and diphtheria started after 7 years of age.
3. Immunization after injury or burn with a risk of tetanus infection, if re-immunization against diphtheria in necessary.

##### 4.2 Posology and method of administration

Posology
The single immunization dose of TETADIF is 0.5 ml.
The vaccination schedule shell be in accordance with the National or Referral Immunization Calendar.

*Primary immunization*
Primary immunization against tetanus and diphtheria, started after 7 years of age is performed three times, administered deep INTRAMUSCULARLY into the deltoid muscle of the arm: the first two doses of 0.5 ml at intervals of not less than 30 days between each dose and the third dose of 0.5 ml -

from 6 to 12 months after the second dose. In case of missed a dose, it should be administered whenever is possible.

*Re-immunization*
Subsequent re-immunizations have to comply with a re-immunization schedule according to the National or Referral Immunization Calendar.

*Vaccination following injuries or burns* with a risk of tetanus infection, if re-immunization against diphtheria is necessary, is performed by administering 0.5 ml INTRAMUSCLARLY, deep into the deltoid muscle of the arm.

*Paediatric population*
The safety and efficacy of TETADIF in children below 7 years of age have not been established.

Method of administration
The vaccine is administered deep INTRAMUSCLARLY (i.m.) into the deltoid muscle of the arm. The ampoule/ vial must be shaken prior to injection until a homogenous whitish suspension is obtained.
The vaccine is drawn from the ampoule (vial) using a sterile syringe and needle. After the air bubbles have been expelled from the syringe, the needle is replaced with a new dry sterile needle which is used for the injection.

#### 4.3 Contraindications

Hypersensitivity to the active substance(s) or to any of the excipients of the vaccine listed in section 6.1.

*The drug product contains thiomersal, as a preservative in multi-dose vials and is possible to cause an allergic reaction to you/ your child.*

Anamnestic data for hypersensitivity reactions to previous vaccine administration.
General contraindications for immunizations:
1. Acute infectious diseases, convalescence period included.
2. Fever conditions.
3. Active tuberculosis.
4. Decompensated heart failure.
5. Diabetes mellitus, thyrotoxicosis, decompensated adrenal insufficiency.
6. Acute central nervous system inflammations – meningites, encephalites, meningoencephlalites.
7. Chronic active hepatites and biliary cirrhosis.
8. Infections of the urinary tract.
9. Nephrotic syndrome.
10. Autoimmune diseases.
11. Allergy, including medical data for shock, Oedema Quincke and other severe allergic reactions to allergens present in vaccines.
12. Epilepsy.

At the presence of contraindications the physician assesses the risk of TETADIF administration or tetanus or diphtheria disease.

#### 4.4 Special warnings and precautions for use

Vaccine administration needs to be preceded by taking the patient’s medical history and performing a medical examination.
HIV infected patients are immunized, resp. re-immunized with TETADIF following the standard immunization schedules.
The effect of immunization is possible to be reduced in immunodeficiencies or in patients subjected to immunosuppressive treatment.

In such cases, it is recommended to postpone the immunization (if scheduled) until the immunosuppressive treatment is withdrawn.
Caution should be exercised in patients with coagulation disorders. The vaccinated person should remain under medical supervision for 30 minutes after immunization due to the possibility of an anaphylactic reaction.

<u>Traceability</u>
In order to improve the traceability of biological medicinal products, the name and the batch number of the administered product should be clearly recorded.

#### 4.5 Interaction with other medicinal products and other forms of interaction

Immunization (resp. re-immunization) with TETADIF may be administered concomitantly with other vaccines. TETADIF is compatible with poliomyelitis, hepatitis B, measles, rubella, mumps, rabies, flu and yellow fever vaccines. The products listed above need to be administered using a different needle and syringe and at a different injection site than TETADIF.
The vaccine could be administered either concomitantly or separately in patients with injected immunoglobulins. The immunoglobulin injection site needs to be different from the one used for TETADIF administration.

There are no data available for interaction of the vaccine with other medicinal products.

#### 4.6 Fertility, pregnancy and lactation

<u>Pregnancy</u>
No studies have been done to investigate the reproductive toxicity of the vaccine. However, immunization during pregnancy is not recommended.

<u>Breastfeeding</u>
There are no data indicating administration of the vaccine during the lactation.

<u>Fertility</u>
TETADIF has not been evaluated in fertility studies.

#### 4.7 Effects on ability to drive and use machines

There are no studies indicating that TETADIF may impair the attention and the ability to drive and use machines. TETADIF has no or negligible influence on the ability to drive and use machines.

#### 4.8 Undesirable effects

The adverse reactions after administration of TETADIF are:
*Frequency is not known (cannot be estimated from the available data)*, according MedDRA-system organ class database).
*General disorders and administration site conditions:*
Local reactions may occur at the vaccine injection site, for example: Pain, redness, mild swelling and induration that may resolve in 1 - 2 days.
In some cases, there may be low-grade fever that resolves in 1 - 2 days.

<u>Reporting of suspected adverse reactions</u>
Reporting suspected adverse reactions after authorisation of the medicinal product is important. It allows continued monitoring of the benefit/risk balance of the medicinal product. Healthcare professionals are asked to report any suspected adverse reactions via the national reporting system listed.
You can also report side effects to the manufacturer (See section 7).

#### 4.9 Overdose

An overdose of the single-dose vaccine is unlikely to occur.
It is recommended to carefully review the medical history of the patient with respect to previous immunizations in order to avoid the administration in recently immunized or re-immunized individuals containing tetanus and diphtheria toxoid.

### 5. PHARMACOLOGICAL PROPERTIES

#### 5.1 Pharmacodynamic properties

Pharmacotherapeutic group: Antiinfectives for systemic use, vaccines, bacterial vaccines, tetanus vaccines, tetanus toxoid, combination with diphtheria toxoid.
ATC code: J07AM51

Mechanism of action
TETADIF is a suspension of purified and adsorbed tetanus and diphtheria toxoid onto aluminum adsorbent, prepared from toxins of *Corynebacterium diphtheriae* and *Clostridium tetani* by detoxification followed by purification. The vaccine contains a reduced quantity of the diphtheria antigen. After vaccination, antibodies are produced against the two antigens of the vaccine, providing protection against diphtheria and tetanus. The immunity is strengthened after re-immunization and is considered to last from 5 to 10 years.

Efficiency against diphtheria
The diphtheria toxoid provides protection against diphtheria.
Immunoprophylaxis is the only way to prevent diphtheria. It is performed with diphtheria toxoid, most often combined with tetanus toxoid or killed pertussis bacteria. Children aged between three months to 8 years are subject to vaccination although in recent years, people up to the age of 35 have also been immunized. The mass application of the vaccine in our country has led to the elimination of any incidence of diphtheria in Bulgaria. The data indicated in Table 1 about the immunological status of the population demonstrated that children and persons under 15 years of age show the highest level of protection against diphtheria. The increasing of age leads to decrease in the percentage of protected persons, especially in the adults over 45. However, the percentage of people protected from diphtheria between the ages of 16 and 65 is higher than the rates from research in some European countries and the United States. For example, in Sweden about 56.9% of the population lacks protection against diphtheria, in Germany, this percentage is about 52.2%, and in Denmark about 36%.

**Table 1: Evaluation of the immune status of the population in Bulgaria against diphtheria in different age groups.**

| Age group | % serums with full protection (titer > 0.1 IU) | % serums with basic immunity (titer 0.01 to 0.09 IU) | % serums with no protection (titer < 0.009 IU) |
|---|---|---|---|
| **Children aged 0 to 7 years** | 94.45 | - | 5.55 |
| **Persons aged 8 to 15 years** | 100 | - | - |
| **Persons aged 16 to 25 years** | 81.39 | 2.9 | 15.71 |
| **Persons aged 26 to 35 years** | 78.69 | 3.6 | 17.71 |
| **Persons aged 36 to 45 years** | 77.0 | 1.0 | 22.0 |
| **Persons aged 46 to 55 years** | 61.92 | 1.58 | 36.5 |
| **Persons between 56 to 65 years** | 61.64 | 7.6 | 30.76 |
| **Adults aged over 65 years** | 11.77 | - | 88.23 |

**The results were obtained by ELISA: Enzyme-linked immunoassay;*

Efficiency against tetanus
Tetanus toxoid provides protection against tetanus.

One of the means of treatment tetanus is a specific immune prophylaxis with tetanus toxoid. In human organism it causes formation of specific antibodies, which play role in protection against tetanus. The immunity in tetanus is conditioned and depends on the quantity and possibility of the specific antibodies in short time to neutralize the tetanus toxin.
Tetanus still represents a health problem for many countries around world mainly due to its high mortality.
In Bulgaria the morbidity and mortality from the tetanus were high. Since 1959, obligatory specific immune prophylaxis against tetanus was introduced in Bulgaria, gradually covering the entire population. This led to a significant decrease in disease incidence (from 2.7 %000 in 1959 to 0.01 %000 in 1998). This proves the good protective effect of tetanus toxoid and the optimal scheme of administration, regardless of the age. In comprehensive clinical studies in persons with chronic diseases like allergies, diabetes, rheumatism, uro-genital diseases and persons over 70 years of age, showed good tolerance for these risk group of patients, witch allows their inclusion in the obligatory immunization schedule.

Seroepidemiological studies conducted in recent years showed the highest antitoxic titer among children and adolescent because they are included in the obligatory immunization schedule. Good immunity is established in the other age groups with slight decrease of protective titer, in relation to an insufficient population density. Single cases of disease in these age groups are observed. The observations on tetanus toxoid administration in over 2 000 persons in the last ten years, reveals that it is not reactogenic.

##### 5.3 Preclinical safety data

Non-clinical data reveal no special hazard for humans.

#### 6. PHARMACEUTICAL PARTICULARS

##### 6.1 List of excipients

**in single dose ampoule:**
Aluminium hydroxide
Sodium chloride
Water for injection

**in multi-dose vials:**
Aluminium hydroxide
Thiomersal
Sodium chloride
Water for injection

##### 6.2 Incompatibilities

In the absence of compatibility studies, this medicinal product must not be mixed with other medicinal products.

In concomitant administration with other injectable medicinal products, each individual product needs to be drawn up and administered using a separate syringe and needle, and at a site different than the vaccine injection site.

##### 6.3 Shelf life

3 years

##### 6.4 Special precautions for storage

Store and transport refrigerated (2°C – 8°C).
Store in the original carton package.
Do not freeze!
**A FROZEN VACCINE IS UNFIT FOR USE!**
Keep this medicine out of the sight and reach of children.
Use the vaccine in the multi-dose vial immediately after withdrawing the first dose!
**Any unused vaccine should be discarded!**

##### 6.5 Nature and contents of container

Clear, colourless, flip-off glass ampoule (type I).
The ampoules contain a vaccine in a volume of 0.5 ml – 1 dose.
Pack size: carton box of 1, 10 or 50 ampoules.

Clear, colourless, glass vial (type I).
The vial contains vaccine in volume of 5.0 ml - 10 doses.
The vial contains vaccine in volume of 10.0 ml - 20 doses.
Pack size: carton boxes of 10 vials.

Not all pack sizes may be marketed.

##### 6.6 Special precautions for disposal and other handling

The precipitated vaccine is a clear, colourless supernatant fluid with whitish precipitate.

FROZEN VACCINE IS UNFIT FOR USE!
Do not administer intravenously.
Do not use expired vaccine.
Do not use in case the integrity of the ampoule (vial) is compromised or the labelling is unclear/ deleted.

Any unused medicinal product or waste material should be disposed of in accordance with local requirements.

#### 7. MARKETING AUTHORISATION HOLDER

BB - NCIPD Ltd.
26, Yanko Sakazov Blvd.
1504 Sofia
Bulgaria


#### 8. MARKETING AUTHORISATION NUMBER(S)

II-15236/ 11 October 2011

Registration No 20011159

#### 9. DATE OF FIRST AUTHORISATION/RENEWAL OF THE AUTHORISATION

Date of first authorisation: 28 November 2001
Date of latest renewal: 11 October 2011

#### 10. DATE OF REVISION OF THE TEXT

April 2022

### TECHNICAL DATA SHEET

#### 1. NAME OF THE MEDICINAL PRODUCT

diTeBooster, injectable suspension in single-dose vials or prefilled single-dose syringe.

Diphtheria and tetanus vaccine (absorbed, reduced antigen(s) content).

#### 2. QUALITATIVE AND QUANTITATIVE COMPOSITION

One dose (0.5 ml) contains:

| | |
|---|---|
| Purified diphtheria toxoid[1] | 6.25 Lf / ≥ 2 IU |
| Purified tetanus toxoid[1] | 6.25 Lf / ≥ 20 IU |

[1] Absorbed in hydrated aluminium hydroxide ($AL(OH)_3$), corresponding to 0.5 mg of aluminium ($Al^{3+}$).

The diphtheria and tetanus toxins, obtained from cultures of *Corynebacterium diphtheriae* and *Clostridium tetani*, are purified and detoxified.

No substance of human origin is used during the manufacture of the vaccine.

The vaccine may contain traces of formaldehyde that is used during the manufacturing process (see section 4.4).

For the full list of excipients, see section 6.1.

#### 3. PHARMACEUTICAL FORM

Suspension for injection in single-dose vials or pre-filled syringe (injection). Colourless

or light yellow suspension of white/grey particles.

#### 4. CLINICAL DATA

##### 4.1. Therapeutic indications

Active immunization against tetanus and diphtheria in persons from 5 years of age.

Revaccination against diphtheria and tetanus, and primary immunization of persons with missing, incomplete or unknown primary immunization.

Tetanus prophylaxis in individuals 5 years of age and older with tetanus-prone lesions and concurrent diphtheria immunization.

Use of diTeBooster should be in accordance with official national recommendations.

##### 4.2. Dosage and form of administration

MARÍA CASANI FERNÁNDEZ DE NAVARRETE
Traductor/a Intérprete Jurado/a de INGLÉS
Nº 3692

Dosage

diTeBooster should be administered intramuscularly as a single 0.5 ml dose for all ages.

*Revaccination*
diTeBoost can be used for revaccination of persons who have been previously vaccinated against diphtheria and tetanus according to national recommendations.

An enhanced response can only be expected in persons with primary vaccination.

Repeated vaccination against diphtheria and tetanus should be carried out at intervals according to official recommendations (usually 10 years).

*Primary immunisation*
Persons with missing, incomplete, or unknown primary immunization may be vaccinated with diTeBooster. More than one vaccine may be needed to achieve protective immunity against diphtheria and tetanus. National recommendations should be followed.

*Tetanus-prone lesions*
In people with tetanus-prone lesions, diTeBooster may be given when diphtheria vaccination is also relevant. Tetanus immunoglobulin can be administered simultaneously according to national recommendations.

*Pediatric population*
Safety and efficacy of diTeBooster in children under the age of 5 years has not been established. No data available.

Method of administration

diTeBooster should be administered intramuscularly (IM), preferably in the deltoid region.

Do not inject intravascularly.

Shake before use.

In certain indications (e.g. hemorrhagic diathesis) diTeBooster can be administered deep subcutaneously.

**4.3. Contraindications**

Serious adverse reactions after previous vaccination with the vaccine or hypersensitivity to the active ingredient(s), or to any of the excipients included in section 6.1.

**4.4. Special warnings and precautions for use**

- As with all injectable vaccines, adequate treatment and medical supervision should always be available in the event of a rare anaphylactic episode following vaccine administration.
- Vaccination should be postponed in case of acute illness with fever.
- Under no circumstances should diTeBooster be administered intravascularly.
- As with any injectable vaccine, diTeBooster should be administered with caution to persons with uncontrolled coagulopathy, as bleeding may occur following intramuscular administrations.
- In individuals with compromised immune response, the serological response may be altered. Vaccination may be carried out in persons receiving immunosuppressive therapy, although a decreased immune response may occur.
- The final product may contain traces of formaldehyde that is used during the process of

MARÍA CASANI FERNÁNDEZ DE NAVARRETE
Traductor/a-Intérprete Jurado/a de INGLÉS
Nº 3692

MINISTERIO DE SANIDAD POLÍTICA SOCIAL E IGUALDAD
Agencia española de medicamentos y

manufacturing. Caution should be exercised in subjects with known hypersensitivity to formaldehyde.

- diTeBooster contains less than 1 mmol sodium (23 mg) per dose, i.e. essentially 'sodium-free'.
- Too frequent booster vaccination will increase the risk of adverse reactions.

Traceability
In order to improve the traceability of biological drugs, the name and batch number of the drug administered should be clearly recorded.

**4.5. Interaction with other drugs and other forms of interaction**

Do not mix with other vaccines in the same syringe or vial.

The concomitant use of diTeBooster with other inactivated vaccines has not been studied. Co-administration is unlikely to result in interference with immune responses. When deemed necessary, diTeBooster can be administered simultaneously with other vaccines, at a different injection site.

If immediate protection is needed, diTeBooster can be given at the same time as tetanus immunoglobulin. Injections of diTeBooster and tetanus immunoglobulin should be done in separate limbs.

**4.6. Fertility, pregnancy and breastfeeding**

Pregnancy

No animal data are available. In humans, data are inadequate to assess the risk of teratogenicity or phototoxicity during pregnancy.
During pregnancy, the potential risk of clinical infection after exposure should be assessed against the theoretical risks of the vaccine.

Breastfeeding

There is no indication that vaccination of nursing mothers with diTeBooster is harmful to the child. Fertility

The effect on reproductive organs has not been studied in developmental toxicology studies. However, there is no indication that vaccination has an effect on male and female fertility.

**4.7. Effects on the ability to drive and use machines**

The influence of diTeBooster on the ability to drive and use machines is null or negligible.

**4.8. Adverse reactions**

In relation to the administration of diTeBooster, the most common adverse reactions are: Redness and swelling at the injection site and fever. Reactions usually start within 48 hours of vaccination.

List in table form of adverse reactions.
The adverse reactions listed below are based on data from clinical trials in children, adolescents, and adults, and are classified according to MedDRA system organ class terminology.
The safety evaluation of diTeBooster also includes adverse reactions from clinical trials and spontaneous reports with vaccines containing the same or higher content of diphtheria and tetanus antigens than diTeBooster in combination with aluminum hydroxide and other vaccine antigens.

MARÍA CASANI FERNÁNDEZ DE NAVARRETE
Traductor/a Intérprete Jurado/a de INGLÉS
Nº 3692

Maria Casa

MINISTERIO DE SANIDAD, POLÍTICA SOCIAL E IGUALDAD
Agencia española de medicamentos y

| Classification by organs, systems and frequency | Adverse reactions |
|---|---|
| **Disorders of the immunological system**<br>Rare ≥ 1/10 000 and < 1/1 000 | Hypersensitivity, including anaphylactic reactions. |
| **Nervous system disorders**<br>Very frequent (≥1/10)<br>Common (≥ 1/100 and <1/10)<br>Very rare < 1/10 000 | Headache.<br>Dizziness<br>Vasovagal syncope |
| **Gastrointestinal disorders**<br>Common (≥ 1/100 and <1/10) | Nausea, vomiting and diarrhoea |
| **Skin and subcutaneous tissue disorders**<br>Uncommon (> 1/ 1 000 and < 1/100)<br>Rare ≥ 1/10 000 and < 1/1 000 | Eczema and dermatitis<br>Urticarial reactions |
| **Musculoskeletal and connective tissue disorders Vascular**<br>Common (≥ 1/100 and <1/10) | Myalgia |
| **General disorders and alterations at the site of administration**<br>Very frequent (≥1/10)<br><br><br><br>Common (≥ 1/100 and <1/10)<br><br><br><br>Rare ≥ 1/10 000 and < 1/1 000 | Redness / Swelling at the injection site*<br>Pain at the site of injection<br>Injection site itching<br>Fatigue<br>Malaise<br>Fever ≥ 38°C<br>Redness/ Swelling ≥ 5 cm at the site of the injection<br>High fever > 40 °C<br>Sterile granuloma or abscess at the injection site. |

*Less frequent redness/swelling at the injection site has been observed in adults.

Pediatric population
The safety evaluation of diTeBooster studied in clinical trials includes children aged 4 years and older and adolescents

Reporting of suspected adverse reactions:

Reporting suspected adverse reactions after authorisation of the drug is important. It allows continued monitoring of the benefit/risk balance of the drug. Healthcare professionals are invited to report suspected adverse reactions through the Spanish Pharmacovigilance System for Medicinal Products for Human Use: www.notificaRAM.es

**4.9. Overdose**

No cases of overdose have been reported.

MARÍA CASANI FERNÁNDEZ DE NAVARRETE
Traductor/a Intérprete Jurado/a de INGLÉS
Nº 3692

MINISTERIO DE SANIDAD POLÍTICA SOCIAL E IGUALDAD
Agencia española de medicamentos y

#### 5. PHARMACOLOGICAL PROPERTIES

##### 5.1. Pharmacodynamic properties

Pharmacotherapeutic group: Tetanus, toxoid, combinations with diphtheria toxoid.
ATC code: J07AM51.

Action mechanism

Antibodies to vaccine antigens are produced shortly after vaccination. Protection against diphtheria and tetanus is expected to last for at least 10 years

Clinical efficacy and safety

Clinical trials have been conducted with diTeBooster in children, adolescents and adults. The immunogenicity assessment of diTeBooster also includes data from clinical trials with vaccines containing the same content of diphtheria and tetanus antigens as diTeBooster in combination with aluminum hydroxide and other vaccine antigens, e.g., acellular pertussis. The results are summarised in the following table.

| Study population | Age | Children 5-6 years | Children 10 years | Teenagers from 14-15 years old | Adults 18-55 years old |
|---|---|---|---|---|---|
| | Vaccination history. | 3 x DTaP First year of life | 3 x DT First year of life | 3 x DTaP First year of life 1 x TdaP 4-6 years old. | 3-4 x D and T First year of life |
| Antigen | Immune system | | | | |
| Diphtheria | ≥ 0.1 IU/mL | 98.6 -100% | - | 100% | 98.5-100% |
| | ≥ 0.01 IU/ml | 100% | 100% | - | 98.8-100% |
| Tetanus | ≥ 0.1 IU/mL | 99.3-100% | 100% | 100% | 99.4- 100 % |
| | ≥ 0.01 IU/ml | 99.3-100% | 100% | - | 99.4- 100 % |

Antibodies to diphtheria and tetanus were measured one month after vaccination.

Diphtheria and tetanus antibody levels of ≥ 0.01 IU/ml are considered the minimum level of antibody required to confer some degree of protection, while levels of at least 0.1 IU/ml are considered protective.

##### 5.2. Pharmacokinetic properties

Evaluation of the pharmacokinetic properties for vaccines is not necessary.

##### 5.3. Preclinical safety data

Subacute and acute toxicity of vaccine components have been studied in animals. No clinical symptoms or systemic toxicity have been recorded.

#### 6 . PHARMACEUTICAL DATA

##### 6.1. List of excipients

Per dose = 0.5 ml:
Sodium hydroxide until pH = 7
Sodium chloride

MARÍA CASANI FERNÁNDEZ DE NAVARRETE
Traductor/a Intérprete Jurado/a de INGLÉS
Nº 3692

MINISTERIO DE SANIDAD, POLITICA SOCIAL E IGUALDAD
Agencia española de medicamentos y

Water for injectable preparations.

The pH of the vaccine is about 7.

For absorbents, see section 2.

**6.2. Incompatibilities**

In the absence of compatibility studies, this vaccine must not be mixed with other vaccines or drugs.

**6.3. Validity period**

3 years.

**6.4. Special storage precautions**

Store in a refrigerator (2°C to 8°C).
Do not freeze.
Discard if vaccine has been frozen.

**6.5. Nature and content of the packaging**

Single-dose vials (type I glass) closed with a stopper (chlorobutyl rubber) containing 0.5 ml (1 dose).
Package size: 1 x 0.5 ml, 5 x 0.5 ml and 10 x 0.5 ml.
The vial caps do not contain latex.

Single-dose pre-filled syringe (type I glass) with 0.5 ml (1 dose).
Package size: 1 x 0.5 ml, 5 x 0.5 ml, 10 x 0.5 ml and 20 x 0.5 ml.
Only some package sizes may be commercially available.

**6.6. Special precautions for disposal and further handling**

Shake before use.

After thorough resuspension, the vaccine should show up as a colorless or light yellow suspension of white and gray particles.

Examine the vaccine for foreign particles and/or discoloration before use. If these conditions are met, the product should not be administered

Disposal of the unused drug and all materials that have been in contact with it should be carried out in accordance with local regulations.

**7. MARKETING AUTHORIZATION HOLDER**

AJ vaccines A/S
Artillerivej 5
DK-2300 Copenhagen S
Denmark

MARÍA CASANI FERNÁNDEZ DE NAVARRETE
Traductor/a Intérprete Jurado/a de INGLÉS
Nº 3692

**8. MARKETING AUTHORISATION NUMBER(S)**

Pre-filled syringe: 68579

MINISTERIO DE SANIDAD, POLÍTICA SOCIAL E IGUALDAD
Agencia española de medicamentos y

#### 9. DATE OF THE FIRST AUTHORIZATION/ RENEWAL OF THE AUTHORIZATION

Date of first authorisation: Pre-filled syringe: 28/March/2007
Date of last renewal: Pre-filled syringe: 25/May/2010

#### 10. DATE OF REVISION OF THE TEXT

01/2024

**Don/Doña MARÍA CASANI FERNÁNDEZ DE NAVARRETE, Traductor/a-Intérprete Jurado/a de nombrado/a por el Ministerio de Asuntos Exteriores y de Cooperación, CERTIFICA que lo que ante traducción fiel y completa al inglés de un documento redactado en español.**

**Mr/Ms. MARÍA CASANI FERNÁNDEZ DE NAVARRETE, Official English Translator/Interpreter of appointed by the Spanish Ministry of Foreign Affairs and Cooperation, HEREBY CERTIFIES that the fore an accurate and complete translation to English of the original document, written in Spanish.**

**En Madrid a dieciséis de diciembre de dos mil veinticuatro.**

MARÍA CASANI FERNÁNDEZ DE NAVARRETE
Traductor/a Intérprete Jurado/a de INGLÉS
Nº 3692

MINISTERIO DE SANIDAD, POLÍTICA SOCIAL E IGUALDAD

### FICHA TÉCNICA

#### 1. NOMBRE DEL MEDICAMENTO

diTeBooster, suspensión inyectable en viales monodosis o jeringa monodosis precargada.

Vacuna contra la difteria y el tétanos (adsorbida, contenido en antígeno(s) reducido).

#### 2. COMPOSICIÓN CUALITATIVA Y CUANTITATIVA

Una dosis (0,5 ml) contiene:

| | |
|---|---|
| Toxoide diftérico purificado[1] | 6,25 Lf / $\geq$ 2 UI |
| Toxoide tetánico purificado[1] | 6,25 Lf / $\geq$ 20 UI |

[1] Absorbido en hidróxido de aluminio hidratado (AL(OH)$_3$), correspondiente a 0,5 mg de aluminio ($Al^{3+}$).

Las toxinas difetérica y tetánica, obtenidas a partir de cultivos de *Corynebacterium diphtheriae* y *Clostridium tetani*, se purifican y detoxifican.

Durante la fabricación de la vacuna no se utiliza ninguna sustancia de origen humano.

La vacuna puede contener trazas de formaldehido que se utiliza durante el proceso de fabricación (ver sección 4.4).

Para consultar la lista completa de excipientes ver sección 6.1.

#### 3. FORMA FARMACÉUTICA

Suspensión inyectable en viales monodosis o jeringa precargada (inyectable).

Suspensión incolora o amarilla clara de partículas blancas/grises.

#### 4. DATOS CLÍNICOS

##### 4.1. Indicaciones terapéuticas

Inmunización activa contra el tétanos y la difteria en individuos a partir de 5 años de edad.

Revacunación contra la difteria y el tétanos, e inmunización primaria de individuos con inmunización primaria faltante, incompleta o desconocida.

Profilaxis del tétano en individuos a partir de 5 años de edad con lesiones propensas al tetanos y con inmunización simultaneo contra la difteria.

El uso de diTeBooster debe realizarse siguiendo las recomendaciones oficiales nacionales.

##### 4.2. Posología y forma de administración

Posología

diTeBooster debe administrase por vía intramuscular en forma de una dosis única de 0,5 ml para todas las edades.

MARÍA CASAN FERNÁNDEZ DE NAVARRETE
Traductor/a Intérprete Jurado/a de INGLÉS
Nº 3692

*Revacunación*
diTeBoost puede ser utilizado para la revacunación de individuos que han sido previamente vacunados contra la difteria y el tétanos según las recomendaciones nacionales.

Solo se puede esperar una respuesta potenciada en los individuos con vacunación primaria.

La vacunación repetida frente a la difteria y el tétanos debe realizarse a intervalos según las recomendaciones oficiales (generalmente 10 años).

*Inmunización primaria*
Los individuos con inmunización primaria faltante, incompleta o desconocida pueden ser vacunados con diTeBooster. Es posible que se necesite más de una vacuna para conseguir una inmunidad protectora contra la difteria y el tétanos. Deben seguirse las recomendaciones nacionales.

*Lesiones propensas al tétanos*
En las personas con lesiones propensas al tétanos, se puede administrar diTeBooster cuando la vacunación contra la difteria también es pertinente. La inmunoglobulina del tétanos puede administrarse simultáneamente de acuerdo a las recomendaciones nacionales.

*Población pediátrica*
No se ha establecido la seguridad y eficacia de diTeBooster en niños menores de 5 años. No se dispone de datos.

<u>Forma de administración</u>

diTeBooster debe administrarse por vía intramuscular (IM), preferiblemente en la región del deltoides.

No se debe inyectar por vía intravascular.

Agitar antes de usar.

En ciertas indicaciones (por ejemplo, diátesis hemorrágica) se puede administrar diTeBooster por vía subcutánea profunda.

**4.3. Contraindicaciones**

Reacciones adversas graves tras vacunación anterior con la vacuna o hipersensibilidad al (a los) principio(s) activo(s), o a alguno de los excipientes incluidos en la sección 6.1.

**4.4. Advertencias y precauciones especiales de empleo**

- Al igual que todas las vacunas inyectables siempre debe disponerse de un tratamiento y una supervisión médica adecuados en caso de que se produzca un episodio anafiláctico poco frecuente después de la administración de la vacuna.
- La vacunación debe posponerse en caso de enfermedad aguda con fiebre.
- Bajo ninguna circunstancia debe administrarse diTeBooster por via intravascular.
- Como con cualquier vacuna inyectable, diTeBooster debe administrarse con precaución a los individuos con coagulopatía no controlada, ya que puede producirse una hemorragia tras las administraciones intramusculares.
- En individuos con la respuesta inmunitaria comprometida, puede alterarse la respuesta serológica. Puede llevarse a cabo la vacunación en individuos que reciban tratamiento inmunosupresor, aunque puede producirse una menor respuesta inmunológica.
- El producto final puede contener trazas de formaldehído que se utiliza durante el proceso de

Traductor/a Intérprete Jurado/a de INGLÉS
Nº 3692

MINISTERIO DE SANIDAD, POLÍTICA SOCIAL E IGUALDAD
Agencia española de medicamentos y productos sanitarios

fabricación. Debe tenerse precaución en sujetos con hipersensibilidad conocida al formaldehído.

- diTeBooster contiene menos de 1 mmol de sodio (23 mg) por dosis; esto es, esencialmente "exento de sodio".
- Una vacunación de refuerzo demasiado frecuente aumentará el riesgo de reacciones adversas.

Trazabilidad

Con objeto de mejorar la trazabilidad de los medicamentos biológicos, el nombre y el número de lote del medicamento administrado deben estar claramente registrados.

**4.5. Interacción con otros medicamentos y otras formas de interacción**

No mezclar con otras vacunas en la misma jeringa o vial.

No se ha estudiado el uso concomitante de diTeBooster con otras vacunas inactivadas. Es improbable que la coadministración pueda originar una interferencia con las respuestas inmunitarias. Cuando se considere necesario, diTeBooster puede administrarse simultáneamente con otras vacunas, en un lugar de inyección distinto.

Si es necesario proporcionar una protección inmediata, se puede administrar diTeBooster al mismo tiempo que la inmunoglobulina del tétanos. Las inyecciones de diTeBooster y la inmunoglobulina del tétanos deben hacerse en miembros separados.

**4.6. Fertilidad, embarazo y lactancia**

Embarazo

No se dispone de datos relevantes en animales. En humanos, los datos son inadecuados para evaluar el riego de teratogenicidad o de fototoxicidad durante el embarazo.
Durante el embarazo debe evaluarse el posible riesgo de infección clínica después de la exposición frente a los riesgos teóricos de la vacuna.

Lactancia

No hay indicios de que la vacunación de madres lactantes con diTeBooster sea perjudicial para el niño.

Fertilidad

El efecto sobre los órganos reproductores no se ha estudiado en estudios toxicológicos del desarrollo. Sin embargo, no hay indicios de que la vacunación tenga un efecto sobre la fertilidad masculina y femenina.

**4.7. Efectos sobre la capacidad para conducir y utilizar máquinas**

La influencia de diTeBooster sobre la capacidad para conducir y utilizar máquinas es nula o insignificante.

**4.8. Reacciones adversas**

En relación con la administración de diTeBooster, las reacciones adversas más frecuentes son:
Enrojecimiento e hinchazón en el lugar de la inyección y fiebre. Las reacciones se inician habitualmente durante las 48 horas posteriores a la vacunación.

Lista tabulada de reacciones adversas.

Las reacciones adversas que se enumeran a continuación se basan en datos de ensayos clínicos en niños, adolescentes y adultos, y se clasifican según la terminología MedDRA de clasificación por órganos y sistemas.
La evaluación de seguridad de diTeBooster también incluye reacciones adversas de los ensayos clínicos e informes espontáneos con vacunas que contienen el mismo o mayor contenido de antígenos de la difteria y el tétanos que diTeBooster en combinación con hidróxido de aluminio y otros antígenos de la vacuna.

MARÍA CASANI FERNÁNDEZ DE NAVARRETE
Traductor/a Intérprete Jurado/a de INGLÉS
Nº 3692

MINISTERIO DE SANIDAD, POLÍTICA SOCIAL E IGUALDAD
Agencia española de medicamentos y productos sanitarios

| Clasificación por órganos, sistemas y frecuencia | Reacciones adversas |
|---|---|
| **Trastornos del sistema inmunológico**<br>Raras (≥1/10 000 y <1/1 000) | <br>Hipersensibilidad, incluyendo reacciones anafilacticas. |
| **Trastornos del sistema nervioso**<br>Muy frecuentes (≥1/10)<br>Frecuentes (≥1/100 y <1/10)<br>Muy raras (<1/10 000) | <br>Dolor de cabeza.<br>Mareos<br>Síncope vasovagal |
| **Trastornos gastrointestinales**<br>Frecuentes (≥1/100 y <1/10) | <br>Nausea, vomito y diarrea |
| **Trastornos de la piel y del tejido subcutáneo**<br>Poco frecuentes (≥1/1 000 y <1/100)<br>Raras (≥1/10 000 y <1/1 000) | <br>Eccema y dermatitis<br>Reacciones de urticaria |
| **Trastornos musculoesqueléticos y del tejido conjuntivo**<br>Frecuentes (≥1/100 y <1/10) | <br>Mialgia |
| **Trastornos generales y alteraciones en el lugar de administración.**<br>Muy frecuentes (≥1/10)<br><br><br><br><br>Frecuentes (≥1/100 y <1/10)<br><br><br><br>Raras (≥1/10 000 y <1/1 000) | <br>Enrojecimiento / Hinchazón en el lugar de la inyección*<br>Dolor en el lugar de la inyección<br>Picazón en el lugar de la inyección<br>Fatiga<br>Malestar<br>Fiebre ≥ 38 °C<br>Enrojecimiento/ Hinchazón ≥ 5 cm en el lugar de la inyección<br>Fiebre alta > 40 °C<br>Granuloma o abceso esteril en el lugar de la inyección. |

*En los adultos se ha observado un enrojecimiento/hinchazón menos frecuente en el lugar de la inyección.

Población pediátrica
La evaluación de seguridad de diTeBooster estudiado en los ensayos clínicos incluye a niños a partir de 4 años y adolescentes

Notificación de sospechas de reacciones adversas:

Es importante notificar sospechas de reacciones adversas al medicamento tras su autorización. Ello permite una supervisión continuada de la relación beneficio/riesgo del medicamento. Se invita a los profesionales sanitarios a notificar las sospechas de reacciones adversas a través del Sistema Español de Farmacovigilancia de medicamentos de Uso Humano: www.notificaRAM.es.

**4.9. Sobredosis**

No se han registrado casos de sobredosis.

MARÍA CASANI FERNÁNDEZ DE NAVARRETE
Traductor/a Intérprete Jurado/a de INGLÉS
Nº 3692

#### 5. PROPIEDADES FARMACOLÓGICAS

##### 5.1. Propiedades farmacodinámicas

Grupo farmacoterapéutico: Tetánico, toxoide, combinaciones con toxoide diftérico.
Código ATC: J07AM51.

Mecanismo de acción

Poco después de la vacunación se producen anticuerpos contra los antígenos de la vacuna. Se espera una protección contra la difteria y el tétanos con una duración de al menos 10 años

Eficacia clínica y seguridad

Se han realizado ensayos clínicos con diTeBooster en niños, adolescentes y adultos. La evaluación de la inmunogenicidad de diTeBooster también incluye datos de ensayos clínicos con vacunas que contienen el mismo contenido de antígenos de la difteria y el tétanos que diTeBooster en combinación con hidróxido de aluminio y otros antígenos de la vacuna, por ejemplo, la tos ferina acelular. Los resultados se resumen en el cuadro que figura a continuación.

| Población dc estudio | Edad | Niños de 5-6 años | Niños de 10 años | Adolescentes de 14-15 años | Adultos de 18-55 años |
|---|---|---|---|---|---|
| | Historial de vacunación. | 3 x DTaP Primer año de vida | 3 x DT Primer año de vida, | 3 x DTaP Primer año de vida<br>1 x TdaP 4-6 años de edad | 3-4 x D and T Primer año de vida |
| Antígeno | Respuesta inmune | | | | |
| Difteria | ≥ 0,1 UI/ml | 98,6-100 % | - | 100 % | 98,5-100 % |
| | ≥ 0,01 UI/ml | 100 % | 100 % | - | 98,8-100 % |
| Tétanos | ≥ 0,1 UI/ml | 99,3-100 % | 100 % | 100 % | 99,4-100 % |
| | ≥ 0,01 UI/ml | 99,3-100 % | 100 % | - | 99,4-100 % |

Los anticuerpos contra la difteria y el tétanos se midieron un mes después de la vacunación.

Los niveles de anticuerpos contra la difteria y el tétanos de ≥ 0,01 UI/ml se consideran el nivel mínimo de anticuerpos requerido para conferir cierto grado de protección, mientras que los niveles de al menos 0,1 UI/ml se consideran protectores.

##### 5.2. Propiedades farmacocinéticas

La evaluación de las propiedades farmacocinéticas para las vacunas no es necesaria.

##### 5.3. Datos preclínicos sobre seguridad

Se han estudiado en animales la toxicidad subaguda y aguda de los componentes de la vacuna. No se han registrado síntomas clínicos ni toxicidad sistémica.

#### 6 . DATOS FARMACÉUTICOS

##### 6.1. Lista de excipientes

Por dosis = 0,5 ml:
Hidróxido de sodio hasta pH = 7
Cloruro de sodio

MARÍA CASANI FERNÁNDEZ DE NAVARRETE
Traductor/a Intérprete Jurado/a de INGLÉS
Nº 3692

Agua para preparaciones inyectables.

El pH de la vacuna es aproximadamente 7.

Para los adsorbentes, ver la sección 2.

**6.2. Incompatibilidades**

En ausencia de estudios de compatibilidad, esta vacuna no debe mezclarse con otras vacunas ni medicamentos.

**6.3. Periodo de validez**

3 años.

**6.4. Precauciones especiales de conservación**

Conservar en nevera (entre 2 ºC y 8 ºC).
No congelar.
Desechar si la vacuna ha sido congelada.

**6.5. Naturaleza y contenido del envase**

Viales monodosis (vidrio tipo I) cerrados con un tapón (goma de clorobutilo) que contiene 0,5 ml (1 dosis).
Tamaño del envase: 1 x 0,5 ml, 5 x 0,5 ml y 10 x 0,5 ml.
Los tapones de los viales no contienen látex.

Jeringa monodosis precargada (vidrio tipo I) con 0,5 ml (1 dosis).
Tamaño del envase: 1 x 0,5 ml, 5 x 0,5 ml, 10 x 0,5 ml y 20 x 0,5 ml.
Puede que solamente estén comercializados algunos tamaños de envases.

**6.6. Precauciones especiales de eliminación y otras manipulaciones**

Agitar antes de usar.

Tras la resuspensión a fondo, la vacuna debe mostrarse como una suspensión incolora o amarilla clara de partículas blancas y grises.

Examine la vacuna para ver si hay partículas extrañas y/o decoloración antes de usarla. Si se dan estas condiciones, el producto no debe ser administrado

La eliminación del medicamento no utilizado y de todos los materiales que hayan estado en contacto con él, se realizará de acuerdo con la normativa local.

#### 7. TITULAR DE LA AUTORIZACIÓN DE COMERCIALIZACIÓN

AJ Vaccines A/S
Artillerivej 5
DK-2300 Copenhague S
Dinamarca

#### 8. NÚMERO(S) DE AUTORIZACIÓN DE COMERCIALIZACIÓN

MARÍA CASANI FERNÁNDEZ DE NAVARRETE
Traductor/a Intérprete Jurado/a de INGLÉS
Nº 3692

Jeringa precargada: 68579

#### 9. FECHA DE LA PRIMERA AUTORIZACIÓN/ RENOVACIÓN DE LA AUTORIZACIÓN

Fecha de la primera autorización: Jeringa precargada: 28/marzo/2007
Fecha de la última renovación: Jeringa precargada: 25/mayo/2010

#### 10. FECHA DE LA REVISIÓN DEL TEXTO

01/2024

MARÍA CASANI FERNÁNDEZ DE NAVARRETE
Traductor/a Intérprete Jurado/a de INGLÉS
Nº 3692

#### 8. ELISA Method: Anti-Diphtheria Toxoid and Anti-Tetanus Toxoid

Anti-Diphtheria toxoid and Tetanus toxoid IgG concentrations in serum were measured using validated enzyme linked immunosorbent assays (ELISAs). Antibody concentration were measured relative to the International Standard for Tetanus Immunoglobulin human (NIBSC product number 26/488) or the International Standard for Diphtheria Antitoxin Human (NIBSC product number 10/262). Nunc® MaxiSorp™ plates were coated with either Diphtheria toxoid or Tetanus toxoid antigen by overnight incubation at 2-8°C, with antigen diluted in 15 mM sodium carbonate, 35 mM sodium bicarbonate buffer, pH9.6. The next day the coated plates were washed five times with 300µl of phosphate buffered saline containing 0.1%(v/v) Tween 20 (PBST) and blocked with PBST containing 5% (v/v) foetal bovine serum (FBS, Sigma Aldrich) for 60 min at 20 °C ±2 °C. Test samples, standard serum and quality control (QC) serum dilutions were prepared in PBST, 5%(v/v) FBS in a separate dilution plate (Sterilin™) using a starting dilution of 1/125 and seven further three-fold serial dilutions down the plate. The blocking buffer from the coated plates was removed and 100µl/well of each serial dilution from the dilution plates was transferred to the coated plate. Plates were incubated with sera for 60 min±10min at 20 °C±2 °C to facilitate antibody binding and then washed five times with 300µl of PBST to wash away unbound antibodies. Bound antibody was detected using a goat anti-human IgG Fcγ antibody conjugated to alkaline phosphatase (Jackson ImmunoResearch), diluted in PBST to 1/5000, and incubated with 100µl/well for 60 min+10min at 20 °C±2 °C. After washing five times with PBST, 100µl/well of Bio FX™ AP yellow substrate (p-nitrophenyl phosphate, Surmodics Inc.) was added and plates were incubated for 60 min±10min at 20 °C ±2 °C after which time the reaction was stopped by the addition of 50µl/well of 3M sodium hydroxide. Five minutes after stopping the reaction, the optical density (OD, A405nm-A690nm) of each well was measured on a VersaMax™ plate reader using Softmax® Pro GxP software customised protocols. The protocols were validated are designed to process the raw data by fitting an unweighted four parameter logistic curve to the standard serum dose response data on each plate. OD values from each serial dilution for each test sample and QC serum were used to interpolate concentrations from the standard dose response curve run on the same plate and a mean of acceptable values for each sample was calculated. System suitability criteria (for the assay plate and test sample) were applied and reportable values were assigned following defined rules to meet data integrity criteria.

### Annex 12

### Diphtheria antitoxin assay in Vero cell (in vitro toxin neutralization)

The method described here involves staining for cell viability with the yellow tetrazolium salt (MTT), adapted from NIBSC method (see below link).

#### A12.1 MATERIALS

##### 12.1.1 CRITICAL REAGENTS AND STANDARDS

- Diphtheria antitoxin: 1st WHO International Standard for diphtheria antitoxin, equine (DI). or the 1st WHO International Standard for Diphtheria Antitoxin Human (10/262) are available from NIBSC (https://www.nibsc.org/science_and_research/bacteriology/diphtheria.aspx)
- In-house standards or a panel of positive control human serum samples of different level of defined activities in IU (e.g. 0.1, 0.01, 0.001 IU/ml) may be included to monitor assay performance;
- Diphtheria toxin: A purified preparation of diphtheria toxin of defined activity (minimum cytopathic dose) and stability should be used;
- Vero cells: Vero cells are available on request from Chief, Biologicals, WHO, Geneva, Switzerland. Cells may also be obtained from culture collections (ATCC, EDQM) or other sources provided that their robustness and sensitivity to diphtheria toxin is known.

##### 12.1.2 EQUIPMENT

- Laminar airflow cabinet
- Incubator (+36 to +38oC)
- Flat bottomed sterile 96 well tissue culture plates
- Multichannel manual or electronic pipettes 50-200 µl
- Sterile tips for pipettes
- Sterile serological pipettes (5-25 ml) and electronic pipette controller
- Haemocytometer (cell counting chamber) with Neubauer rulings or Burker counting chamber for cell counts
- Microscope
- Tissue culture flasks, 75 cm2 (or 150 cm2)
- Polyester pressure-sensitive film or microtiter sealing tapes
- pH indicator paper (optional if metabolic activity used as an end-point)

##### 12.1.3 BUFFERS AND REAGENTS

- Minimal Essential Medium (MEM) (commercially available from suitable suppliers)
- Fetal or newborn calf serum (must be confirmed free from diphtheria antitoxin)
- Antibiotic solution containing penicillin (10,000 IU/ml) and streptomycin (10 mg/ml)
- 200 mM L-glutamine solution
- 10% D (+)-Glucose solution
- 1 M HEPES buffer
- Hanks' balanced salt solution (HBSS) or PBS
- 0.25% trypsin/EDTA solution
- Tetrazolium dye 3-(4,5-dimethylthiazol-2-yl)-2,5-diphenyl tetrazolium bromide, MTT
- Sodium lauryl sulfate, SDS
- N,N-dimethylformamide, DMF

##### 12.1.4 COMPLETE CULTURE MEDIUM FOR VERO CELLS

Supplement MEM with:

- calf serum (final concentration 5-10% v/v)
- L-glutamine (2 mM)
- D-glucose (0.1% w/v)
- HEPES (0.01 M)
- penicillin (100 U/ml)
- streptomycin (100 µg/ml)

Other preparations of cell culture medium may also be suitable for use.

- Trypan blue (0.4%) solution

**Note: Medium and all the solutions have to be sterile.**

If MTT dye is used for staining viable cells, extraction buffer is prepared with solution of sodium lauryl sulfate (SDS, 10% w/v) in dimethylformamide (DMF, 50% v/v, pH to 4.7). Alternatively, other staining reagents may be used (e.g. crystal violet staining solution) prepared with 5 g of crystal violet dissolved in 100 ml of 37% formaldehyde, 200 ml abs. ethanol, 1665 ml distilled water, 35 ml 2 M Tris base and 10 g calcium chloride.

#### A12.2 PROCEDURES

All procedures are performed aseptically using the laminar airflow cabinet.

##### 12.2.1 CULTURE, HARVESTING AND COUNTING OF VERO CELLS

Established cultures of Vero cells can be maintained in 75 $cm^2$ tissue culture flasks in complete medium. Depending on the split ratio following passage and the percentage of serum in the medium, a confluent monolayer of cells is obtained after 4-6 days. The following procedure is suitable for routine passage and harvesting of Vero cell cultures. The cells are handled aseptically in the laminar airflow cabinet. Note that one T-75 tissue culture flask of Vero cells at 80-90% confluence will provide enough cells for 3 x 96-well tissue culture plates (at a cell density of 4 x $10^5$ cells/ml).

1. Remove the supernatant from a flask containing a confluent monolayer of Vero cells using a sterile pipette.
2. Add 1 ml of sterile HBSS (or PBS) solution to the flask rinse the cells and then remove using a sterile pipette.
3. Add 1 ml of sterile trypsin-EDTA solution to the flask and place in a 37 °C incubator until the cells are detached from the flask (2-5 minutes). The trypsin/EDTA solution should be pre-warmed to 37 °C to speed up the trypsinization process.
4. Add approximately 5 ml of complete medium to the flask to randomize the trypsin and resuspend the cell suspension using a sterile pipette to obtain a suspension of single cells for counting (gently mix the cell suspension within the sterile serological pipette to disperse cell clusters).
5. Prepare a 1 in 5 dilution of the cell suspension in 0.4% trypan blue solution and complete medium (e.g. 100 µl cell suspension + 100 µl 0.4% trypan blue solution + 300 µl complete medium). Depending on the total number of cells present a lower or higher dilution may be required. As a guide, the cell suspension should be diluted such that the total number of cells counted exceeds 100 (minimum required for statistical significance).
6. Prepare the haemocytometer by placing the coverslip over the mirrored counting surface. It may be necessary to moisten the edges of the chamber (this can be done by breathing on the glass) such that Newton's rings (rainbow-like interference patterns) appear indicating that the coverslip is in the correct position to allow accurate cell counting (the depth of the counting chamber is 0.1 mm).
7. Using a pipette, introduce a small sample (approximately 10 µl) of the diluted cell suspension into the counting chamber such that the mirrored surface is just covered. The chamber fills by capillary action. Fill both sides of the chamber to allow for counting in duplicate.
8. The entire grid on a standard haemocytometer is comprised of nine large squares (bounded by 3 lines), each of which has a surface area of 1 $mm^2$. The total volume of each large square is 1 x $10^{-4}$ $cm^3$ (0.0001 ml).
9. Count the number of cells in one large square and calculate the cell concentration as follows: *cells/ml = total cell count in one large square x* $10^4$.
10. For example, if 150 cells are counted in one large square (1 $mm^2$), the concentration of the cell suspension = 150 x $10^4$ cells/ml. If fewer than 100 cells are counted in 1 large square it may be necessary to count multiple large squares (for example, the 4 corner squares plus the centre square) and divide the total cell count by the total number of large squares used for counting.
11. For the Vero cell assay, prepare a cell suspension containing approximately 4 x $10^5$ cells/ml in complete medium. Note that the cell suspension should be prepared immediately before use and after all dilutions and neutralization steps have been performed.
12. To maintain the culture of Vero cells, seed approximately 1 x 106 cells into a new 75 $cm^2$ tissue culture flask and add 10 – 15 ml of complete medium prior to incubation at 37 °C.

#### 12.2.2 DETERMINATION OF THE TEST DOSE OF DIPHTHERIA TOXIN

The protocol described here is performed using the Lcd/1000 level of toxin defined by the lowest concentration of toxin (in Lf/ml) which when mixed with 0.001 IU/ml of antitoxin is capable of causing cytotoxic effects on Vero cells after 6 days of culture. At this toxin dose level, the sensitivity of the assay is approximately 0.002 IU/ml for the DI equine antitoxin. The sensitivity of Vero cells to diphtheria toxin may vary when different batches of cells and/or serum are used. As a result, the Lcd/1000 should be determined by each individual laboratory or whenever one of these variables is changed. These parameters should also be confirmed for every new lot of diphtheria toxin and reference antitoxin used. The test dose of toxin is determined by titration of a stable, purified diphtheria toxin against a suitable reference antitoxin as follows:

1. In a sterile 96-well tissue culture plate, fill all the wells of columns 2–11 with 50 µl of complete medium using a multichannel micropipette.
2. Dilute the diphtheria toxin in complete medium to give a starting concentration of approximately 0.02 Lf/ml.

3. Add 100 µl of the diluted diphtheria toxin solution to each well in column 1 using a micropipette.
4. Prepare serial twofold dilutions in 50 µl volumes starting at column 1 through to column 11 using a multichannel micropipette. Discard 50 µl from column 11.
5. Prepare a dilution of the reference diphtheria antitoxin in complete medium to give a diphtheria antitoxin concentration of 0.001 IU/ml.
6. Add 50 µl of the diluted diphtheria antitoxin preparation to all wells in columns 1-11.
7. Add 100 µl of complete medium to 4 "cell control" wells in column 12.
8. Add 50 µl of complete medium and 50 µl of diluted diphtheria antitoxin to 4 "antitoxin control" wells in column 12.
9. Allow the plate to stand at room temperature for 1 hour to allow toxin neutralization to occur.
10. Prepare a suspension of Vero cells in complete medium containing approximately 4 x 105 cells/ml (as described previously).
11. Add 50 µl of the cell suspension to all wells of the microplate. The total volume in all wells is 150 µl.
12. Shake the plates gently and cover with plate sealers to prevent the exchange of gas between medium and air. Note that the use of pressure film to seal plates is an important step for methods based on colour changes in the culture medium to determine assay end-points.
13. Incubate for 6 days at 37 °C in 5% CO2 incubator.
14. Perform staining of Vero cells (see example below) or follow an alternative detection method.

The Lcd/1000 dose of diphtheria toxin is defined as the lowest concentration of toxin causing more than 50% cytotoxicity in Vero cells in the presence of 0.001 IU/ml diphtheria antitoxin.

**Note that the minimum cytotoxic dose (MCD) of diphtheria toxin can be determined using the same procedure but without the addition of diphtheria antitoxin. To determine the MCD, 50 µl of complete medium should be added in step 6 instead of the diluted diphtheria antitoxin. The antitoxin control wells in column 12 should contain complete medium only and become blank control wells. Because this titration is performed without diphtheria antitoxin, it may be necessary to use a lower starting concentration of diphtheria toxin to determine the MCD.**

##### 12.2.3 STAINING OF VERO CELLS

The MTT is reduced in metabolically active, viable cells to form insoluble purple formazan crystals which are then randomized by the addition of detergent or solvent:

1. After 6 days of incubation at 37 °C, remove the plate sealer and check the wells for microbial contamination.
2. Prepare a solution of MTT in PBS (5 mg/ml). Sterilize by passing through a 0.2 µM syringe filter. Add 10 µl of the sterile MTT solution to each well of the microplate using a multichannel micropipette.
3. Return the microplate to the 37 °C incubator for 4 h to allow metabolism of the MTT by viable cells and formation of the blue formazan product.
4. Carefully remove the medium from all wells using a multichannel micropipette (set to >160 µl).
5. Add 100 µl of extraction buffer to all wells and return the microplate to the 37 °C incubator and leave overnight to allow extraction and solubilization of the formazan product.
6. Alternatively, monolayers of Vero cells can be stained for 5 minutes with crystal violet solution dissolved in formalin-ethanol. After staining the solution is discarded and wells rinsed with hot running tap water.

7. The plates are examined visually and the absorbance could be measured at 550-570 nm on a microplate reader. Once staining is complete, the colour is extremely stable at room temperature.

The presence of a dark blue colour indicates viable cells in both methods. A light blue colour indicates partial toxicity, while the absence of colour indicates complete toxicity and cell death.

#### 12.2.4 DETERMINATION OF POTENCY OF THE ANTITOXIN

Serial twofold dilutions of test and reference serum sample are prepared in complete medium in a 96-well tissue culture microplate. After the addition of the test dose of diphtheria toxin (defined in Lcd/1000 and previously determined), the mixtures are incubated at room temperature for 1 h to allow toxin randomization to occur. Vero cells are then added, and the plates incubated for 6 days. After 6 days of culture, the MTT assay or cell staining is performed to determine assay end-points. Reference antitoxin is included on each plate to calculate activity in IU/ml. As an internal control, antitoxin samples of known defined low/high activity may be titrated within each assay.

1. Fill all the wells of columns 2–11 with 50 µl of complete medium using a multichannel micropipette.
2. Fill the first four wells in column 12 (12A–12D) with 100 µl of complete medium using a multichannel micropipette ("cell control").
3. Fill the last four wells in column 12 (12E–12H) with 50 µl of complete medium using a multichannel micropipette ("toxin control").
4. Add 100 µl of each test serum sample into the appropriate well in column 1 (sera from all animals should be randomized across the plates).
5. Prepare a suitable dilution of the reference antitoxin in complete medium (for assays performed at the Lcd/1000 dose level, a starting concentration of 0.064 IU/ml should be suitable). Add 100 µl of the diluted reference antitoxin to the appropriate well in column one in every plate.
6. Make a twofold dilution series in 50 µl volumes starting at column 1 through to column 11 using a multichannel micropipette. Discard 50 µl from column 11 to randomized volumes.
7. Prepare a dilution of the test toxin in complete medium as determined previously. Test dose of toxin is defined as Lcd/1000.
8. Add 50 µl of the diluted diphtheria toxin solution to all wells in columns 1-11 using a multichannel micropipette. Add 50 µl of the diluted diphtheria toxin solution to the last four wells in column 12 (12E – 12H, toxin control).
9. Mix antitoxin with toxin by gently shaking and cover the plate with a lid.
10. Incubate at room temperature (20–25 °C) for one hour to allow toxin neutralization to occur.
11. Meanwhile, prepare a Vero cell suspension in complete medium containing approximately 4 x 105 cells/ml (as described previously).
12. Add 50 µl of the cell suspension to all wells of the microplate. The total volume in all wells should be 150 µl.
13. Shake the plates gently and cover with plate sealers to prevent the exchange of gas between medium and air. Note that the use of pressure film to seal plates is an important step for methods based on colour changes in the culture medium to determine assay end-points.
14. Incubate for 6 days at 37 °C in 5% CO2 incubator.
15. Perform the MTT assay or cell staining as described previously.

#### A12.3 CALCULATION OF RESULTS

The end-point of each test and reference serum sample is defined as the last well showing neutralization of toxin which in turn can be defined as an optimal density (OD) value greater than the 50% control OD value (if OD recorded). The end-point is recorded as a score based on the dilution of the serum sample at the end-point.

**FIGURE A11 Example arrangement of microtiter plate layout for titration of sera using the Vero cells where only one reference sample is included.**

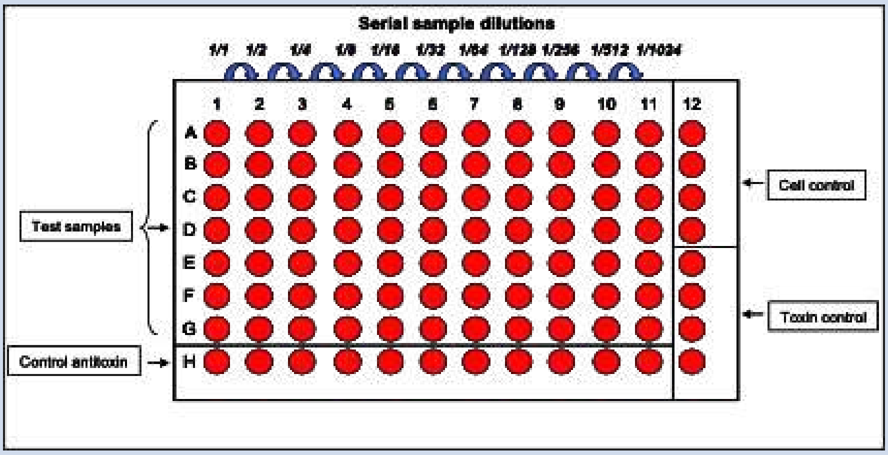

The plate format can be modified to include additional serum controls. The position of the reference (control) antitoxin should be randomized when multiple plates are used.

**FIGURE A12 Example of the Vero cell assay following the MTT extraction or staining**

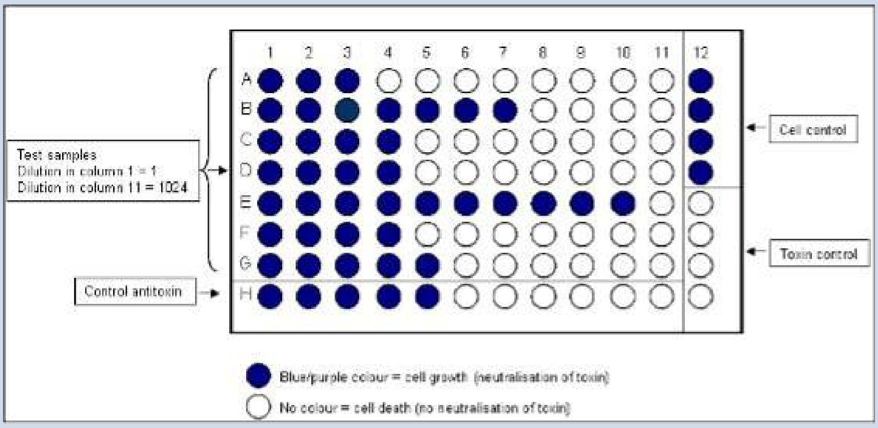

The end-point scores for individual test and reference serum samples should then be converted to titres in IU/ml by comparison with the end-point of the reference antitoxin on each plate. The antibody concentration of each serum under test can be calculated by multiplying the dilution ratio (sample titre end-point or score/standard titre end-point or score) with the calibrated concentration in IU of a reference standard.

**Note that any dilution of the serum samples prior to titration in the assay must be taken into account to obtain the final end-point titre in IU/ml.**

Note that any dilution of the serum samples prior to titration in the assay must be taken into account to obtain the final end-point titre in IU/ml.

#### A12.4 VALIDITY OF THE TEST

- The test is not valid if no toxicity is observed in the wells containing Vero cells and diphtheria toxin ("toxin control");
- The test is not valid if the wells containing Vero cells alone (in complete medium, "cell control") do not show positive cell growth with a confluent monolayer of cells after 6 days of culture. As a guide, the OD 570 nm after MTT staining should be >1;
- The end-point for the reference antitoxin should be 0.002 IU/ml for assays performed at the Lcd/1000 dose level;
- The results of the negative control serum must be below the limit of detection and calculated value for the two positive control sera (if used) are within the agreed limits so that the difference between the titres of the lowest and highest standard dilution is not more that 4-fold.

### STATISTICAL ANALYSIS PLAN

### VERSION 01.00, 12SEP2025

#### A Phase 1, Randomised, Single-Blind Clinical Study to Evaluate the Safety, Immunogenicity and Tolerability of SPVX02, a Tetanus and Diphtheria Booster Vaccine, Against Two Comparator Vaccines in Healthy Adult Participants

**SPL-SPVX02-01**

Prepared by: S-cubed Biometrics Ltd

For: Stablepharma Ltd

### 1. STATISTICAL ANALYSIS PLAN APPROVAL FORM

| | Signature (Date) |
|---|---|
| **Author(s):** | |
| Laura Grey<br>Statistics Manager<br>S-cubed Biometrics | Signer Name: Laura Grey<br>Signer Reason: I am the owner of this document<br>Signer Time: 12/Sep/2025 15:14:56<br>9d93d39b-8d10-4db9-a875-18ef874b950d |
| **Approval(s):** | |
| Karen O'Hanlon<br>Chief Development Officer<br>Stablepharma Ltd | Karen O'Hanlon<br>Signer Name: Karen O'Hanlon<br>Signer Reason: I approve this document<br>Signer Time: 16/Sep/2025 10:06:26<br>b16b2fe9-43b2-4c6e-b9e2-4b32efd99686 |
| Juana de la Torre Arrieta<br>Development Project Manager<br>Stablepharma Ltd | Juana de la Torre Arrieta<br>Signer Name: Juana De La Torre Arrieta<br>Signer Reason: I approve this document<br>Signer Time: 15/Sep/2025 09:09:41<br>3e51565f-d7bd-49c6-9f3e-f17090b53f05 |

#### 2. STATISTICAL ANALYSIS PLAN AUTHOR(S)

Laura Grey Msci MSC PhD
Statistics Manager
S-cubed Biometrics Ltd
99 Milton Park
Abingdon
Oxfordshire
OX14 4RY

### 3. LIST OF ABBREVIATIONS

| Abbreviation | Definition |
|---|---|
| AE | Adverse event |
| ALP | Alkaline Phosphatase |
| ALT | Alanine Aminotransferase |
| BDRM | Blinded Data Review Meeting |
| BLQ | Below the level of quantification |
| BMI | Body Mass Index |
| CI | Confidence Interval |
| CRF | Case Report Form |
| CS | Clinically Significant |
| CSR | Clinical Study Report |
| ECG | Electrocardiogram |
| FAS | Full Analysis Set |
| GM | Geometric Mean |
| GMT | Geometric Mean Titres |
| GSD | Geometric Standard Deviation |
| HCT | Haematocrit |
| HIV | Human Immunodeficiency Virus |
| IC | Informed Consent |
| LLOD | Lower Limit of Detection |
| LLOQ | Lower Limit of Quantification |
| LSC | Local Safety Committee |
| MCV | Mean Corpuscular Volume |
| MCH | Mean Corpuscular Haemoglobin |
| MedDRA | Medical dictionary for regulatory activities |

| Abbreviation | Definition |
|---|---|
| NCS | Not Clinically Significant |
| PI | Principal Investigator |
| PT | Preferred Term |
| RBC | Red Blood Cells |
| SAE | Serious adverse event |
| SAP | Statistical Analysis Plan |
| SAS | Safety Analysis Set |
| SD | Standard deviation |
| SOC | System Organ Class |
| TE | Treatment emergent |
| TFL | Tables Figures and Listings |
| WBC | White Blood Cells |
| WHO | World Health Organisation |
| WOCBP | Women of Child-Bearing Potential |

### 4. INTRODUCTION

This statistical analysis plan (SAP) explains in detail the statistical analyses that will be performed for the Stablepharma study SPL-SPVX02-01. The analysis is outlined within the study protocol (version 3.0 dated 22 May 2025), and this SAP contains a more technical and detailed description of those analyses. In particular, information is provided on the definitions of the participant analysis sets, and it also details the list of Tables, Figures and Listings (TFL) that will be produced by S-cubed Biometrics for use and inclusion within the Clinical Study Report (CSR). The SAP has been written and finalised before the database is locked and before provision of any unblinded outputs.

Any deviations from the protocol specified analysis are noted in Section 11. These along with any deviations from the analyses stated within this SAP will be described within the CSR.

### 5. STUDY OBJECTIVES

The objectives of the study are:

Primary:

- Evaluate the safety of a single dose of SPVX02
- Evaluate the tolerability of a single vaccination of SPVX02, Tetadif® or diTeBooster®

Secondary:

- To evaluate the seroprotection rates observed in sera collected from participants on Day 28 post-dose, following administration of a single dose of SPVX02, Tetadif® or diTeBooster®.
- To evaluate the post vaccination Geometric Mean Titres (GMT) of anti-TT and anti-DT antibodies observed in sera collected from participants at Day 28 post-dose, following administration of a single dose of SPVX02, Tetadif® or diTeBooster®.
- To evaluate the longer-term seroprotection rates observed in sera collected from participants on Day 28 post-dose, following administration of a single dose of SPVX02, Tetadif® or diTeBooster®.

Exploratory:

- To determine the post-vaccination geometric mean titres (GMT) of anti-DT neutralising antibodies observed in sera collected from participants on Day 28 post-dose, following administration of a single dose of SPVX02, Tetadif® or diTeBooster®.

### 6. STUDY DESIGN

#### 6.1. Summary of Study Design

This is a Phase 1, first-in-human study of SPVX02. This single-blind study will be conducted in 60 healthy adult participants randomised 1:1:1 to receive a single dose of SPVX02, Tetadif® or diTeBooster®. The primary objective is to evaluate safety and tolerability of SPVX02, while the secondary objective includes assessing its immunogenicity. Observations from Tetadif® and diTeBooster® will provide comparative context for these assessments.

The study consists of the following:

- A 42-day screening period before the start of the study intervention and follow up period.
- A study intervention and follow up period of 28 days, consisting of dosing on Day 1, a follow-up telephone call on Day 2, and follow-up visits on Days 7 and 28.
- End of study procedures will be performed at the final visit on Day 28.

In addition, the data from Day 1 to Day 7 (including diary card data) from the first 3 participants (sentinel group) dosed in the study will be reviewed by the Local Safety Committee at Southampton Clinical Research Facility chaired by Prof Chris Edwards BSc, MBBS, MD, FRCP.

The first data review will occur as soon as is reasonably possible after the first 3 participants in the sentinel group have received a single dose of IMP.

This sentinel group of 3 participants will be randomised as follows:

- 2 participants are randomised to receive SPVX02
- 1 participant is randomised to receive Tetadif®
- 0 participants to receive diTeBooster®

The remaining 57 participants in the study will not be dosed until the Local Safety Committee has given its written instruction to proceed.

#### 6.2. Randomisation / Intervention Allocation and Blinding

At the screening visit, participants will be sequentially allocated a participant number (participant ID) upon providing written informed consent. This number will be used to identify the participant throughout the study. The participant number will be 5 digits (SS-NNN) consisting of a two-digit site number (SS) and a three-digit participant number (NNN) (e.g. 001, 002, 003, etc).

The randomisation schedule, generated prior to the study by S-cubed, ensures balanced allocation of participants to the three intervention groups.

- On Day 1, participants will be assigned a unique randomisation number in ascending numerical order. This number encodes the participant's intervention allocation and will be recorded in the study database. The Randomisation number will be a 3 digit number RNN, where R is a one-digit replacement number starting from 0 and NN is a sequential number starting from 01.
- If a participant is withdrawn before receiving the study vaccine, they may be replaced following discussion with the Sponsor. Any replacement participants will be assigned a randomisation number with the next sequential replacement number (R) but the same sequential number (NN). Participants who discontinue after vaccination will not automatically be replaced unless determined necessary by the Sponsor.

- The site will dispense the assigned study intervention at the study visit.
- Study interventions will be labelled with the participant's unique participant number to maintain accurate tracking.
- Once randomisation numbers have been assigned, no attempt will be made to use those numbers again. If a randomisation number is allocated incorrectly, no attempt will be made to remedy the error once the study vaccine has been dispensed.

A sentinel group of 3 participants will be randomised on a ratio of 2:1:0, before the remaining participants are randomised to achieve an overall 1:1:1 ratio.

#### 6.3. Blinding

This is a single-blind study in which participants are blinded to the specific study intervention they receive. The team performing the post-dose assessments, and the sponsor will also remain blinded to intervention allocation. The pharmacy team will be unblinded as will the clinical team performing the study drug administrations. Participants will be randomly assigned in a 1:1:1 ratio to receive SPVX02, Tetadif® or diTeBooster®.

Participants will be randomised on Day 1 prior to dosing where each participant will receive a unique randomisation number. Participants will then receive the corresponding study intervention according to the randomisation scheme. To ensure the number of unblinded personnel is kept to a minimum, the randomisation schedule will only be available to Pharmacy staff. The actual intervention administered should be recorded in the source documentation but not recorded in the eCRF. Blinded personnel will perform key safety and immunogenicity assessments.

#### 6.4. Study Procedures Flow Chart

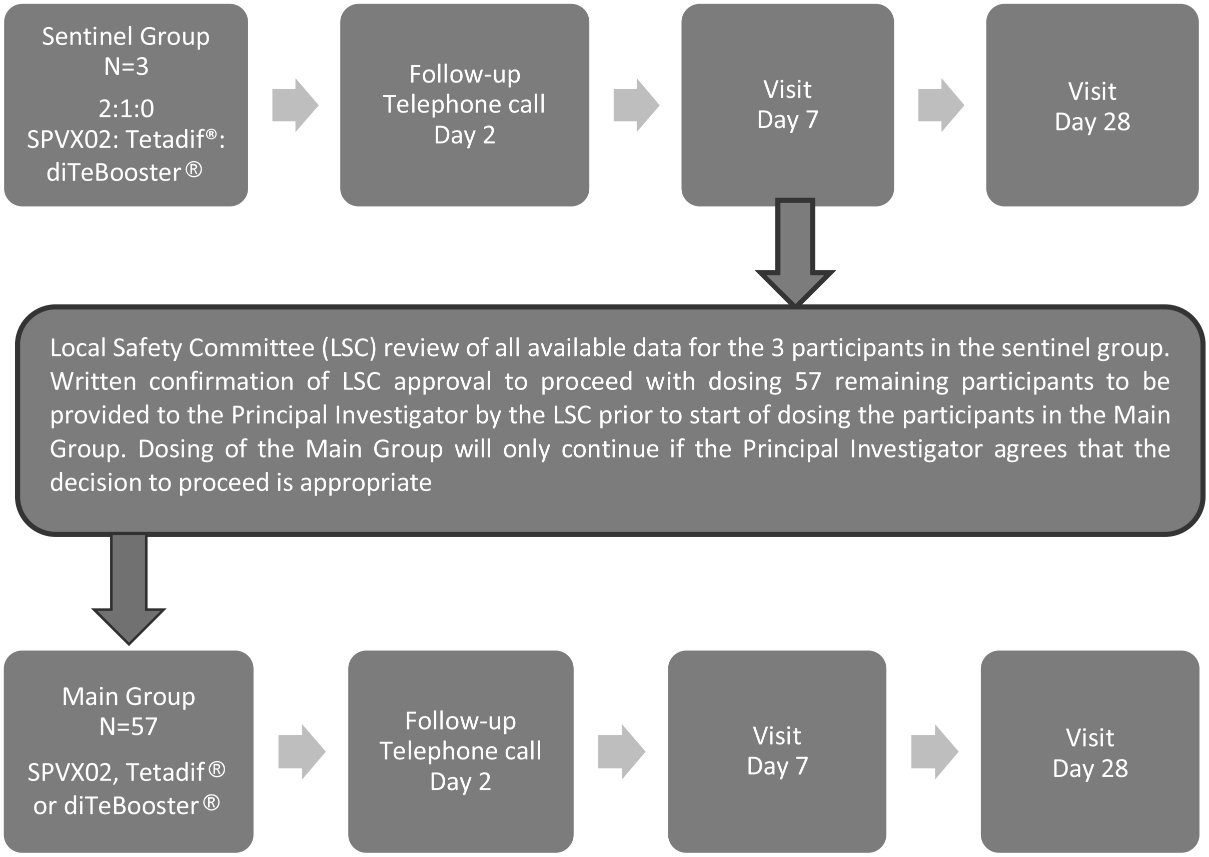

*Abbreviations: N= number of participants*

*Overall, participants will be randomized 1:1:1 for SPVX02:Tetadif®:diTeBooster®*

For all participants, all available data relating to safety, immunogenicity, tolerability (including AE and SAE data) will be reviewed on an ongoing basis by the Sponsor's Medical Monitor and the Principal Investigator at the study site. The Sponsor's Medical Monitor and the Principal Investigator will make recommendations concerning the continuation, modification or termination of the study if one of the study stopping criteria defined in Section 9.2 of the protocol is met.. If the study is halted following triggering of a stopping rule, it will only be restarted following approval of a substantial amendment by the regulatory authorities.

In addition, further study oversight will be provided by a designated Local Safety Committee.

#### 6.5. Schedule of Activities

**Table 1: Schedule of Activities**

| Procedures and Assessments | | | | | | |
|---|---|---|---|---|---|---|
| **Study Period** | **Screening** | **Intervention Period** | | | | **ET[j]** |
| **Visit** | **1** | **2** | **3** | **4** | **5** | |
| **Day** | **-42 to 0** | **1** | **2** | **7 (± 3 days)** | **28 (± 3 days)** | |
| Informed consent | X | | | | | |
| Review of eligibility criteria by the PI[b] | X | X[c] | | | | |
| Demographics and baseline characteristics | X | | | | | |
| Full physical examination[d] | X | X | | X[h] | X[h] | |
| Height and weight | X | | | | | |
| Medical history | X | | | | | |
| Clinical laboratory tests (biochemistry, haematology, coagulation, thyroid function) | X | X | | | X | X |
| Urinalysis | X | X | | | X | X |
| Pregnancy test (WOCBP only) | X | X | | | | |
| Drug and alcohol screen | X | X | | | | |
| HIV, Hepatitis B and C screen | X | | | | | |
| 12-lead ECG[e] | X[f] | | | | | |
| Vital signs | X | X | | X[h] | X[h] | |
| Randomisation | | X | | | | |
| Dosing | | X | | | | |
| AE review[g] | X | X | X | X | X | X |
| Concomitant medications | X | X | X | X | X | X |
| Blood samples for tetanus and diphtheria antibody analysis | X | X | | | X | X |
| Blood samples for microneutralisation assay | | X | | | X | |
| Local and systemic reactogenicity daily evaluation (pain, redness, swelling, etc.) – e-diary (for first 7 days post dose only) | | X[i] | X[i] | X[i] | | |
| Telephone call to assess concomitant medications and AEs | | | X | | | |

a In the case of a participant withdrawing from a study (between intervention and follow up (Day 1 to Day 28)), or being withdrawn by the investigator, a written confirmation of the withdrawn will be provided and signed by the investigator. Criteria for withdrawal of participants from the study are described in the protocol.

b Eligibility must be confirmed by a qualified medical doctor who has received relevant training in this study and is entered on the study delegation log

c Eligibility must be re-confirmed at randomisation prior to first dose of investigational intervention

[d] Full physical examination.
[e] 12-lead ECGs will be obtained.
[f] At screening, a 12-lead ECGs will be obtained
[g] AE and concomitant medication review will be carried out throughout the study from informed consent to EoS.
[h] To be symptom directed and performed at the discretion of the Investigator
[i] e-Diaries are recorded by participants each day (day 1 to 7)
[j] Further details for the Early Termination are described in the protocol.

AE=adverse event; ECG=electrocardiogram; ET=early termination; HIV=human immunodeficiency virus; EoS=end of study; PI=principal investigator; WOCBP=women of childbearing potential.

#### 6.6. Interim Analysis / Data Monitoring

There will be no interim analysis for this study. The safety of the sentinel group will be assessed prior to proceeding with the remainder of the study. Additionally a Local Safety Committee (LSC) may monitor the data for the study. This will be done in a blinded manner unless unblinding is necessary, in which case the relevant outputs will be restricted to a small unblinded team.

### 7. STUDY ENDPOINTS

#### 7.1. Primary Safety Endpoints

Incidence of safety and reactogenicity events which include:

- Adverse events
- Serious adverse events
  - Incidence of local and systemic reactogenicity events observed for 7 days post-dose which include: pain, induration/swelling, tenderness, warmth, erythema at the vaccination site plus feverishness, chills, myalgia (muscle ache) , fatigue, headache, arthralgia (joint pain), (skin) rash.

#### 7.2. Secondary Endpoints

Secondary endpoints will include:

- Incidence of seroprotection resulting from a single dose of SPVX02, Tetadif® or diTeBooster® at 28 days post-dose.
- Evaluation of GMT of anti-TT and anti-DT antibodies resulting from a single dose of SPVX02, Tetadif® or diTeBooster® at 28 days post-dose.
- Incidence of longer term seroprotection resulting from a single dose of SPVX02, Tetadif® or diTeBooster® at 28 days post-dose.

#### 7.3. Exploratory Endpoints

- Determination of geometric mean neutralising antibody titres (GMT) for diphtheria resulting from a single vaccination with SPVX02, Tetadif® or diTeBooster® at 28 days post-dose.

Exploratory endpoints will not be described within this SAP.

### 8. SAMPLE SIZE

The protocol specified sample size section is as follows:

As there is no formal hypothesis testing for this study there has been no formal sample size estimation. However, 20 participants in each of the 3 intervention groups is judged to be sufficient so as to meet the study objectives.

### 9. STUDY ANALYSIS SETS

Analysis sets defined below will be reviewed against the study database at a blinded data review meeting (BDRM). The database at this time will be nearly final (i.e. meeting may result in further data queries/changes post meeting), so participant inclusion/exclusion from analysis sets defined at this meeting, will be further checked (post meeting) against a locked database, and will then be finalised prior to unblinding the study.

| Analysis Sets | Description |
|---|---|
| Full Analysis Set (FAS) | All randomised participants. |
| Safety Analysis Set (SAS) | All randomised participants who were exposed to an investigational intervention. |
| Immunogenicity Analysis Set | All randomised participants who were exposed to an investigational intervention and have at least one baseline and one corresponding post-baseline immunogenicity measurement and have no protocol deviations that may impact their immunogenicity results. |

### 10. PLANNED STATISTICAL METHODS

#### 10.1. Statistical Considerations

##### *10.1.1. General Definitions*

In all applicable summary/analysis presentations of safety endpoints, Baseline is defined as the last non-missing assessment value for a participant, for that particular parameter, that is prior to dosing with study intervention, unless over-ruled at the BDRM or otherwise stated in the appropriate endpoint sections below.

Within summary presentations/analyses it is envisaged that only scheduled protocol visit values will be used for post-baseline time points. In the clinical database a number of data points have been labelled as unscheduled/additional recordings of data. These data points will be included within participant listings only. However, at the BDRM the occurrence of such unscheduled data will be reviewed to decide if (and how) any such data points should be included within summary presentations/analyses. Any such decisions will be documented in the BDRM minutes.

##### *10.1.2. Data Presentation*

The full list of TFLs to be produced for the final study analysis are shown in Section 13, and the specific format and content of each data Table/Listing presentation is shown in Section 14.

All summaries will be presented by intervention group. Selected presentations will include an overall column which will include all three intervention groups.

Within all Tables and Figures the interventions will be labelled as in Table 2, while the abbreviated labels may be used in the listings if required. Listings will be presented by intervention group and participant number.

**Table 2: Intervention Group Labels**

| Label | Abbreviated Labels |
|---|---|
| SPVX02 | SPVX |
| Tetadif | Teta |
| diTeBooster | diTe |

The scheduled protocol visits will be labelled in tables and figures as follows:

- Baseline
- Day 1
- Day 7
- Day 28

The baseline definition may vary by endpoint, as stated in Section 10.1.1 and the specific endpoint sections. Day 1 is the day that the intervention is administered following baseline safety measurements.

An electronic reactogenicity diary is completed every day following intervention from Day 1 to Day 7 (inclusive), the visits labels for the data collected within the diary will be presented in relevant summary tables as Day 1, Day 2, Day 3, Day 4, Day 5, Day 6 and Day 7.

Listings will include all relevant data collected within the study database, with the visit listed as recorded.

Unscheduled visit data will be labelled as "Unscheduled" together with a date in the relevant data Listings. Unscheduled data will not be used in the summary tables unless otherwise agreed at the BDRM.

Where duplicate information is collected in both the database and in the vendor data transfer(s) (e.g., sampling date and time) this information will be reconciled by data management and the information from the database will be included in participant Listings.

All variables will be listed to the same number of decimal places as reported. Descriptive statistics for all endpoints that are continuous data will have the following summary statistics presented in the following order: n, mean (rounded to one more decimal place than recorded), standard deviation (rounded to two more decimal places than recorded), median (rounded to one more decimal place than recorded), lower quartile (rounded to one more decimal place than recorded), upper quartile (rounded to one more decimal place than recorded), minimum (as recorded), and maximum (as recorded).

Note: for endpoint(s) that require a geometric mean to be produced, and those endpoint(s) can have raw values of 0 (zero), the geometric mean calculation will add an appropriate constant value to all raw values prior to logging and will subtract that constant value from the final calculated anti-logged mean. The constant value used will be documented in the footnote of the tables. An example of such a calculation is shown below:

$$GM = \left( 10^{\left[\frac{\sum_{i=1}^{N} log_{10}(x_i+1)}{N}\right]} \right) - 1$$

Categorical variables will be summarised using proportions (counts and percentages). All percentages in summary tables will be calculated using as the denominator, either the participant analysis set or the number of non-missing observations. The specific approach is detailed within each (relevant) table template (Section 14).

Unless otherwise stated in the appropriate endpoint section(s) below, laboratory parameter values that are <X, below the level of quantification (BLQ), or less than the lower limit of quantification

(<LLOQ) will be set to zero in computations for summary presentations and analysis but will be noted as collected in Listings.

##### *10.1.3. Statistical Testing and Estimation*

There will be no statistical testing in this study. Where appropriate 95% confidence intervals may be presented.

##### *10.1.4. Handling of Dropouts or Missing Data*

In general, missing data will not be imputed and all summary statistics will be reported based upon observed data. For a limited number of summary presentations, missing data rules may be introduced. In particular, methods of handling incomplete dates for adverse events (Section 10.9.1) and incomplete dates for concomitant medications (Section 10.5) are presented. Any other data with partial dates, which are required for use in time related calculations, will be completed using a suitably conservative approach. Dates will be shown in Listings as they have been recorded.

##### *10.1.5. Subgroups*

For both diphtheria and tetanus the seroprotection status at baseline will be derived for each participant. A participant was seroprotected at baseline if their IgG serum antibody titre was ≥ 0.1 IU/mL.

Summaries may be presented by seroprotection status at baseline, or for only the subgroup of participants who were not seroprotected at baseline. Similarly summaries may be presented by whether a participant had achieved longer-term seroprotection at baseline or not, where this is derived for both diphtheria and tetanus if the IgG serum antibody was > 1.0 IU/mL.

Other subgroups that may be used include:

- Sex
    - Male
    - Female
- BMI Group
    - <18.5 – Underweight
    - 18.5 – 24.9: Normal Weight
    - 25.0 – 29.9: Overweight
    - 30.0 – 34.9: Obese Class I
    - 35.0 – 39.9: Obese Class II
    - >= 40.0: Obese Class III
- Age Group
    - <=25
    - 26-35
    - 36-45
    - >45

Age groups or BMI groups with less than 2 participants over the three intervention groups may be merged.

##### *10.1.6. Multiple Comparison/Multiplicity*

As there is no statistical testing no adjustment for multiple comparisons is required.

##### *10.1.7. Software*

Data will be reported using SAS (version 9.4 or later).

#### 10.2. Participant Disposition

The number of participants enrolled (non-screen failure), the number randomised, the number withdrawing (also split by reason for withdrawal) from and completing the study, and the numbers in each analysis set will be summarised by intervention group. The data on participant disposition, informed consent and analysis set membership will also be listed.

#### 10.3. Protocol Deviations

Protocol deviations will be listed using information collected on the CRF and will be described appropriately within the CSR. Important protocol deviations (i.e. those that may impact the analysis) will be discussed at the BDRM.

#### 10.4. Demographic and Other Baseline Characteristics

Unless otherwise described, the Safety Analysis Set will be used in summaries of demographic and baseline data. No statistical testing will be used to compare interventions for different baseline characteristics.

##### *10.4.1. Demographics*

Demographic variables at informed consent (sex, age, race, ethnicity, height (cm), weight (kg), body mass index (kg/m$^2$) and child-bearing potential) will be summarised across all participants.

##### *10.4.2. Child-Bearing Potential*

Child-bearing potential and contraceptive use will be listed.

##### *10.4.3. Medical History*

Past and present significant medical history data will be coded using MedDRA version 28.0. Ongoing medical history, identified as those conditions of medical history that were ongoing at screening, will be summarised by MedDRA system organ class (SOC) and preferred term by intervention group.

#### 10.5. Prior and Concomitant Medications

All medication terms will be coded using the World Health Organisation (WHO) Drug Dictionary Enhanced (WHO Drug Global, version March 2025).

Medications will be assigned as being prior to or concomitant with study intervention, based on the start and stop dates of the medication and the date of study intervention. If the medication stop date is before the date of study intervention, the medication will be assigned as being prior to study intervention. Medications that are ongoing at administration of study intervention or started after first administration will be deemed to be concomitant medications. If medication dates are incomplete and it is not clear whether the medication was concomitant, it will be assumed to be concomitant.

All concomitant medications will be summarised by ATC class (Level 2) and preferred base name (WHO Code 01001) by intervention group. If a participant has separate periods of taking specific medications, then that medication is only counted once within the specific period of observation (i.e. prior or concomitant) where it is taken. All medications will be listed, where concomitant medications will be flagged.

#### 10.6. Study Drug Administration

Details of study intervention administration, including date, time and intervention received will be listed.

#### 10.7. Immunogenicity Analysis

All immunogenicity analyses will be performed on the Immunogenicity Analysis Set and participants will be summarised/analysed by the actual intervention.

Any antibody results that are below the lower limit of quantification (LLOQ) will be reported as measured, while the results below the lower limit of detection (LLOD) will be reported as <LLOD. Any results <LLOD will be replaced with half of the LLOD before summarising. Values above the LLOD will be summarised as reported.

All antibody results will be listed as reported, with any relevant limits (LLOD and/or LLOQ) highlighted.

##### *10.7.1. Seroprotection*

Seroprotection is defined as an IgG serum antibody titre of ≥ 0.1 IU/mL which is the antibody titre level of anti-TT and anti-DT antibodies that are considered to confer protection against tetanus[1] and diphtheria [2] infection.

The seroprotection rate for tetanus and diphtheria will be summarised by intervention group as the number and percentage of participants who meet the criteria for seroprotection at Day 28. The summary will also present the 95% Clopper-Pearson Confidence interval for each group.

Seroprotection rate will be presented similarly for the subgroup of participants who were not seroprotected at baseline, and who were not longer-term seroprotected at baseline.

To account for the possibility of participants already meeting the protocol defined seroprotection criteria at baseline additional definitions of seroprotection have been included to allow further comparison of the study interventions.

1) IgG serum antibody titre of >= 0.1 IU/mL and an increase from baseline of at least 2-fold.
2) IgG serum antibody titre of >= 0.1 IU/mL and an increase from baseline of at least 4-fold.

For both of these definitions the seroprotection rate for tetanus and diphtheria will be summarised as described above.

##### *10.7.2. Geometric Mean Antibody Titres*

The absolute antibody titres will be summarised, at both baseline and Day 28, by intervention group. The summary will include the geometric mean and standard deviation.

The summary will also include the geometric mean ratio of the Day 28 titre to the baseline titre. This will be derived by back-transforming the mean of the [log(titre at Day 28) – log(titre at Baseline)].

A box-scatterplot will be produced of the antibody titres, separately for tetanus and diphtheria. Each page will present the titres at baseline and at Day 28 for each of the 3 intervention groups. The y-axis will be presented on the log-scale.

Additionally summary tables and box-scatterplots will be presented by seroprotection status at baseline, longer-term seroprotection status at baseline, age group, sex and BMI group.

##### *10.7.3. Longer Term Seroprotection*

Longer-term seroprotection is defined as an IgG serum antibody titre of >= 1.0 IU/mL which is the antibody titre level of anti-TT and anti-DT antibodies that are considered to confer longer-term protection against tetanus[1] and diphtheria [2] infection.

The longer-term seroprotection rate for tetanus and diphtheria will be summarised by intervention group as the number and percentage of participants who meet the criteria at Day 28. The summary will also present the 95% Clopper-Pearson Confidence interval for each group.

Longer-term seroprotection rate will be described similarly for the subgroup of participants who were not seroprotected at baseline and those who were not longer-term seroprotected at baseline.

Additional definitions have also been added for longer-term seroprotection:

1) IgG serum antibody titre of >=1.0 IU/mL and an increase from baseline of at least 2-fold.
2) IgG serum antibody titre of >=1.0 IU/mL and an increase from baseline of at least 4-fold.

For both of these definitions the seroprotection rate for tetanus and diphtheria will be summarised as described above.

#### 10.8. Safety Analysis

The SAS will be used for all safety presentations. All analyses of safety endpoints will be descriptive and no statistical analysis of safety data will be performed. Any presentations of safety data by intervention group will be presented by actual intervention.

##### *10.8.1. Reactogenicity Events*

An e-diary will be completed by participants every evening from Day 1 to Day 7 to assess the tolerability of the study interventions. The diary will collect information on symptoms including temperature, reactions local to the injection site (pain, induration/swelling, tenderness, warmth, erythema/redness) and systemic reactions (feverishness, chills, muscle ache, fatigue, headache, joint pain, skin rash).

Where a participant has reported a symptom (i.e. answered yes to having experienced the symptom) a grade will be derived using the information from the e-diary using Table 3. Any temperature below 38.0 $^{O}$C will be classified as not having a fever.

**Table 3: Severity Grading for Reactogenicity Events**

| | Mild (Grade 1) | Moderate (Grade 2) | Severe (Grade 3) | Potentially Life Threatening (Grade 4) |
|---|---|---|---|---|
| Fever ($^{O}$C) | 38.0-38.4 | 38.5-38.9 | 39.0-40.0 | >40.0 |
| Erythema/Redness (cm) | 2.5-5.0 | 5.1-10.0 | >10.0 | Necrosis or exfoliative dermatitis |
| Induration/Swelling (cm) | 2.5 - 5.0 and no interference | 5.1 - 10.0 or some interference | >10.0 or significant interference | Necrosis |
| Pain | no interference | Repeated use of pain relief > 24 hour or some interference | significant interference | ER visit or hospitalisation |
| Warmth | no interference | some interference | significant interference | ER visit or hospitalisation |
| Tenderness | Mild discomfort to touch | Discomfort with movement | Significant discomfort at rest | ER visit or hospitalisation |
| Feverishness | no interference | some interference | significant interference | ER visit or hospitalisation |
| Chills | no interference | some interference | significant interference | ER visit or hospitalisation |
| Muscle Ache | no interference | some interference | significant interference | ER visit or hospitalisation |
| Fatigue | no interference | some interference | significant interference | ER visit or hospitalisation |
| Headache | no interference | Repeated use of pain relief > 24 hour or some interference | significant interference | ER visit or hospitalisation |
| Joint Pain | no interference | some interference | significant interference | ER visit or hospitalisation |
| Skin Rash | no interference | some interference | significant interference | ER visit or hospitalisation |

The number and percentage of participants with each symptom, or any symptom, will be summarised by intervention group and study day, as well as the number and percentage of participants who experience the symptom at any time over the 7 days. Additionally the summary will be presented by severity. In the summary by severity the ‘any time’ and ‘any symptom’ summary will present the worst severity of the symptoms.

##### *10.8.2. Adverse Events*

All unsolicited adverse events will be coded using MedDRA, Version 28.0. This may include adverse events identified through the symptom e-diaries.

An adverse event is defined as treatment-emergent if the onset date is on or after the date of first dose of study intervention. Should any onset date for an adverse event be missing or only a partial date recorded (such that it cannot be determined if the event onset was prior to the date of first dose) then it will be assumed that the event is treatment-emergent, unless the adverse event stop date indicates otherwise. Any adverse event with an onset date earlier than the date of first dose of study intervention will be classified as a pre-treatment adverse event and will be identified in participant listings only.

If a participant experiences more than one AE with the same preferred term, that preferred term will be counted only once. It will be assigned the greatest observed severity and the strongest relationship to study intervention among those events for the tables in which those characteristics are summarised.

Treatment-emergent AEs (participant incidence and number of events) will be summarised by:

- Intervention group, system organ class, and preferred term;
- Intervention group, system organ class, preferred term and severity;

Additionally serious treatment-emergent AEs (participant incidence) will be summarised by intervention group, system organ class, and preferred term, as well as related treatment-emergent adverse events where related includes any AE that has been recorded as possible, probable, unlikely, unknown or the relationship is missing.

In all AE summary tables results will be displayed ordered in terms of decreasing frequency of SOC occurrence (based on the total across intervention groups), and within each SOC also ordered in terms of decreasing frequency of preferred term occurrence (also based on the total across intervention groups). In all AE summary tables, in addition to presenting the information by intervention group, a total column (all three intervention groups combined) will also be presented.

##### *10.8.3. Laboratory Variables*

The following haematology, biochemistry, coagulation and thyroid parameters will be included within summary presentations (and presented in the units as shown):

- Haematology: Red blood cell (RBC) count (10^12/L), mean corpuscular haemoglobin (MCH) (pg), mean corpuscular volume (MCV) (fL), haematocrit (HCT) (%), haemoglobin (g/L), platelet count (10^9/L), white blood cell (WBC) count (absolute, 10^9/L), neutrophils (absolute, 10^9/L), lymphocytes (absolute, 10^9/L), monocytes (absolute, 10^9/L), eosinophils (absolute, 10^9/L), basophils (absolute, 10^9/L).

- Coagulation: Prothrombin time (s), Prothrombin Intl. normalised ratio (ratio), activated partial thromboplastin time (s).

- Thyroid Function: Thyroid stimulating hormone, Thyroxine, free.

- Biochemistry: Alanine aminotransferase (ALT) (IU/L), albumin (g/L), alkaline phosphatase (ALP) (IU/L), total bilirubin (umol/L), calcium (mmol/L), creatinine (umol/L), glucose (mmol/L), potassium (mmol/L), total protein (g/L), sodium (mmol/L), urea (mmol/L).

Laboratory data collected in different units to that shown will be converted to the above specified units (if possible) for presentation in tables and listings.

Laboratory values outside the reference range will be identified in the participant listings as above or below the reference range. Laboratory values that are below the level of quantification (BLQ) or <X will be set to zero in computations for summary presentations but will be noted as recorded (i.e. BLQ or <X) in participant listings. Laboratory values that are missing will remain missing and will not be presented in listings.

Observed parameter values and changes from baseline will be summarised for the above highlighted haematology, biochemistry, coagulation and thyroid parameters by intervention group at each protocol scheduled visit.

Unscheduled visit assessments will be included within the relevant participant listings.

Urinalysis results and, where available, microscopy results will be listed.

##### *10.8.4. Pregnancy*

The results of pregnancy tests, both urine and serum, carried out throughout the study will be listed for all females.

##### *10.8.5. Drugs of Abuse Screening*

The results of drugs of abuse screening will be listed.

##### *10.8.6. Viral Serology*

The results of viral serology screening (HBsAg, HCV and HIV) will be listed.

##### *10.8.7. Vital Signs*

All vital signs results will be listed.

##### *10.8.8. Electrocardiogram (ECG)*

The ECG results obtained at screening will be listed.

##### *10.8.9. Physical Examination*

Physical examination results will be listed.

### 11. CHANGES TO THE PROTOCOL SPECIFIED ANALYSIS

The following changes have been made in the SAP compared to the protocol specified analysis:

Additional seroprotection definitions have been included to account for the possibility of participants having a high antibody level prior to receiving the study vaccination.

Two subgroups, based on the baseline antibody levels (seroprotection and longer-term seroprotection) have also been added along with a number of presentations that will be displayed by these subgroups.

Additionally a number of presentations will be displayed by Age group, BMI group and Sex.

### 13. TABLES, FIGURES AND LISTINGS

#### 13.1. Specific Presentation Details

Tables, listings and figures will be provided in word and pdf format. All summary tables and figures will have source data footnotes that refer to the relevant listings. Dates will appear as DDMMMYYYY times as HH:MM. All listings will be ordered by Intervention group, participant number and date/time of visit. For the presentation of summary data, values will be aligned based on the unit column, and not left/right justified. For example:

```
 Parameter                           n                 XX
                                     Mean (SD)         XX.X (XX.XX)
                  [optional]         GM (GSD)          XX.X (XX.XX)
                  [optional]         CV (%)            XX.X
                                     Median            XX.X
                                     Q1, Q3            XX.X, XX.X
                                     Min, Max          XX, XX
```

All Tables, Listings and Figures will have the SAS program name and date of production in the footnote.

All Tables, Listings and Figures will include the following study header and footer:

```
CONFIDENTIAL                                 Stablepharma                 Page x of y
                                            SPL-SPVX02-01

                                              Table X.X
                                                Title
                                             Analysis Set

Source Data: Listing 16.2.x [Source data footnote only appears for tables and figures, where x references
relevant listing number]

Program: XXXXXXXXX                    Output: XXXXXXX                Date: XXXXXXXXX
```

#### 13.2. List of Tables

| Table Number | Table Title |
|---|---|
| 14.1.1 | Participant Disposition – All Participants |
| 14.1.2 | Demography – Safety Analysis Set |
| 14.1.3 | Medical History by System Organ Class and Preferred Term – Safety Analysis Set |
| 14.1.4 | Concomitant Medications – Safety Analysis Set |
| 14.2.1.1.1 | Seroprotection Rate – Diphtheria – Immunogenicity Analysis Set |
| 14.2.1.1.2 | Seroprotection Rate – Diphtheria - Participants Not Seroprotected at Baseline – Immunogenicity Analysis Set |
| 14.2.1.1.3 | Seroprotection Rate – Diphtheria - Participants Not Longer-Term Seroprotected at Baseline – Immunogenicity Analysis Set |
| 14.2.1.2.1 | Seroprotection Rate – Tetanus – Immunogenicity Analysis Set |
| 14.2.1.2.2 | Seroprotection Rate – Tetanus - Participants Not Seroprotected at Baseline – Immunogenicity Analysis Set |
| 14.2.1.2.3 | Seroprotection Rate – Tetanus - Participants Not Longer-Term Seroprotected at Baseline – Immunogenicity Analysis Set |
| 14.2.2.1.1 | Geometric Mean Antibodies - Diphtheria – Immunogenicity Analysis Set |
| 14.2.2.1.2 | Geometric Mean Antibodies - Diphtheria – By Seroprotection Status at Baseline – Immunogenicity Analysis Set |
| 14.2.2.1.3 | Geometric Mean Antibodies - Diphtheria – By Longer-Term Seroprotection Status at Baseline – Immunogenicity Analysis Set |
| 14.2.2.1.4 | Geometric Mean Antibodies - Diphtheria – By Age Group – Immunogenicity Analysis Set |
| 14.2.2.1.5 | Geometric Mean Antibodies - Diphtheria – By Sex – Immunogenicity Analysis Set |
| 14.2.2.1.6 | Geometric Mean Antibodies - Diphtheria – By BMI Group – Immunogenicity Analysis Set |
| 14.2.2.2.1 | Geometric Mean Antibodies - Tetanus – Immunogenicity Analysis Set |
| 14.2.2.2.2 | Geometric Mean Antibodies - Tetanus – By Seroprotection Status at Baseline – Immunogenicity Analysis Set |

| Table Number | Table Title |
|---|---|
| 14.2.2.2.3 | Geometric Mean Antibodies - Tetanus – By Longer-Term Seroprotection Status at Baseline – Immunogenicity Analysis Set |
| 14.2.2.2.4 | Geometric Mean Antibodies - Tetanus – By Age Group – Immunogenicity Analysis Set |
| 14.2.2.2.5 | Geometric Mean Antibodies - Tetanus – By Sex– Immunogenicity Analysis Set |
| 14.2.2.2.6 | Geometric Mean Antibodies - Tetanus – By BMI Group – Immunogenicity Analysis Set |
| 14.2.3.1.1 | Longer-Term Seroprotection Rate – Diphtheria – Immunogenicity Analysis Set |
| 14.2.3.1.2 | Longer-Term Seroprotection Rate – Diphtheria - Participants Not Seroprotected at Baseline – Immunogenicity Analysis Set |
| 14.2.3.1.3 | Longer-Term Seroprotection Rate – Diphtheria - Participants Not Longer-Term Seroprotected at Baseline – Immunogenicity Analysis Set |
| 14.2.3.2.1 | Longer-Term Seroprotection Rate – Tetanus – Immunogenicity Analysis Set |
| 14.2.3.2.2 | Longer-Term Seroprotection Rate – Tetanus - Participants Not Seroprotected at Baseline – Immunogenicity Analysis Set |
| 14.2.3.2.3 | Longer-Term Seroprotection Rate – Tetanus - Participants Not Longer-Term Seroprotected at Baseline – Immunogenicity Analysis Set |
| 14.3.1.1 | Reactogenicity Events Summary – Safety Analysis Set |
| 14.3.1.2 | Reactogenicity Events Summary by Severity – Safety Analysis Set |
| 14.3.2.1 | Overall Summary of Treatment-Emergent Adverse Events – Safety Analysis Set |
| 14.3.2.2 | Treatment-Emergent Adverse Events by System Organ Class and Preferred Term – Safety Analysis Set |
| 14.3.2.3 | Treatment-Emergent Adverse Events by System Organ Class, Preferred Term and Severity – Safety Analysis Set |
| 14.3.2.4 | Treatment-Emergent Adverse Events by System Organ Class, Preferred Term and Relationship to Study Intervention – Safety Analysis Set |
| 14.3.2.5 | Serious Treatment-Emergent Adverse Events by System Organ Class and Preferred Term – Safety Analysis Set |

| **Table Number** | **Table Title** |
|---|---|
| 14.3.2.6 | Treatment-Emergent Adverse Events Leading to Death or Discontinuation by System Organ Class and Preferred Term – Safety Analysis Set |
| 14.3.3.1 | Listing of Participants with Serious Adverse Events – Safety Analysis Set |
| 14.3.3.2 | Listing of Participants with Adverse Events Directly Resulting in Withdrawal – Safety Analysis Set |
| 14.3.4.1 | Haematology – Change from Baseline Summary – Safety Analysis Set |
| 14.3.4.2 | Chemistry – Change from Baseline Summary – Safety Analysis Set |
| 14.3.4.3 | Coagulation – Change from Baseline Summary – Safety Analysis Set |
| 14.3.4.4 | Thyroid Function – Change from Baseline Summary – Safety Analysis Set |

#### 13.3. List of Figures

| Figure Number | Figure Title |
|---|---|
| 14.2.1.1.1 | Box-Scatter Plot of Diphtheria Antibodies Titre – Immunogenicity Analysis Set |
| 14.2.1.1.2 | Bar Chart of Diphtheria Antibodies Titre – Immunogenicity Analysis Set |
| 14.2.1.2.1 | Box-Scatter Plot of Diphtheria Antibodies Titre - By Seroprotection Status at Baseline – Immunogenicity Analysis Set |
| 14.2.1.2.2 | Bar Chart of Diphtheria Antibodies Titre - By Seroprotection Status at Baseline – Immunogenicity Analysis Set |
| 14.2.1.3.1 | Box-Scatter Plot of Diphtheria Antibodies Titre - By Longer-Term Seroprotection Status at Baseline – Immunogenicity Analysis Set |
| 14.2.1.3.2 | Bar Chart of Diphtheria Antibodies Titre - By Longer-Term Seroprotection Status at Baseline – Immunogenicity Analysis Set |
| 14.2.1.4 | Box-Scatter Plot of Diphtheria Antibodies Titre - By Age Group – Immunogenicity Analysis Set |
| 14.2.1.5 | Box-Scatter Plot of Diphtheria Antibodies Titre - By Sex – Immunogenicity Analysis Set |
| 14.2.1.6 | Box-Scatter Plot of Diphtheria Antibodies Titre - By BMI Group – Immunogenicity Analysis Set |
| 14.2.2.1.1 | Box-Scatter Plot of Diphtheria Antibodies Titre – Immunogenicity Analysis Set |
| 14.2.2.1.2 | Bar Chart of Tetanus Antibodies Titre – Immunogenicity Analysis Set |
| 14.2.2.2.1 | Box-Scatter Plot of Tetanus Antibodies Titre - By Seroprotection Status at Baseline – Immunogenicity Analysis Set |
| 14.2.2.2.2 | Bar Chart of Tetanus Antibodies Titre - By Seroprotection Status at Baseline – Immunogenicity Analysis Set |
| 14.2.2.3.1 | Box-Scatter Plot of Tetanus Antibodies Titre - By Longer-Term Seroprotection Status at Baseline – Immunogenicity Analysis Set |
| 14.2.2.3.2 | Bar Chart of Tetanus Antibodies Titre - By Longer-Term Seroprotection Status at Baseline – Immunogenicity Analysis Set |
| 14.2.2.4 | Box-Scatter Plot of Tetanus Antibodies Titre - By Age Group – Immunogenicity Analysis Set |
| 14.2.2.5 | Box-Scatter Plot of Tetanus Antibodies Titre - By Sex – Immunogenicity Analysis Set |
| 14.2.2.6 | Box-Scatter Plot of Tetanus Antibodies Titre - By BMI Group – Immunogenicity Analysis Set |

#### 13.4. List of Listings

| Listing Number | Listing Title |
|---|---|
| 16.2.1.1 | Participant Disposition |
| 16.2.1.2 | Failed Inclusion and Exclusion Criteria |
| 16.2.2 | Protocol Deviations |
| 16.2.3 | Participant Analysis Sets |
| 16.2.4.1 | Demographic Characteristics and Body Measurements |
| 16.2.4.2 | Child-Bearing Potential and Contraception Use |
| 16.2.4.3 | Medical History |
| 16.2.4.4 | Prior and Concomitant Medications |
| 16.2.4.5 | Substance Use History |
| 16.2.5.1 | Study Drug Administration |
| 16.2.5.2 | Post-Administration Observation |
| 16.2.6.1 | Antibody Titres |
| 16.2.6.2 | Seroprotection |
| 16.2.7.1 | Reactogenicity Events |
| 16.2.7.2 | Adverse Events |
| 16.2.8.1 | Haematology |
| 16.2.8.2 | Chemistry |
| 16.2.8.3 | Coagulation |
| 16.2.8.4 | Thyroid Function |
| 16.2.8.5 | Urinalysis |
| 16.2.9.1 | Pregnancy Tests |
| 16.2.9.2 | Drugs of Abuse Screen |
| 16.2.9.3 | Viral Serology |
| 16.2.10 | Vital Signs |
| 16.2.11 | Electrocardiogram |

| Listing Number | Listing Title |
|---|---|
| 16.2.12 | Physical Examination |
| 16.2.13 | Visit Dates |

#### 14. TABLE AND LISTING SHELLS

E-Sign ID: dc6e1da8-9522-4e0f-8283-c8e65365bf28

Table 14.1.1: Participant Disposition
All Participants

| | SPVX02 (N = XX) | | Tetadif (N = XX) | | diTeBooster (N = XX) | |
|---|---|---|---|---|---|---|
| | n | (%) | n | (%) | n | (%) |
| Enrolled | XX | | XX | | XX | |
| Randomised | XX | | XX | | XX | |
| Safety Analysis Set [1] | XX | (XX.X) | XX | (XX.X) | XX | (XX.X) |
| Full Analysis Set [2] | XX | (XX.X) | XX | (XX.X) | XX | (XX.X) |
| Immunogenicity Analysis Set [3] | XX | (XX.X) | XX | (XX.X) | XX | (XX.X) |
| Completed Study [4] | XX | (XX.X) | XX | (XX.X) | XX | (XX.X) |
| Withdrew from Study | XX | (XX.X) | XX | (XX.X) | XX | (XX.X) |
| Reason for Withdrawal: | | | | | | |
| Reason 1 | XX | (XX.X) | XX | (XX.X) | XX | (XX.X) |
| … | XX | (XX.X) | XX | (XX.X) | XX | (XX.X) |

[1] Safety Analysis Set = All randomised participants who were exposed to an investigational intervention.
[2] Full Analysis Set = All randomised participants.
[3] Immunogenicity Analysis Set = All randomised participants who were exposed to an investigational intervention and have at least one baseline and one corresponding post-baseline immunogenicity measurement and have no protocol deviations that may impact their immunogenicity results.
[4] Completed Study = Received study intervention and completed all study visits up to, and including, the follow-up visit.
Note: Percentages are based on the number of randomised participants.

Source: Listing 16.2.1.1 and 16.2.3.

Table 14.1.2: Demography
Safety Analysis Set

| | Statistic | SPVX02 (N = XX) | Tetadif (N = XX) | diTeBooster (N = XX) |
|---|---|---|---|---|
| Sex | | | | |
| Male | n (%) | XX (XX.X) | XX (XX.X) | XX (XX.X) |
| Female | n (%) | XX (XX.X) | XX (XX.X) | XX (XX.X) |
| Age at Informed Consent (years) | n | XX | XX | XX |
| | Mean (SD) | XX.X (XX.XX) | XX.X (XX.XX) | XX.X (XX.XX) |
| | Median | XX.X (XX.XX) | XX.X (XX.XX) | XX.X (XX.XX) |
| | Q1, Q3 | XX.X | XX.X | XX.X |
| | Min, Max | XX.X | XX.X | XX.X |
| Ethnicity | | | | |
| Hispanic or Latino | n (%) | XX (XX.X) | XX (XX.X) | XX (XX.X) |
| Not Hispanic or Latino | n (%) | XX (XX.X) | XX (XX.X) | XX (XX.X) |
| Not Reported | n (%) | XX (XX.X) | XX (XX.X) | XX (XX.X) |
| Unknown | n (%) | XX (XX.X) | XX (XX.X) | XX (XX.X) |
| Race | | | | |
| Asian | n (%) | XX (XX.X) | XX (XX.X) | XX (XX.X) |
| American Indian or Alaska Native | n (%) | XX (XX.X) | XX (XX.X) | XX (XX.X) |
| Black or African American | n (%) | XX (XX.X) | XX (XX.X) | XX (XX.X) |
| Native Hawaiian or Other Pacific Islander | n (%) | XX (XX.X) | XX (XX.X) | XX (XX.X) |
| White | n (%) | XX (XX.X) | XX (XX.X) | XX (XX.X) |
| Other | n (%) | XX (XX.X) | XX (XX.X) | XX (XX.X) |
| Unknown | n (%) | XX (XX.X) | XX (XX.X) | XX (XX.X) |
| Child-Bearing Potential | | | | |
| Yes | n (%) | XX (XX.X) | XX (XX.X) | XX (XX.X) |
| No | n (%) | XX (XX.X) | XX (XX.X) | XX (XX.X) |
| Height at Screening (cm) | n | XX | XX | XX |
| | Mean (SD) | XX.X (XX.XX) | XX.X (XX.XX) | XX.X (XX.XX) |
| | Median | XX.X (XX.XX) | XX.X (XX.XX) | XX.X (XX.XX) |
| | Q1, Q3 | XX.X | XX.X | XX.X |
| | Min, Max | XX.X | XX.X | XX.X |
| *[Repeat for, Weight at Screening (kg) and BMI at Screening (kg/m^2)]* | | | | |

Note: Percentages are based on the number of non-missing observations.

Source: Listing 16.2.4.1.

E-Sign ID: dc6e1da8-9522-4e0f-8283-c8e65365bf28

Table 14.1.3: Medical History by System Organ Class and Preferred Term
Safety Analysis Set

| System Organ Class<br>Preferred Term [1] | SPVX02<br>(N = XX) | | Tetadif<br>(N = XX) | | diTeBooster<br>(N = XX) | |
|---|---|---|---|---|---|---|
| | n | (%) | n | (%) | n | (%) |
| Number of Events | XX | | XX | | XX | |
| Number of Participants with any History | XX | (XX.X) | XX | (XX.X) | XX | (XX.X) |
| System Organ Class 1 | XX | (XX.X) | XX | (XX.X) | XX | (XX.X) |
| Preferred Term 1 | XX | (XX.X) | XX | (XX.X) | XX | (XX.X) |
| Preferred Term 2 | XX | (XX.X) | XX | (XX.X) | XX | (XX.X) |
| System Organ Class 2 | XX | (XX.X) | XX | (XX.X) | XX | (XX.X) |
| Preferred Term 1 | XX | (XX.X) | XX | (XX.X) | XX | (XX.X) |
| Preferred Term 2 | XX | (XX.X) | XX | (XX.X) | XX | (XX.X) |

[1] MedDRA Version 28.0
Note: Percentages are based on the safety analysis set.

Source: Listing 16.2.4.3.

Table 14.1.4: Concomitant Medications
Safety Analysis Set

| Drug Class (L2)/ WHO Drug Preferred Base Name [1] | SPVX02 (N = XX) | | Tetadif (N = XX) | | diTeBooster (N = XX) | | Overall (N = XX) | |
|---|---|---|---|---|---|---|---|---|
| | n | (%) | n | (%) | n | (%) | n | (%) |
| Number of Participants with any Medication | XX | (XX.X) | XX | (XX.X) | XX | (XX.X) | XX | (XX.X) |
| Drug Class 1 | XX | (XX.X) | XX | (XX.X) | XX | (XX.X) | XX | (XX.X) |
| WHO Drug Name 1 | XX | (XX.X) | XX | (XX.X) | XX | (XX.X) | XX | (XX.X) |
| WHO Drug Name 2 | XX | (XX.X) | XX | (XX.X) | XX | (XX.X) | XX | (XX.X) |
| Drug Class 2 | XX | (XX.X) | XX | (XX.X) | XX | (XX.X) | XX | (XX.X) |
| WHO Drug Name 1 | XX | (XX.X) | XX | (XX.X) | XX | (XX.X) | XX | (XX.X) |
| WHO Drug Name 2 | XX | (XX.X) | XX | (XX.X) | XX | (XX.X) | XX | (XX.X) |

[1] WHO Drug Dictionary Global version 2025 (March 2025). L2: The second level of Anatomical Therapeutic Chemical (ATC) classification.
Note: Concomitant medications are defined as those that were ongoing at the start of study intervention or started after the initiation of study intervention.
Note: Percentages are based on the safety analysis set.

Source: Listing 16.2.4.4.

E-Sign ID: dc6e1da8-9522-4e0f-8283-c8e65365bf28

Table 14.2.1.1.1: Seroprotection Rate - Diphtheria
Immunogenicity Analysis Set

| | Statistic | SPVX02 (N = XX) | Tetadif (N = XX) | diTeBooster (N = XX) |
|---|---|---|---|---|
| Seroprotection | n (%) | XX (XX.X) | XX (XX.X) | XX (XX.X) |
| | 95% CI | XX.X, XX.X | XX.X, XX.X | XX.X, XX.X |
| 2-Fold Seroprotection | n (%) | XX (XX.X) | XX (XX.X) | XX (XX.X) |
| | 95% CI | XX.X, XX.X | XX.X, XX.X | XX.X, XX.X |
| 4-Fold Seroprotection | n (%) | XX (XX.X) | XX (XX.X) | XX (XX.X) |
| | 95% CI | XX.X, XX.X | XX.X, XX.X | XX.X, XX.X |

Note: Seroprotection is defined as IgG serum antibody titre of >= 0.1 IU/mL.
Note: 2-Fold seroprotection is defined as IgG serum antibody titre of >= 0.1 IU/mL and at least a 2-fold increase from baseline.
Note: 4-Fold seroprotection is defined as IgG serum antibody titre of >= 0.1 IU/mL and at least a 2-fold increase from baseline.
Note: Percentages are based on the immunogenicity analysis set.

Source: Listing 16.2.6.2.

This shell will be used for the following tables

Table 14.2.1.1.2: Seroprotection Rate - Diphtheria - Participants Not Seroprotected at Baseline
Table 14.2.1.1.3: Seroprotection Rate - Diphtheria - Participants Not Longer-Term Seroprotected at Baseline
Table 14.2.1.2.1: Seroprotection Rate - Tetanus
Table 14.2.1.2.2: Seroprotection Rate - Tetanus - Participants Not Seroprotected at Baseline
Table 14.2.1.2.3: Seroprotection Rate - Tetanus - Participants Not Longer-Term Seroprotected at Baseline

Table 14.2.2.1.1: Geometric Mean Antibodies - Diphtheria
Immunogenicity Analysis Set

| | Statistic | SPVX02 (N = XX) | Tetadif (N = XX) | diTeBooster (N = XX) |
|---|---|---|---|---|
| Diphtheria Antibody Titre (IU/mL) | | | | |
| Baseline | n | X | X | X |
| | Mean (SD) | XX.X (XX.XX) | XX.X (XX.XX) | XX.X (XX.XX) |
| | GM (GSD) | XX.X (XX.XX) | XX.X (XX.XX) | XX.X (XX.XX) |
| | Median | XX.X | XX.X | XX.X |
| | Q1, Q3 | XX.X, XX.X | XX.X, XX.X | XX.X, XX.X |
| | Min, Max | XX, XX | XX, XX | XX, XX |
| Day 28 | n | X | X | X |
| | Mean (SD) | XX.X (XX.XX) | XX.X (XX.XX) | XX.X (XX.XX) |
| | GM (GSD) | XX.X (XX.XX) | XX.X (XX.XX) | XX.X (XX.XX) |
| | Median | XX.X | XX.X | XX.X |
| | Q1, Q3 | XX.X, XX.X | XX.X, XX.X | XX.X, XX.X |
| | Min, Max | XX, XX | XX, XX | XX, XX |
| Geometric Mean Ratio | GMR (GSD) | XX.X (XX.XX) | XX.X (XX.XX) | XX.X (XX.XX) |
| | 95% CI | XX.X, XX.X | XX.X, XX.X | XX.X, XX.X |

Source: Listing 16.2.6.1.

This shell will be used for

Table 14.2.2.1.2: Geometric Mean Antibodies - Diphtheria - By Seroprotection Status at Baseline
Table 14.2.2.1.3: Geometric Mean Antibodies - Diphtheria - By Longer-Term Seroprotection Status at Baseline
Table 14.2.2.1.4: Geometric Mean Antibodies - Diphtheria - By Age Group
Table 14.2.2.1.5: Geometric Mean Antibodies - Diphtheria - By Sex
Table 14.2.2.1.6: Geometric Mean Antibodies - Diphtheria - By BMI Group
Table 14.2.2.2.1: Geometric Mean Antibodies - Tetanus
Table 14.2.2.2.2: Geometric Mean Antibodies - Tetanus - By Seroprotection Status at Baseline
Table 14.2.2.2.3: Geometric Mean Antibodies - Tetanus - By Longer-Term Seroprotection Status at Baseline
Table 14.2.2.2.4: Geometric Mean Antibodies - Tetanus - By Age Group
Table 14.2.2.2.5: Geometric Mean Antibodies - Tetanus - By Sex
Table 14.2.2.2.6: Geometric Mean Antibodies - Tetanus - By BMI Group

Table 14.2.3.1.1: Longer-Term Seroprotection Rate - Diphtheria
Immunogenicity Analysis Set

| | Statistic | SPVX02 (N = XX) | Tetadif (N = XX) | diTeBooster (N = XX) |
|---|---|---|---|---|
| Seroprotection | n (%) | XX (XX.X) | XX (XX.X) | XX (XX.X) |
| | 95% CI | XX.X, XX.X | XX.X, XX.X | XX.X, XX.X |
| 2-Fold Seroprotection | n (%) | XX (XX.X) | XX (XX.X) | XX (XX.X) |
| | 95% CI | XX.X, XX.X | XX.X, XX.X | XX.X, XX.X |
| 4-Fold Seroprotection | n (%) | XX (XX.X) | XX (XX.X) | XX (XX.X) |
| | 95% CI | XX.X, XX.X | XX.X, XX.X | XX.X, XX.X |

Note: Longer-term seroprotection is defined as IgG serum antibody titre of >= 1.0 IU/mL.
Note: 2-Fold longer-term seroprotection is defined as IgG serum antibody titre of >= 1.0 IU/mL and at least a 2-fold increase from baseline.
Note: 4-Fold longer-term seroprotection is defined as IgG serum antibody titre of >= 1.0 IU/mL and at least a 2-fold increase from baseline.
Note: Percentages are based on the immunogenicity analysis set.

Source: Listing 16.2.6.2.

This shell will be used for the following tables

Table 14.2.3.1.2: Longer-Term Seroprotection Rate - Diphtheria - Participants Not Seroprotected at Baseline
Table 14.2.3.1.3: Longer-Term Seroprotection Rate - Diphtheria - Participants Not Longer-Term Seroprotected at Baseline
Table 14.2.3.2.1: Longer-Term Seroprotection Rate - Tetanus
Table 14.2.3.2.2: Longer-Term Seroprotection Rate - Tetanus - Participants Not Seroprotected at Baseline
Table 14.2.3.2.3: Longer-Term Seroprotection Rate - Tetanus - Participants Not Longer-Term Seroprotected at Baseline

E-Sign ID: dc6e1da8-9522-4e0f-8283-c8e65365bf28

Table 14.3.1.1: Reactogenicity Events Summary
Safety Analysis Set

| Symptom<br>Day | SPVX02<br>(N = XX) | | Tetadif<br>(N = XX) | | diTeBooster<br>(N = XX) | | Overall<br>(N = XX) | |
|---|---|---|---|---|---|---|---|---|
| | n | (%) | n | (%) | n | (%) | n | (%) |
| Symptom 1 | | | | | | | | |
| Any | XX | (XX.X) | XX | (XX.X) | XX | (XX.X) | XX | (XX.X) |
| Day 1 | XX | (XX.X) | XX | (XX.X) | XX | (XX.X) | XX | (XX.X) |
| Day 2 | XX | (XX.X) | XX | (XX.X) | XX | (XX.X) | XX | (XX.X) |
| Day 3 | XX | (XX.X) | XX | (XX.X) | XX | (XX.X) | XX | (XX.X) |
| Day 4 | XX | (XX.X) | XX | (XX.X) | XX | (XX.X) | XX | (XX.X) |
| Day 5 | XX | (XX.X) | XX | (XX.X) | XX | (XX.X) | XX | (XX.X) |
| Day 6 | XX | (XX.X) | XX | (XX.X) | XX | (XX.X) | XX | (XX.X) |
| Day 7 | XX | (XX.X) | XX | (XX.X) | XX | (XX.X) | XX | (XX.X) |
| *[Repeat for each symptom]* | | | | | | | | |
| Any Symptom | | | | | | | | |
| Any | XX | (XX.X) | XX | (XX.X) | XX | (XX.X) | XX | (XX.X) |
| Day 1 | XX | (XX.X) | XX | (XX.X) | XX | (XX.X) | XX | (XX.X) |
| Day 2 | XX | (XX.X) | XX | (XX.X) | XX | (XX.X) | XX | (XX.X) |
| Day 3 | XX | (XX.X) | XX | (XX.X) | XX | (XX.X) | XX | (XX.X) |
| Day 4 | XX | (XX.X) | XX | (XX.X) | XX | (XX.X) | XX | (XX.X) |
| Day 5 | XX | (XX.X) | XX | (XX.X) | XX | (XX.X) | XX | (XX.X) |
| Day 6 | XX | (XX.X) | XX | (XX.X) | XX | (XX.X) | XX | (XX.X) |
| Day 7 | XX | (XX.X) | XX | (XX.X) | XX | (XX.X) | XX | (XX.X) |

Note: Symptoms recorded by the participants in their eDiaries, summarised here, were interpreted by the Investigator and subsequently recorded as AEs.
Note: Percentages are based on the safety analysis set.

Source: Listing 16.2.7.1.

Table 14.3.1.2: Reactogenicity Events by Severity Summary
Safety Analysis Set

| Symptom<br>Day | SPVX02<br>(N = XX) | | Tetadif<br>(N = XX) | | diTeBooster<br>(N = XX) | | Overall<br>(N = XX) | |
|---|---|---|---|---|---|---|---|---|
| | n | (%) | n | (%) | n | (%) | n | (%) |
| Symptom 1 | | | | | | | | |
| Any | | | | | | | | |
| Mild | XX | (XX.X) | XX | (XX.X) | XX | (XX.X) | XX | (XX.X) |
| Moderate | XX | (XX.X) | XX | (XX.X) | XX | (XX.X) | XX | (XX.X) |
| Severe | XX | (XX.X) | XX | (XX.X) | XX | (XX.X) | XX | (XX.X) |
| Day 1 | | | | | | | | |
| Mild | XX | (XX.X) | XX | (XX.X) | XX | (XX.X) | XX | (XX.X) |
| Moderate | XX | (XX.X) | XX | (XX.X) | XX | (XX.X) | XX | (XX.X) |
| Severe | XX | (XX.X) | XX | (XX.X) | XX | (XX.X) | XX | (XX.X) |
| Day 2 | | | | | | | | |
| Mild | XX | (XX.X) | XX | (XX.X) | XX | (XX.X) | XX | (XX.X) |
| Moderate | XX | (XX.X) | XX | (XX.X) | XX | (XX.X) | XX | (XX.X) |
| Severe | XX | (XX.X) | XX | (XX.X) | XX | (XX.X) | XX | (XX.X) |
| *[Repeat for each day and for each symptom]* | | | | | | | | |
| Any Symptom | | | | | | | | |
| Any | | | | | | | | |
| Mild | XX | (XX.X) | XX | (XX.X) | XX | (XX.X) | XX | (XX.X) |
| Moderate | XX | (XX.X) | XX | (XX.X) | XX | (XX.X) | XX | (XX.X) |
| Severe | XX | (XX.X) | XX | (XX.X) | XX | (XX.X) | XX | (XX.X) |
| Day 1 | | | | | | | | |
| Mild | XX | (XX.X) | XX | (XX.X) | XX | (XX.X) | XX | (XX.X) |
| Moderate | XX | (XX.X) | XX | (XX.X) | XX | (XX.X) | XX | (XX.X) |
| Severe | XX | (XX.X) | XX | (XX.X) | XX | (XX.X) | XX | (XX.X) |
| Day 2 | | | | | | | | |
| Mild | XX | (XX.X) | XX | (XX.X) | XX | (XX.X) | XX | (XX.X) |
| Moderate | XX | (XX.X) | XX | (XX.X) | XX | (XX.X) | XX | (XX.X) |
| Severe | XX | (XX.X) | XX | (XX.X) | XX | (XX.X) | XX | (XX.X) |
| *[Repeat for each day and for each symptom]* | | | | | | | | |

Note: Symptoms recorded by the participants in their eDiaries, summarised here, were interpreted by the Investigator and subsequently recorded as AEs.
Note: For the Any Symptom summaries, if the participant experienced more than one symptom the worst severity will be summarised.
Note: Percentages are based on the safety analysis set.

Source: Listing 16.2.7.1.

Table 14.3.2.1: Overall Summary of Treatment-Emergent Adverse Events
Safety Analysis Set

| | SPVX02 (N = XX) | | Tetadif (N = XX) | | diTeBooster (N = XX) | | Overall (N = XX) | |
|---|---|---|---|---|---|---|---|---|
| Total Number of Participants | n | (%) | n | (%) | n | (%) | n | (%) |
| TEAEs | XX | (XX.X) | XX | (XX.X) | XX | (XX.X) | XX | (XX.X) |
| Related | | | | | | | | |
| Related | XX | (XX.X) | XX | (XX.X) | XX | (XX.X) | XX | (XX.X) |
| Unrelated | XX | (XX.X) | XX | (XX.X) | XX | (XX.X) | XX | (XX.X) |
| Severity | XX | (XX.X) | XX | (XX.X) | XX | (XX.X) | XX | (XX.X) |
| Mild | XX | (XX.X) | XX | (XX.X) | XX | (XX.X) | XX | (XX.X) |
| Moderate | XX | (XX.X) | XX | (XX.X) | XX | (XX.X) | XX | (XX.X) |
| Severe | XX | (XX.X) | XX | (XX.X) | XX | (XX.X) | XX | (XX.X) |
| Serious TEAE | XX | (XX.X) | XX | (XX.X) | XX | (XX.X) | XX | (XX.X) |
| TEAE Leading to Discontinuation | XX | (XX.X) | XX | (XX.X) | XX | (XX.X) | XX | (XX.X) |

Note: Treatment-Emergent Adverse Event (TEAE) is an AE occurring on or after the first dose of study intervention.
Note: If a participant experiences more than one event, then the event with the worst severity or strongest relationship is included in the relevant level of summarisation.
Note: Related AEs are those where the relationship has been recorded as possible, probable, unlikely, unknown or missing. Unrelated AEs are those recorded as not related.
Note: Adverse Event Leading to Discontinuation = Participant withdrew from the study due to an adverse event or serious adverse event.
Note: Percentages are based on the safety analysis set.

Source: Listing 16.2.7.2.

Table 14.3.2.2: Treatment-Emergent Adverse Events by System Organ Class and Preferred Term
Safety Analysis Set

| System Organ Class<br>Preferred Term [1] | SPVX02<br>(N = XX) | | | Tetadif<br>(N = XX) | | | diTeBooster<br>(N = XX) | | | Overall<br>(N = XX) | | |
|---|---|---|---|---|---|---|---|---|---|---|---|---|
| | E | n | (%) | E | n | (%) | E | n | (%) | E | n | (%) |
| Number of Participants with any Adverse Event | XX | XX | (XX.X) | XX | XX | (XX.X) | XX | XX | (XX.X) | XX | XX | (XX.X) |
| System Organ Class 1 | XX | XX | (XX.X) | XX | XX | (XX.X) | XX | XX | (XX.X) | XX | XX | (XX.X) |
| Preferred Term 1 | XX | XX | (XX.X) | XX | XX | (XX.X) | XX | XX | (XX.X) | XX | XX | (XX.X) |
| Preferred Term 2 | XX | XX | (XX.X) | XX | XX | (XX.X) | XX | XX | (XX.X) | XX | XX | (XX.X) |
| System Organ Class 2 | XX | XX | (XX.X) | XX | XX | (XX.X) | XX | XX | (XX.X) | XX | XX | (XX.X) |
| Preferred Term 1 | XX | XX | (XX.X) | XX | XX | (XX.X) | XX | XX | (XX.X) | XX | XX | (XX.X) |
| Preferred Term 2 | XX | XX | (XX.X) | XX | XX | (XX.X) | XX | XX | (XX.X) | XX | XX | (XX.X) |

[1] MedDRA Version 28.0
Note: Treatment-Emergent Adverse Event is an AE occurring on or after the first dose of study intervention.
Note: Table shows distinct number of participants with events (n) and number of events (E) for each system organ class/preferred term.
Note: Percentages are based on the safety analysis set.

Source: Listing 16.2.7.2.

Table 14.3.2.3: Treatment-Emergent Adverse Events by System Organ Class, Preferred Term and Severity
Safety Analysis Set

| System Organ Class<br>Preferred Term [1] | SPVX02<br>(N = XX) | | | Tetadif<br>(N = XX) | | | diTeBooster<br>(N = XX) | | | Overall<br>(N = XX) | | |
|---|---|---|---|---|---|---|---|---|---|---|---|---|
| | E | n | (%) | E | n | (%) | E | n | (%) | E | n | (%) |
| Number of Participants with any Adverse Event | XX | XX | (XX.X) | XX | XX | (XX.X) | XX | XX | (XX.X) | XX | XX | (XX.X) |
| System Organ Class 1 | | | | | | | | | | | | |
| Mild | XX | XX | (XX.X) | XX | XX | (XX.X) | XX | XX | (XX.X) | XX | XX | (XX.X) |
| Moderate | XX | XX | (XX.X) | XX | XX | (XX.X) | XX | XX | (XX.X) | XX | XX | (XX.X) |
| Severe | XX | XX | (XX.X) | XX | XX | (XX.X) | XX | XX | (XX.X) | XX | XX | (XX.X) |
| Preferred Term 1 | | | | | | | | | | | | |
| Mild | XX | XX | (XX.X) | XX | XX | (XX.X) | XX | XX | (XX.X) | XX | XX | (XX.X) |
| Moderate | XX | XX | (XX.X) | XX | XX | (XX.X) | XX | XX | (XX.X) | XX | XX | (XX.X) |
| Severe | XX | XX | (XX.X) | XX | XX | (XX.X) | XX | XX | (XX.X) | XX | XX | (XX.X) |
| Preferred Term 2 | | | | | | | | | | | | |
| Mild | XX | XX | (XX.X) | XX | XX | (XX.X) | XX | XX | (XX.X) | XX | XX | (XX.X) |
| Moderate | XX | XX | (XX.X) | XX | XX | (XX.X) | XX | XX | (XX.X) | XX | XX | (XX.X) |
| Severe | XX | XX | (XX.X) | XX | XX | (XX.X) | XX | XX | (XX.X) | XX | XX | (XX.X) |

[1] MedDRA Version 28.0
Note: Treatment-Emergent Adverse Event is an AE occurring on or after the first dose of study intervention.
Note: Table shows distinct number of participants with events (n) and number of events (E) for each system organ class/preferred term/severity.
Note: If a participant experienced a specific event more than once then the event with the worst severity is summarised.
Note: Percentages are based on the safety analysis set.

Source: Listing 16.2.7.2.

This shell will be used for the following tables:

Table 14.3.2.5: Serious Treatment-Emergent Adverse Events by System Organ Class and Preferred Term
Table 14.3.2.6: Treatment-Emergent Adverse Events Leading to Death or Discontinuation by System Organ Class and Preferred Term

Table 14.3.2.4: Treatment-Emergent Adverse Events by System Organ Class, Preferred Term and Relationship to Study Intervention
Safety Analysis Set

| System Organ Class<br>Preferred Term [1] | SPVX02<br>(N = XX) | | | Tetadif<br>(N = XX) | | | diTeBooster<br>(N = XX) | | | Overall<br>(N = XX) | | |
|---|---|---|---|---|---|---|---|---|---|---|---|---|
| | E | n | (%) | E | n | (%) | E | n | (%) | E | n | (%) |
| Number of Participants with any Adverse Event | XX | XX | (XX.X) | XX | XX | (XX.X) | XX | XX | (XX.X) | XX | XX | (XX.X) |
| System Organ Class 1 | | | | | | | | | | | | |
| Related | XX | XX | (XX.X) | XX | XX | (XX.X) | XX | XX | (XX.X) | XX | XX | (XX.X) |
| Unrelated | XX | XX | (XX.X) | XX | XX | (XX.X) | XX | XX | (XX.X) | XX | XX | (XX.X) |
| Preferred Term 1 | | | | | | | | | | | | |
| Related | XX | XX | (XX.X) | XX | XX | (XX.X) | XX | XX | (XX.X) | XX | XX | (XX.X) |
| Unrelated | XX | XX | (XX.X) | XX | XX | (XX.X) | XX | XX | (XX.X) | XX | XX | (XX.X) |
| Preferred Term 2 | | | | | | | | | | | | |
| Related | XX | XX | (XX.X) | XX | XX | (XX.X) | XX | XX | (XX.X) | XX | XX | (XX.X) |
| Unrelated | XX | XX | (XX.X) | XX | XX | (XX.X) | XX | XX | (XX.X) | XX | XX | (XX.X) |

[1] MedDRA Version 28.0
Note: Treatment-Emergent Adverse Event is an AE occurring on or after the first dose of study intervention.
Note: Table shows distinct number of participants with events (n) and number of events (E) for each system organ class/preferred term.
Note: Related AEs are those where the relationship has been recorded as possible, probable, unlikely, unknown or missing. Unrelated AEs are those recorded as not related.
Note: Percentages are based on the safety analysis set.

Source: Listing 16.2.7.2.

E-Sign ID: dc6e1da8-9522-4e0f-8283-c8e65365bf28

Table 14.3.3.1: Listing of Participants with Serious Adverse Events
Safety Analysis Set

| Intervention / Participant Number / AE Number | MedDRA SOC / Preferred Term[1] / Adverse event (verbatim) | Start Date Time (Study day) / Stop Date Time (Study day) | TE [2] | Related [3] / Severity [4] |
|---|---|---|---|---|
| XXXX<br>XXXX<br>XX | XXXXXXXXX /<br>XXXXXX /<br>XXXXXX | DDMMMYYYY HH:MM (XX) /<br>DDMMMYYYY HH:MM (XX) | Yes or No | X /<br>X |

[1] MedDRA Version 28.0.
[2] TE = Treatment-Emergent. An AE occurring on or after administration of study intervention.
[3] Related: 1 = Related; 2 = Probably Related; 3 = Possibly Related; 4 = Unlikely Related; 5 = Not Related.
[4] Severity: 1 = Mild; 2 = Moderate; 3 = Severe.

Source: Listing 16.2.7.2.

E-Sign ID: dc6e1da8-9522-4e0f-8283-c8e65365bf28

Table 14.3.3.2: Listing of Participants with Adverse Events Directly Resulting in Withdrawal
Safety Analysis Set

| Treatment Sequence / Screening Number / AE Number | MedDRA SOC / Preferred Term [1] / Adverse event (verbatim) | Start Date Time (Study day) / Stop Date Time (Study day) | TE [2] | Related [3] / Severity [4] |
|---|---|---|---|---|
| XXXX<br>XXXX<br>XX | XXXXXXXXX /<br>XXXXXX /<br>XXXXXX | DDMMMYYYY HH:MM (XX) /<br>DDMMMYYYY HH:MM (XX) | Yes or No | X /<br>X |

[1] MedDRA Version 28.0
[2] TE = Treatment-Emergent. An AE occurring on or after administration of study intervention.
[3] Related: 1 = Related; 2 = Probably Related; 3 = Possibly Related; 4 = Unlikely Related; 5 = Not Related.
[4] Severity: 1 = Mild; 2 = Moderate; 3 = Severe;

Source: Listing 16.2.7.2.

E-Sign ID: dc6e1da8-9522-4e0f-8283-c8e65365bf28

Table 14.3.4.1: Haematology – Change from Baseline Summary
Safety Analysis Set

| Parameter (Units) Visit | Statistic | SPVX02 (N = XX) Absolute | SPVX02 (N = XX) Change from Baseline | Tetadif (N = XX) Absolute | Tetadif (N = XX) Change from Baseline | diTeBooster (N = XX) Absolute | diTeBooster (N = XX) Change from Baseline |
|---|---|---|---|---|---|---|---|
| XXXX (XXX) | | | | | | | |
| Baseline | n | X | | X | | X | |
| | Mean (SD) | XX.X (XX.XX) | | XX.X (XX.XX) | | XX.X (XX.XX) | |
| | Median | XX.X | | XX.X | | XX.X | |
| | Q1, Q3 | XX.X, XX.X | | XX.X, XX.X | | XX.X, XX.X | |
| | Min, Max | XX, XX | | XX, XX | | XX, XX | |
| Follow-up | n | X | X | X | X | X | X |
| | Mean (SD) | XX.X (XX.XX) | XX.X (XX.XX) | XX.X (XX.XX) | XX.X (XX.XX) | XX.X (XX.XX) | XX.X (XX.XX) |
| | Median | XX.X | XX.X | XX.X | XX.X | XX.X | XX.X |
| | Q1, Q3 | XX.X, XX.X | XX.X, XX.X | XX.X, XX.X | XX.X, XX.X | XX.X, XX.X | XX.X, XX.X |
| | Min, Max | XX, XX | XX, XX | XX, XX | XX, XX | XX, XX | XX, XX |

Source: Listing 16.2.8.1.

This shell will be used for the following tables, with the source listing number updated as required:

Table 14.3.4.2: Chemistry – Change from Baseline Summary
Table 14.3.4.3: Coagulation – Change from Baseline Summary
Table 14.3.4.4: Thyroid Function – Change from Baseline Summary

Listing 16.2.1.1: Participant Disposition

| Intervention | Participant Number/ Randomisation Number | Informed Consent Protocol Version/IC Version: Date/Time | Date of Randomisation | Date (Study Day) of Study Completion | Date (Study Day) of Early Withdrawal | Reason not Randomised/ Reason for Early Withdrawal |
|---|---|---|---|---|---|---|
| XXX | XXXXXX/ XXXXXX | X/X: DDMMMYYYY HH:MM | DDMMMYYYY | DDMMMYYYY (XX) | DDMMMYYYY (XX) | XXXXXXXXXXXXXXX |

*Programming Note: Include every re-consent*

*Programming Note: The last column should include any reasons for not randomizing or reasons for withdrawal.*

Listing 16.2.1.2: Failed Inclusion and Exclusion Criteria

| Participant Number | Reason for Exclusion | Comments |
|---|---|---|
| XXXXXX | XXXXXXXXXXXXXXXXXX | XXXXXXXXXXX |

*Programming note: Listing will state 'No Data to Report' if no participants fail the criteria.*

E-Sign ID: dc6e1da8-9522-4e0f-8283-c8e65365bf28

Listing 16.2.2: Protocol Deviations

| Intervention | Participant Number | Date Occurred (Study Day) | Deviation Description | Deviation Category | Other, Details |
|---|---|---|---|---|---|
| XXXXX | XXXXXX | DDMMMYYYY (XX) | XXXXXXXXX | XXXXXXX | XXXXXXXX |

E-Sign ID: dc6e1da8-9522-4e0f-8283-c8e65365bf28

Listing 16.2.3: Participant Analysis Sets

| Intervention | Participant Number | Safety Analysis Set [1] | Full Analysis Set [2] | Immunogenicity Analysis Set [3] |
|---|---|---|---|---|
| XXXXX | XXXXXX | Yes / No | Yes / No | Yes / No |

[1] Safety Analysis Set = All randomised participants who were exposed to an investigational intervention.
[2] Full Analysis Set = All randomised participants.
[3] Immunogenicity Analysis Set = All randomised participants who were exposed to an investigational intervention and have at least one baseline and one corresponding post-baseline immunogenicity measurement and have no protocol deviations that may impact their immunogenicity results.

Listing 16.2.4.1: Demographic Characteristics and Body Measurements

| Intervention | Participant Number | Age at informed consent (years) | Sex | Race / Ethnicity | Height (cm) | Weight (kg) | BMI (kg/m^2) |
|---|---|---|---|---|---|---|---|
| XXXXX | XXXXXX | xx | Male / Female | XXXXXX / XXXXXX | XXX | XX.X | XX.X |

*Programming Note: Height, Weight and BMI here are those collected at screening. The weight and BMI recorded at any other visits will be included in the vital signs listing.*

E-Sign ID: dc6e1da8-9522-4e0f-8283-c8e65365bf28

Listing 16.2.4.2: Child-Bearing Potential and Contraception Use

| Intervention | Participant Number | Child-Bearing Potential | If Not, Reason | Contraceptive Precautions | Other, Details |
|---|---|---|---|---|---|
| XXXXX | XXXXXX | Yes/No | XXXXXXXXX | XXXX / XXXX/ XXXX | XXXXXX |

Note: ^ = Female.

E-Sign ID: dc6e1da8-9522-4e0f-8283-c8e65365bf28

Listing 16.2.4.3: Medical History

| Intervention | Participant Number | MedDRA SOC / Preferred Term [1] / Medical Condition | Start Date/ End Date | Medical History ID |
|---|---|---|---|---|
| XXXXX | XXXXXX | XXXXXXXXX / XXXXXXXXX / XXXXXXXXX | DDMMMYYYY / DDMMMYYYY | XXX |

[1] MedDRA Version 28.0.

Note: The modified term is the term that was used for MedDRA coding of the event where the event required multiple codes.

Listing 16.2.4.4: Prior and Concomitant Medications

| Intervention | Participant Number | Drug Class (L2) / WHO Drug Preferred Base Name [1] / Medication (verbatim) <modified> | Start Date/Time (Study Day)/ End Date/Time (Study Day) | Indication / AE ID/ Med ID | Dose / Dose Units | Freq [2] / Route [3] | Prior or Concomitant[4] |
|---|---|---|---|---|---|---|---|
| XXXXX | XXXXXX | XXXXXXXXX / XXXXXXXXX / XXXXXXXXXXXX | DDMMMYYYY HH:MM (XX) / DDMMMYYYY HH:MM (XX) | XXXXXX / AEn / MHn | XXX / XXX | XXX / XXX | P / C |

[1] WHO Drug Dictionary Global version 2025 (March 2025). L2: The second level of Anatomical Therapeutic Chemical (ATC) classification.
[2] OD = Once a day, BD = Twice a day, PRN = As needed, TDS = Three times a day, QDS = Four times a day, CTS = Continuous.
[3] A = Aural, IM = Intramuscular, INH = Inhalation, IUD = Intrauterine Device, IV = Intravenous, N = Nasal, O = Ocular, PO = Oral, PR = Rectal, PV = Vaginal, SC = Subcutaneous, SL = Sublingual, TOP = Topical.
[4] If the medication stop date is before the date of study intervention the medication will be assigned as being prior (P). In all other cases, the medication is assigned as being concomitant with study intervention (C).

*Programming Note: Please update the footnotes and codes for [2] and [3] to match what is seen in the data.*

E-Sign ID: dc6e1da8-9522-4e0f-8283-c8e65365bf28

Listing 16.2.4.5: Substance Use History

| Intervention | Participant Number | Smoking History | Alcohol History |
| --- | --- | --- | --- |
| XXXXX | XXXXXX | Never/Past/Current | Past/Current |

Listing 16.2.5.1: Study Drug Administration

| Intervention | Participant Number | Administered (as Per Protocol)? | Reason not Administered (as Per Protocol) | Administration Date/Time (Study Day)/ | Location of Administration | Monitored for Required Time? | If Not Monitored, Reason |
|---|---|---|---|---|---|---|---|
| XXXXX | XXXXXX | Yes/No | XXXXXXXXXXXXX | DDMMMYYYY HH:MM (XX) | Left Arm/ Right Arm | Yes/No | XXXXXXXXXXXXX |

Note: Required observation time is at least 1 hour for participants in the sentinel group and at least 30 mins for all other participants.

Listing 16.2.5.2: Post-Administration Observation

| Intervention | Participant Number | Symptoms | Experienced? | Severity | Measurement[1] |
|---|---|---|---|---|---|
| XXXXX | XXXXXX | XXXXXXXX | Yes/No | Mild/Moderate/Severe | XX.X |

[1] Measurement is only expected for temperature (Degrees C), induration/swelling (cm) and erythema/redness (cm).

Listing 16.2.6.1: Antibody Titres

| Intervention | Participant Number | Disease | Visit | Assessment Date/Time (Study Day) | Titre | Change from Baseline | Ratio of Baseline | Sero-Protected at Baseline | Longer-Term Sero-Protected at Baseline | Comments |
|---|---|---|---|---|---|---|---|---|---|---|
| XXXXX | XXXXXX | Tetanus/ Diphtheria | Day 1 | DDMMMYYYY HH:MM (XX) | XX.XX | | | Yes/No | Yes/No | |
| XXXXX | XXXXXX | | Day 28 | DDMMMYYYY HH:MM (XX) | XX.XX | XX.XX | XX.XX | | | |

Note: # = Baseline.

Programming Note: Comments column should include any comments from external data or reasons not done.
Programming Note: Seroprotection at baseline should appear on the baseline row for each of diphtheria and tetanus

E-Sign ID: dc6e1da8-9522-4e0f-8283-c8e65365bf28

Listing 16.2.6.2: Seroprotection

| Intervention | Participant Number | Disease | Seroprotection | Long-Term Seroprotection | 2-Fold Seroprotection | 2-Fold Long-Term Seroprotection | 4-Fold Seroprotection | 4-Fold Long-Term Seroprotection |
|---|---|---|---|---|---|---|---|---|
| XXXXX | XXXXXX | Tetanus/Diphtheria | Yes/No | Yes/No | Yes/No | Yes/No | Yes/No | Yes/No |

Listing 16.2.7.1: Reactogenicity Events

| Intervention | Participant Number | e-Diary Day | Date/Time Completed | Temp [1] | Symptoms | Experienced ? | Interference with Activities | Measurement [2] | Medication? | A&E/Hospital? | Severity Grade |
|---|---|---|---|---|---|---|---|---|---|---|---|
| XXXXX | XXXXXX | Day X | DDMMMYYYY HH:MM | | XXXXXXXX | Yes/No | None/Some/Significant | XXX | Yes/No | Yes/No | Mild/ Moderate/ Severe |

[1] Temperature is measured at the point of diary entry. Fever severity is defined as: Mild = 38.0-38.4; Moderate = 38.5-38.9; Severe = 39.0-40.0; Potentially Life Threatening = >40.0.
[2] Measurement is only expected for feverishness (Degrees C), induration/swelling (cm) and erythema/redness (cm), where it is reported that the symptom is experienced.

Listing 16.2.7.2: Adverse Events

| Intervention / Participant Number / AE Number | MedDRA SOC / Preferred Term[1] / Adverse Event (verbatim) | Start Date/Time (Study Day) / Stop Date/Time (Study Day) | TE [2] | Related [3] / Severity [4] | SAE[5] / Serious Criteria[6] | Outcome[7] / Action Taken [8]/ Other Actions, Details |
|---|---|---|---|---|---|---|
| XXXX<br>XXXX<br>XX | XXXXXXXXX /<br>XXXXXX /<br>XXXXXX | DDMMMYYYY HH:MM (XX) /<br>DDMMMYYYY HH:MM (XX) | Yes or No | X /<br>X | Yes or No /<br>XX | X /<br>X /<br>XXXXXXXXXXXXXXXXXX |

[1] MedDRA Version 28.0.
[2] TE = Treatment-Emergent. An AE occurring on or after administration of study intervention.
[3] Related: 1 = Definitely Related; 2 = Probably Related; 3 = Possibly Related; 4 = Unlikely Related; 5 = Not Related.
[4] Severity: 1 = Mild; 2 = Moderate; 3 = Severe;
[5] SAE = Serious Adverse Event.
[6] Serious Criteria: 1 = Death; 2 = Life-threatening; 3 = Hospitalization; 4 = Persistent or significant disability/incapacity; 5 = Congenital anomaly/birth defect; 6 = Other.
[7] Outcome: 1 = Recovered/Resolved; 2 = Recovered/Resolved with Sequelae; 3 = Recovering/Resolving; 4 = Not Recovered/Not Resolved; 5 = Fatal; 6 = Unknown.
[8] Action Taken: 1 = None; 2 = Non-drug therapy given; 3 = Concomitant medication taken; 4 = Participant withdrawn; 5 = Participant hospitalised; 6 = Other

*Programming Note: Update the footnotes to match what is in the final data*

Listing 16.2.8.1: Haematology

| Intervention | Participant Number | Visit | Sample Collection Date/Time (Study Day) | Parameter | Result | Change from Baseline | Units | Lower Limit | Upper Limit | Interpretation | Comments |
|---|---|---|---|---|---|---|---|---|---|---|---|
| XXXXX | XXXXXX | XXX | DDMMMYYYY HH:MM (XX) | Basophils<br>XXXX<br>XXXX<br>XXXX | X.XX | X.XX | X10^9/L | X.XX | X.XX | Normal/ Abnormal NCS / Abnormal CS | XXXXXXXXXXXX |

Note: # = Baseline; CS = Clinically Significant; NCS = Not Clinically Significant.
Note: Values prefixed with < are treated as 0 when calculating the change from baseline.

*Programming Note: The parameters should be listed in alphabetical order.*
*Programming Note: # should be placed against the baseline measurement for each parameter.*
*Programming Note: Comments column should include any comments from external data or reasons not done.*

*The same listing template will be used for*

Listing 16.2.8.2: Chemistry
Listing 16.2.8.3: Coagulation
Listing 16.2.8.4: Thyroid Function

E-Sign ID: dc6e1da8-9522-4e0f-8283-c8e65365bf28

Listing 16.2.8.5: Urinalysis

| Intervention | Participant Number | Visit | Sample Collection Date/Time (Study Day) | Parameter | Result | Comments |
|---|---|---|---|---|---|---|
| XXXXX | XXXXXX | XXX | DDMMMYYYY HH:MM (XX) | Basophils | X.XX | XXXXXXXXXXXX |
| | | | | XXXX | | |
| | | | | XXXX | | |
| | | | | XXXX | | |

*Programming Note: The parameters should be listed in alphabetical order.*

E-Sign ID: dc6e1da8-9522-4e0f-8283-c8e65365bf28

Listing 16.2.9.1: Pregnancy Tests

| Intervention | Participant Number | Visit | Test Date/Time (Study Day) | Sample Type | Result |
|---|---|---|---|---|---|
| XXXXX | XXXXXX | XXX | DDMMMYYYY HH:MM (XX) | Urine/Serum | Positive / Negative |

E-Sign ID: dc6e1da8-9522-4e0f-8283-c8e65365bf28

Listing 16.2.9.2: Drugs of Abuse Screen

| Intervention | Participant Number | Visit | Sample Collection Date/Time (Study Day) | Drug | Result | Reason Not Done |
|---|---|---|---|---|---|---|
| XXXXX | XXXXXX | XXX | DDMMMYYYY HH:MM (XX) | XXXXX | Positive / Negative / Not done | XXXXXXXX |

E-Sign ID: dc6e1da8-9522-4e0f-8283-c8e65365bf28

Listing 16.2.9.3: Viral Serology

| Intervention | Participant Number | Visit | Sample Collection Date/Time (Study Day) | Parameter | Result | Reason not Done |
|---|---|---|---|---|---|---|
| XXXXX | XXXXXX | XXX | DDMMMYYYY HH:MM (XX) | HIV | Positive / Negative | XXXXXXXX |
| | | | | HBsAg | Positive / Negative | XXXXXXXX |
| | | | | HCV | Positive / Negative | XXXXXXXX |

E-Sign ID: dc6e1da8-9522-4e0f-8283-c8e65365bf28

Listing 16.2.10: Vital Signs

| Intervention | Participant Number | Visit | Date/Time (Study Day) | Parameter | Method | Result | Units | Reason Not Done |
|---|---|---|---|---|---|---|---|---|
| XXXXX | XXXXXX | XXX | DDMMMYYYY HH:MM (XX) | XXXX | Tympani/Oral | XX | XX | XXXXXXXXX |
| | | | | XXXX | | XX | XX | |
| | | | | XXXX | | XX | XX | |
| | | | | XXXX | | XX | XX | |

*Programming Note: Method is only relevant to temperature*

Listing 16.2.11: Electrocardiogram

| Intervention | Participant Number | Visit | Date/Time (Study Day) | Parameter | Result | Units | Interpretation Detail | Reason Not Done |
|---|---|---|---|---|---|---|---|---|
| XXXXX | XXXXXX | XXX | DDMMMYYYY / HH:MM (XX) | XXX<br>XXX<br>XXX<br>Interpretation [1] | XXX | XXXX | XXXXXXXXX | XXXXXXXXX |

[1] NCS = Not clinically significant, CS = Clinically significant.

*Programming Note: The interpretation will be shown as an additional parameter, with the interpretation outcome shown in the result column.*

E-Sign ID: dc6e1da8-9522-4e0f-8283-c8e65365bf28

Listing 16.2.12: Physical Examination

| Intervention | Participant Number | Visit | Assessment Date/Time (Study Day) | System | Interpretation [1] | Interpretation Detail | Reason not Done |
|---|---|---|---|---|---|---|---|
| XXXXX | XXXXXX | XXX | DDMMMYYYY / HH:MM (XX) | XXX | XXX | XXXXXXXXX | XXXXXXXXX |

[1] NCS = Not clinically significant, CS = Clinically significant.

E-Sign ID: dc6e1da8-9522-4e0f-8283-c8e65365bf28

Listing 16.2.13: Visit Dates

| Intervention | Participant Number | Visit | Date | Source of Unscheduled Visit | Comments |
|---|---|---|---|---|---|
| XXXXX | XXXXXX | XXX | DDMMMYYYY | XXXXXXXXX | XXXXXXXXXXXX |

*Programming note: Unscheduled visits and visit information will be taken from specific database modules and the "Reason for additional collection" will be placed in the comments column in this listing.*

This Document has been Signed with a **secure electronic signature** via E-Sign.

#### Certification Of Completion

| | |
|---|---|
| **Title** | SPL-SPVX02-01 - Final Statistical Analysis Plan |
| **Author** | Laura Grey |
| **Envelope Created on** | Fri, 12 Sep 2025 15:14:43 |
| **Envelope ID** | dc6e1da8-9522-4e0f-8283-c8e65365bf28 |

#### Document Details

| | |
|---|---|
| **Title** | SPL-SPVX02-01_Stablepharma_SAP_Final_1.00_12Sep2025.pdf.pdf |
| **Digital Fingerprint** | 7a4a87e5-25e6-4c0c-bba4-b6f5e8b16a67 |

#### Document Signers Scan/Click the QR Code to view signature information

| | |
|---|---|
| **Name** | Laura Grey |
| **Email** | |
| **Status** | **SIGNED** at Fri, 12 Sep 2025 15:14:56 BST(+0100) |
| **Signature Fingerprint** | 9d93d39b-8d10-4db9-a875-18ef874b950d |
| **Signing Reason** | I am the owner of this document |

| | |
|---|---|
| **Name** | Karen O'Hanlon |
| **Email** | |
| **Status** | **SIGNED** at Tue, 16 Sep 2025 10:06:26 BST(+0100) |
| **Email signer verification** | **** |
| **Signature Fingerprint** | b16b2fe9-43b2-4c6e-b9e2-4b32efd99686 |
| **Signing Reason** | I approve this document |

| | |
|---|---|
| **Name** | Juana De La Torre Arrieta |
| **Email** | |
| **Status** | **SIGNED** at Mon, 15 Sep 2025 09:09:41 BST(+0100) |
| **Email signer verification** | **** |
| **Signature Fingerprint** | 3e51565f-d7bd-49c6-9f3e-f17090b53f05 |
| **Signing Reason** | I approve this document |

British Assessment Bureau
UKAS MANAGEMENT SYSTEMS 8289
ISO 27001 INFORMATION SECURITY MANAGEMENT

#### Signer Events

| | |
|---|---|
| **Tue, 16 Sep 2025 10:06:27** | **Karen O'Hanlon** Signed the Document (IP: 90.222.114.193) |
| **Mon, 15 Sep 2025 09:09:41** | **Juana De La Torre Arrieta** Signed the Document (IP: 84.121.249.69) |
| **Fri, 12 Sep 2025 15:14:56** | Secured by document PIN |
| **Fri, 12 Sep 2025 15:14:56** | **Laura Grey** Signed the Document (IP: 86.144.147.214) |
